# How Does Early, Midday, and Late Time-Restricted Eating Impact Anthropometry and Cardiometabolic Health? A Systematic Review and Network Meta-Analysis of RCTs

**DOI:** 10.64898/2026.01.29.26345140

**Authors:** Mohammed Hamsho, Wijdan Shkorfu, Merve Terzi, Yazan Ranneh, Krista A. Varady, Abdulmannan Fadel

## Abstract

**Background:** Time-restricted eating (TRE) has gained popularity for weight loss and metabolic health. While some evidence suggests greater benefits when TRE aligns with circadian rhythms—characterized by early daytime eating and avoidance of nighttime intake, often referred to as early TRE (eTRE), other studies report no meaningful differences between eTRE, other TRE approaches with or without exercise, or calorie restriction (CR), and robust comparative evidence remains limited.

**Aim:** Therefore, the aim of this network meta-analysis (NMA) is to evaluate the physiological effects of eTRE, midday time-restricted eating (mTRE), late time-restricted eating (lTRE), with and without exercise, CR, and control (without prescribed energy or fasting windows) on anthropometric measures and cardiometabolic markers in adults with cardiometabolic risk factors.

**Methods:** A comprehensive literature search was conducted in four major databases (PubMed, Web of Science, Scopus, and Embase) up to April 24, 2025. A Bayesian NMA was performed, using a control group as the reference comparator across interventions. Treatment effects were expressed as mean differences with 95% confidence intervals. The relative ranking of the included arms on the outcomes was assessed using surface under the cumulative ranking curve, values derived from the NMA, where higher values reflect a higher probability of superior effectiveness.

**Results:** a total of 40 trials comprising 3259 subjects were included in the analysis. There were significant reductions in most anthropometric measures in all intervention groups compared to control group. Whereas eTRE and eTRE + exercise (EX) significantly improved glucoregulatory outcomes compared to control, eTRE + EX showed superior results over other interventions.

**Conclusion:** While our results did not detect statistically significant differences between TRE patterns and CR, the consistent SUCRA rankings in favor of eTRE (particularly with exercise) suggest that meal timing may play an important role in metabolic regulation.

## 1. Introduction

In recent years, time-restricted eating (TRE) has drawn the attention of the scientific community as a form of intermittent fasting that emphasizes the timing of food intake rather than caloric reduction (1). It involves limiting energy intake to a fixed daily window-typically between 6 to 10 hours—followed by prolonged fasting (1). While this approach seems simple in application, scientific evidence suggests that the timing of eating and fasting directly influences human biomarkers (2). Research reveals that TRE exerts significant physiological effects by aligning eating patterns with the endogenous circadian rhythm (3) (2). which orchestrates key biological processes over a 24-hour, including metabolic homeostasis such as insulin secretion, glucose metabolism and energy expenditure, all of which exhibit diurnal variations (4); (5).

The rationale for TRE is grounded in the principles of chronobiology indicating that metabolic efficiency is believed to peak during daytime hours (6); (7); (2) whereas delayed food intake—particularly in the evening or at night—may disrupt circadian alignment. However, modern dietary behaviors are increasingly characterized by irregular meal timing, including frequent breakfast skipping and delayed eating patterns. Such late-phase eating is often marked by prolonged caloric intake, which can promote excess energy consumption and further desynchronize peripheral circadian clocks in metabolically active organs. Thereby altering lipid metabolism, impairing glucose homeostasis, and dysregulating insulin signaling (6). This misalignment contributes to adverse metabolic outcomes and increased risk of various conditions such as metabolic syndrome, diabetes and cardiovascular disorders (3) (2).

In parallel, breakfast skipping has been linked to compensatory increased energy intake later in the day as reported by several observational studies (8,9). As such, TRE represented a targeted behavioral strategy by consolidating food intake within specific windows earlier in the day, thereby aligning energy intake with internal biology. A recent meta-analysis demonstrated that consuming the majority of daily calories intake earlier in the day correlates with lower body weight (BW) and body mass index (BMI) compared with consuming them later in the day (10).

Accordingly, several clinical trials have examined the differential effects of TRE timing using two main models where early TRE (eTRE) restricts meals to early morning through midday, and late TRE or midday TRE (lTRE/mTRE) where meals concentrated from midday through evening hours (11–16).

In our current analysis, we aimed to enhance the precision of dietary timing by categorizing this dietary model (TRE) into three distinct models—early, midday, and late TRE— to examine the impact of meal timing on health outcomes. Existing evidence indicates that early-phase eating models have yielded better outcomes in controlling insulin sensitivity, blood pressure, and oxidative stress, even under isocaloric conditions without weight loss (17). These effects are further potentiated when combined with calorie restriction (CR) (18–20). Additionally, giving that numerous studies have examined the effects of exercise in combination with different models of TRE (21–25), which seems to amplify the effects of TRE, we incorporated exercise as a secondary factor in addition to our approach in isolating meal timing as a factor, allowing for multidimensional analysis of eating and exercise timing.

Therefore, we conducted a systematic review and network meta-analysis (NMA) of randomized controlled trials (RCTs) to evaluate and compare the physiological effects of early (eTRE), mid-day (mTRE), and late (lTRE) time-restricted eating, with and without exercise, as well as calorie restriction (CR), on anthropometric measures and cardiometabolic markers in adults with overweight and obesity. As direct head-to-head comparisons among all interventions were not consistently available, control group commonly used across the included studies served as the reference comparator within the network. To strengthen the interpretation of the findings, the certainty of evidence was assessed using the Grading of Recommendations Assessment, Development, and Evaluation (GRADE) approach.

## 2. Methods

We performed a meta-analysis based on the Preferred Reporting Items for Systematic Reviews and Meta-Analyses (PRISMA) (26). The study protocol was registered in PROSPERO with ID: <u>CRD420251018349</u>. Interventions definitions were clearly explained in **(Supplementary Table 1)**

### 2.1. Search strategy

A search of online databases including PubMed, Web of Science, Scopus, and Embase was conducted up to the cut-off date of 24-4-2025 by two independent authors (A.F. and Y.R.). We systematically searched the literature to identify RCTs that assessed the impact of various TRE interventions on anthropometric measurements, metabolic profile, and blood pressure. We used these key words in our search (“time-restricted eating” OR “time-restricted feeding” OR “tre” OR “time-restricted fasting” OR “early time-restricted eating” OR “late time-restricted eating” OR “Delayed time-restricted eating” OR “intermittent fasting” OR IF) AND (“randomized controlled trial” OR “randomised controlled trial” OR “crossover” OR “RCT” OR “TRIAL” OR random OR randomization OR randomisation) AND (“body mass index” OR “bmi” OR “weight” OR “fat mass” OR “muscle mass” OR “lean body mass” OR “waist circumference” OR “hip circumference” OR “fasting blood glucose” OR “fasting blood insulin” OR Hba1c OR “cholesterol” OR “hdl” OR “ldl” OR “triglyceride” OR “blood pressure” OR “vldl” OR “low density lipoprotein” OR “very low density lipoprotein”).

### 2.2. Eligibility criteria

Studies were included if they met the following criteria: (1) *Population*: adult participants with cardiometabolic risk factor(s), including individuals with overweight (BMI 25.0–29.9 kg/m²), obesity (BMI ≥ 30.0 kg/m²), or normal BMI (18.5–24.9 kg/m²) with hidden obesity (body fat % ≥ 30%), as well as those with cardiometabolic-related chronic conditions, (2) *Intervention*: time-restricted eating (TRE) interventions were eligible if implemented as one of the prespecified forms, including early TRE (eTRE) with the first caloric intake before 10:00 AM, mid-day TRE (mTRE) with the first caloric intake between 10:00 AM and 12:00 PM, or late TRE (lTRE) with the first caloric intake after 12:00 PM, as well as these same TRE schedules combined with structured exercise (eTRE + EX, mTRE + EX, and lTRE + EX). All TRE interventions were required to involve a daily fasting duration of ≥ 14 hours, be implemented on ≥ 6 days per week, and have a minimum intervention duration of 1 month; (3) *Comparison*: control diets without prescribed eating or fasting windows, such as ad libitum intake, usual care, or no intervention; calorie-restricted diets without time restriction; and alternative prespecified TRE schedules (eTRE, mTRE, lTRE, eTRE + EX, mTRE + EX, and lTRE + EX); *Outcomes*: anthropometric measurements, glucose metabolism, lipid metabolism, and blood pressure; *Study Settings*: Randomized controlled trials published in English without date restriction.

In contrast, studies were excluded if they met the following criteria: (1) *Population*: inclusion of pregnant women; professional athletes or highly physically active individuals (defined as participants engaging in structured exercise training for ≥ 6 months prior to study enrollment); night-shift or rotating-shift workers; or studies exclusively enrolling participants with normal BMI and normal body fat percentage who were free from chronic diseases, (2) *Intervention*: eating windows exceeding 10 hours per day or fasting durations of < 14 hours per day; use of self-selected or non-prescribed eating windows; or interventions that included follow-up periods primarily aimed at weight maintenance (e.g. an initial intervention phase followed by a weight-maintenance phase); (3) *Comparison*: prescription of structured exercise to the control group; (4) *Study Settings*: Insufficient data to perform statistical analysis (e.g. only post intervention data availability) **(Table 1).**

**Table 1:** PICO framework table.

| PICOS framework | Inclusion criteria | Exclusion criteria |
| --- | --- | --- |
| Population (P) | A) adult participants with cardiometabolic risk factor(s), including individuals with overweight (BMI 25.0–29.9 kg/m<sup>2</sup>), obesity (BMI ≥ 30.0 kg/m<sup>2</sup>), or normal BMI (18.5–24.9 kg/m<sup>2</sup>) with hidden obesity (body fat % ≥ 30%), as well as those with cardiometabolic-related chronic conditions. | B) Pregnant women<br>C) Performing exercise for at least 6 months prior to study (e.g. professional athletes and reactionary active subjects)<br>D) Night shift workers<br>E) Studies exclusively include participants with normal BMI and normal body fat percentage who were free from chronic diseases. |
| Intervention (I) | A) Time-restricted eating (TRE) implemented as one of the prespecified forms<br>1) early TRE (eTRE): first caloric intake before 10:00 AM;<br>2) mid-day TRE (mTRE): first caloric intake between 10:00 AM and 12:00 PM;<br>3) late TRE (lTRE): first caloric intake after 12:00 PM<br>4) early TRE (eTRE): first caloric intake before 10:00 AM combined with exercise (eTRE + EX);<br>5) midday TRE (mTRE): first caloric intake between 10:00 AM and 12:00 PM combined with exercise (mTRE + EX),<br>6) ) late TRE (lTRE): first caloric intake after 12:00 PM combined with exercise (lTRE + EX),<br>all defined by a daily fasting duration of ≥14 hours implemented on ≥6 days per week, with or without exercise. | A) Eating window > 10 hours or fasting window <14 hours a day.<br>B) Self-selected eating windows.<br>C) Follow up periods aiming for weight maintenance (e.g. 3 months intervention and 3 months follow up weight maintenance) |
| Comparison (C) | A) Control conditions without prescribed eating or fasting windows (e.g., ad libitum intake, usual care, or no intervention), calorie restriction without time restriction, or alternative TRE schedules (eTRE, mTRE, lTRE, eTRE + EX, mTRE + EX, or lTRE + EX). | A) prescription of structured exercise to the control group. |
| Outcome (O) | A) Anthropometric measurements, glucose metabolism, lipid metabolism, and blood pressure. | Not applicable |
| Study setting (S) | A) Randomized controlled trials (RCTs)<br>B) Minimum duration of 1 month | A) Insufficient data to perform statistical analysis (e.g. only post intervention data availability). |
| Language | English language published articles |  |
| Date | No limits |  |

Footnotes: , eTRE = early time-restricted eating, mTRE = midday time-restricted eating, lTRE = late time-restricted eating, eTRE + EX = early time-restricted eating + exercise, mTRE + EX = midday time-restricted eating + exercise,

### 2.3. Outcomes

There were 18 prespecified outcomes in our present study: 1) BW (kg), 2) BMI (kg/m^2^), 3) fat%, 4) fat mass (kg), 5) lean body mass (LBM) (kg), 6) waist circumference (WC) (cm), 7) hip circumference (HC) (cm), 8) waist-hip ratio (WHR), 9) fasting blood glucose (FBG) (mg/dl), 10) fasting blood insulin (FBI) (mIU/L), 11) Homeostatic Model Assessment for Insulin Resistance (HOMA-IR), 12) hemoglobin A1C (HbA1c) (%), 13) triglyceride (TG) (mmol/l), 14) total cholesterol (TC) (mg/dl), 15) low density lipoprotein cholesterol (LDL-C) (mg/dl), 16) high density lipoprotein cholesterol (HDL-C) (mg/dl), 17) systolic blood pressure (SBP) (mmHg), and 18) diastolic blood pressure (DBP) (mmHg). The NMA was conducted for all outcomes.

### 2.4. Study selection

Study selection was performed independently and in duplicate by two reviewers (M.H. and W.S). In the first stage, titles and abstracts of all retrieved records were screened according to the predefined eligibility criteria. In the second stage, the full texts of potentially eligible articles were assessed in detail against the inclusion and exclusion criteria. Any discrepancies between reviewers were resolved through discussion or, when necessary, by consultation with a third senior investigator (Y.R.). The study selection process was documented using a PRISMA flow diagram, and reasons for exclusion were recorded at each stage.

### 2.5. Data extraction

After the selection process of articles, with regard to the inclusion and exclusion criteria, the following data were extracted by two independent reviewers (W.S. and M.T.) from each eligible study: study characteristics (first author, study location, and study design), participant characteristics (sample size, age, population BMI and health condition), intervention details (intervention(s) and amount of CR, and control and amount of CR, and intervention duration) and listed in **(Table 2)**.

**Table 2:**
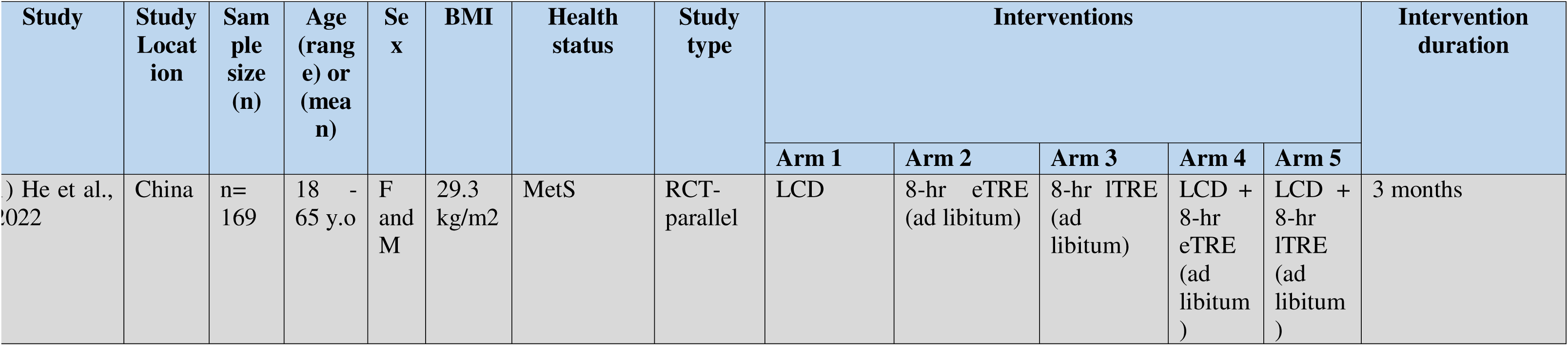
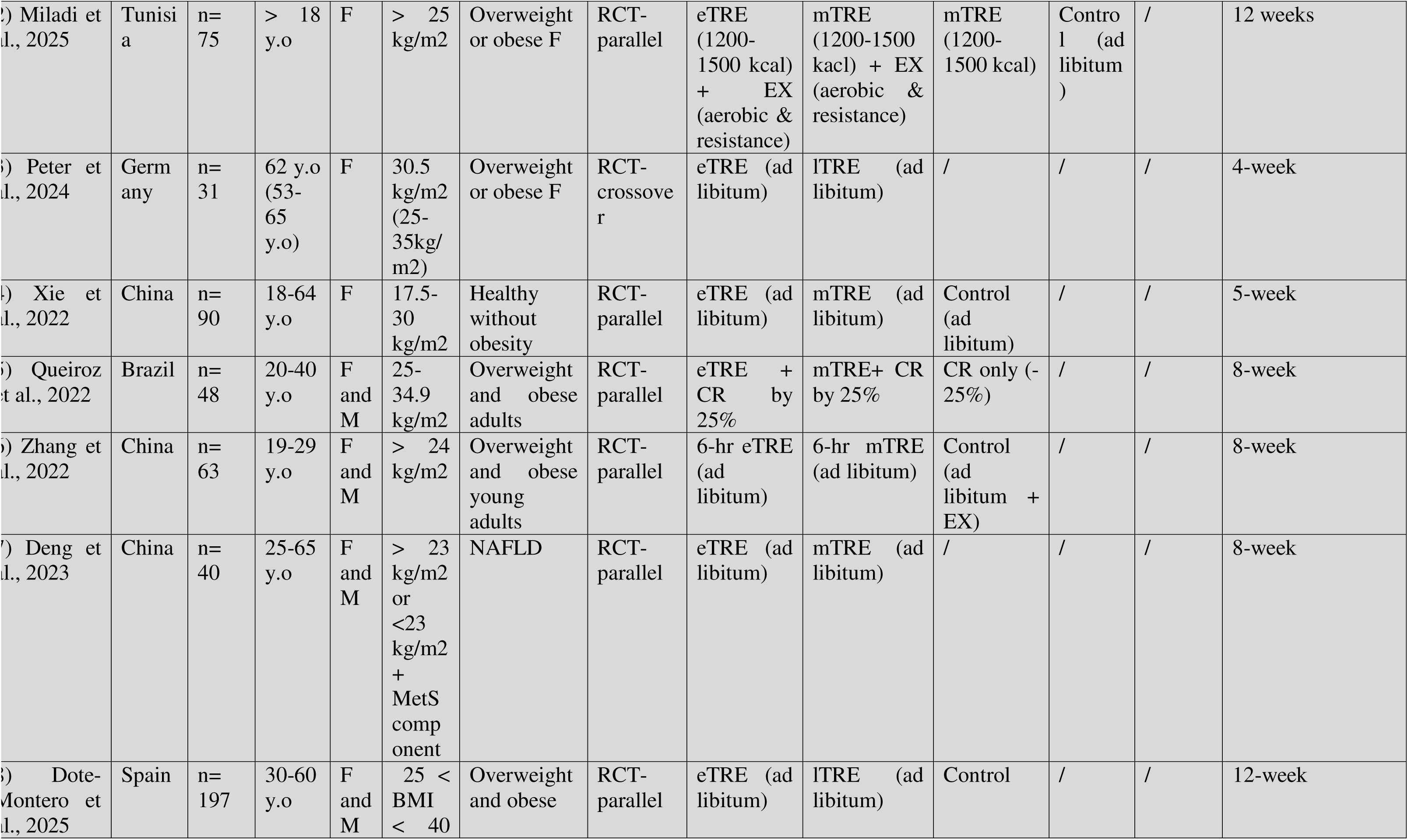
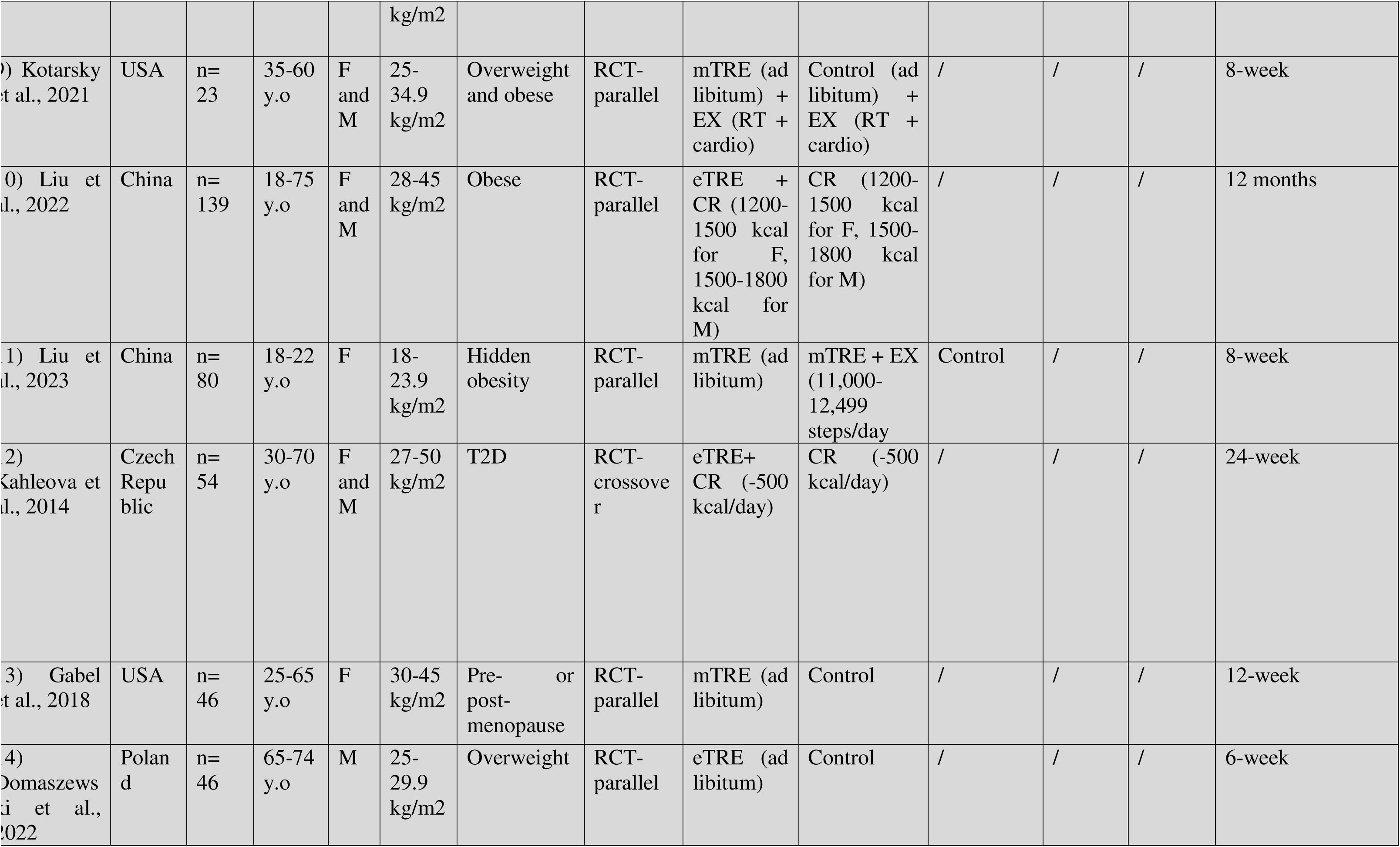
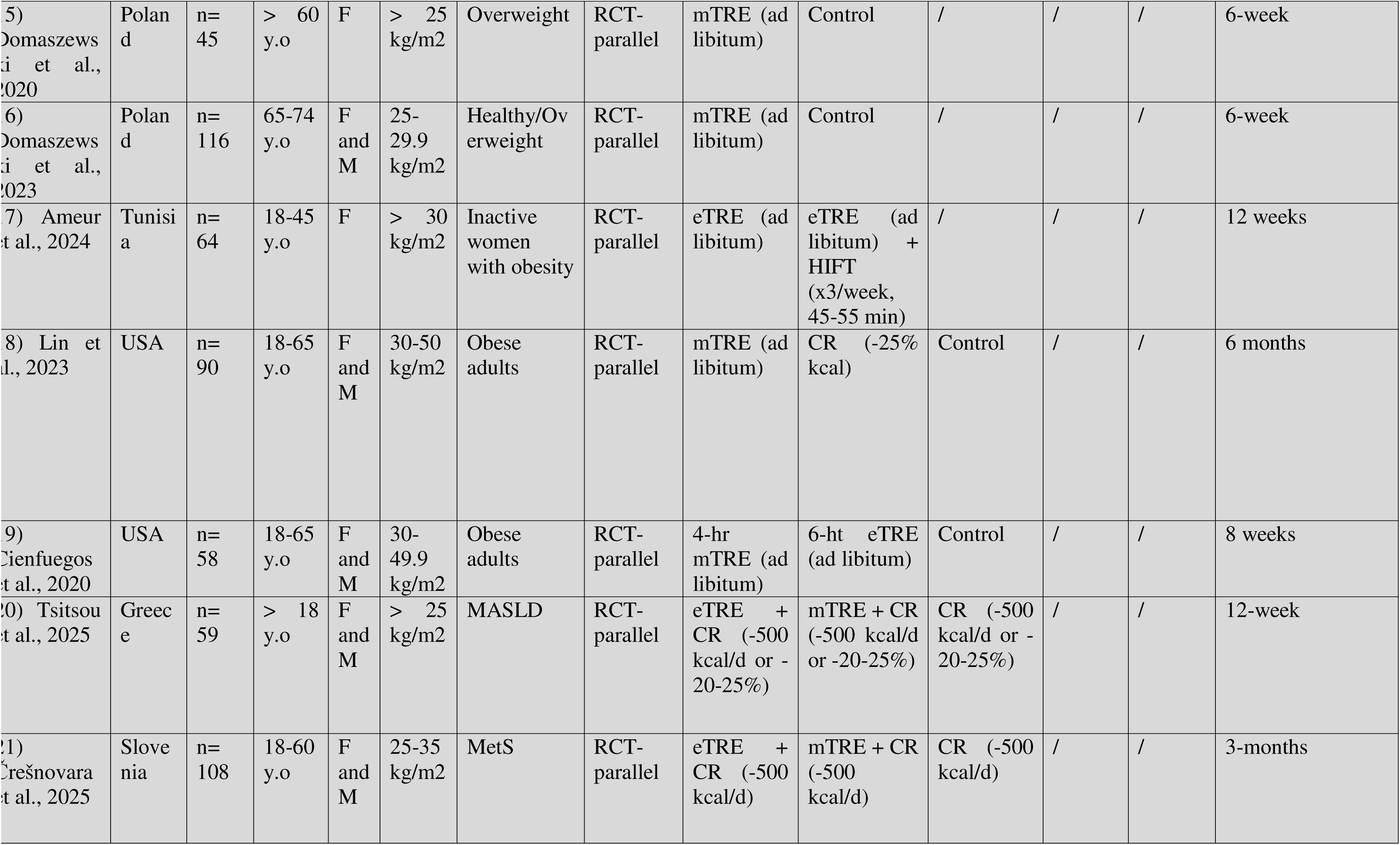
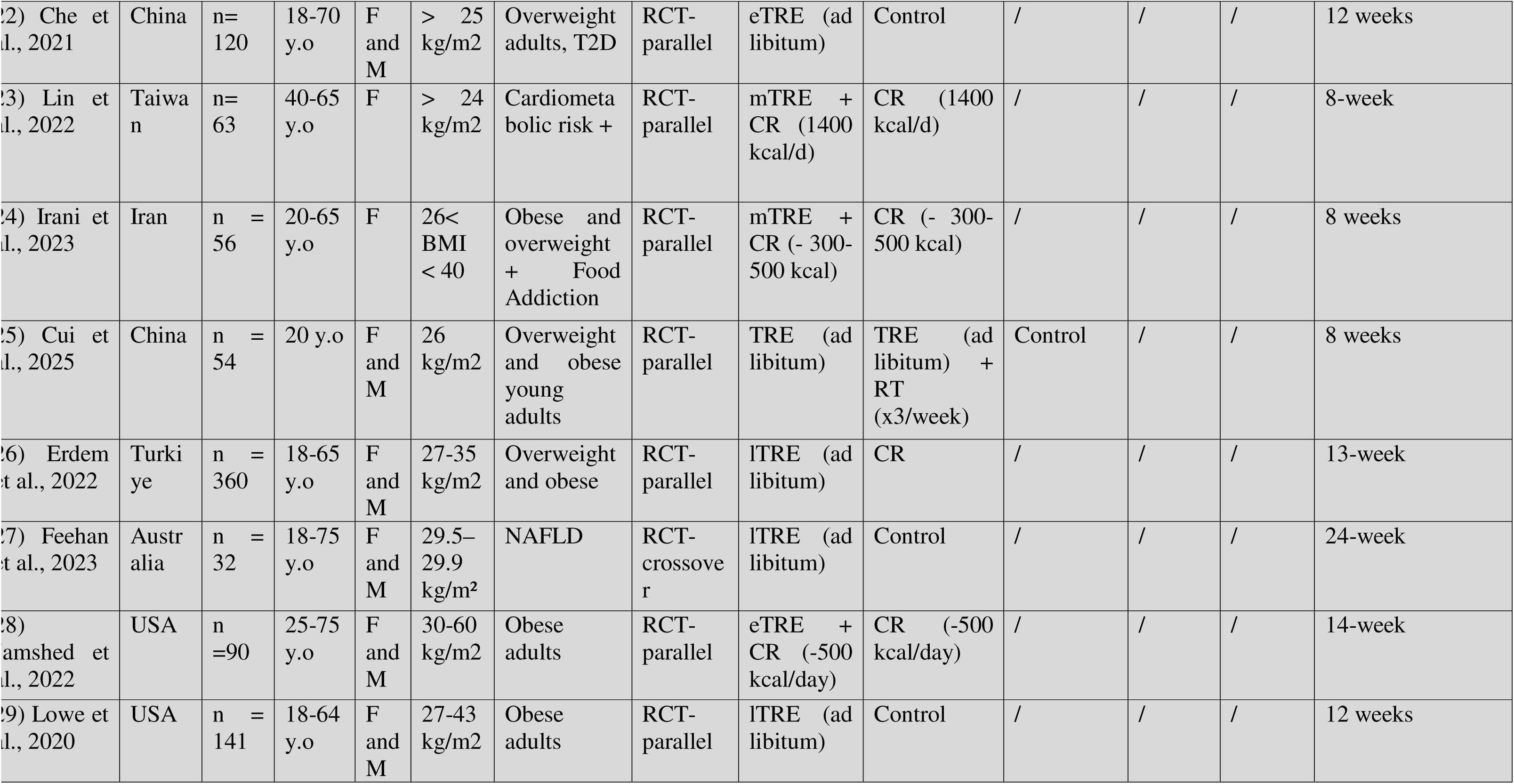
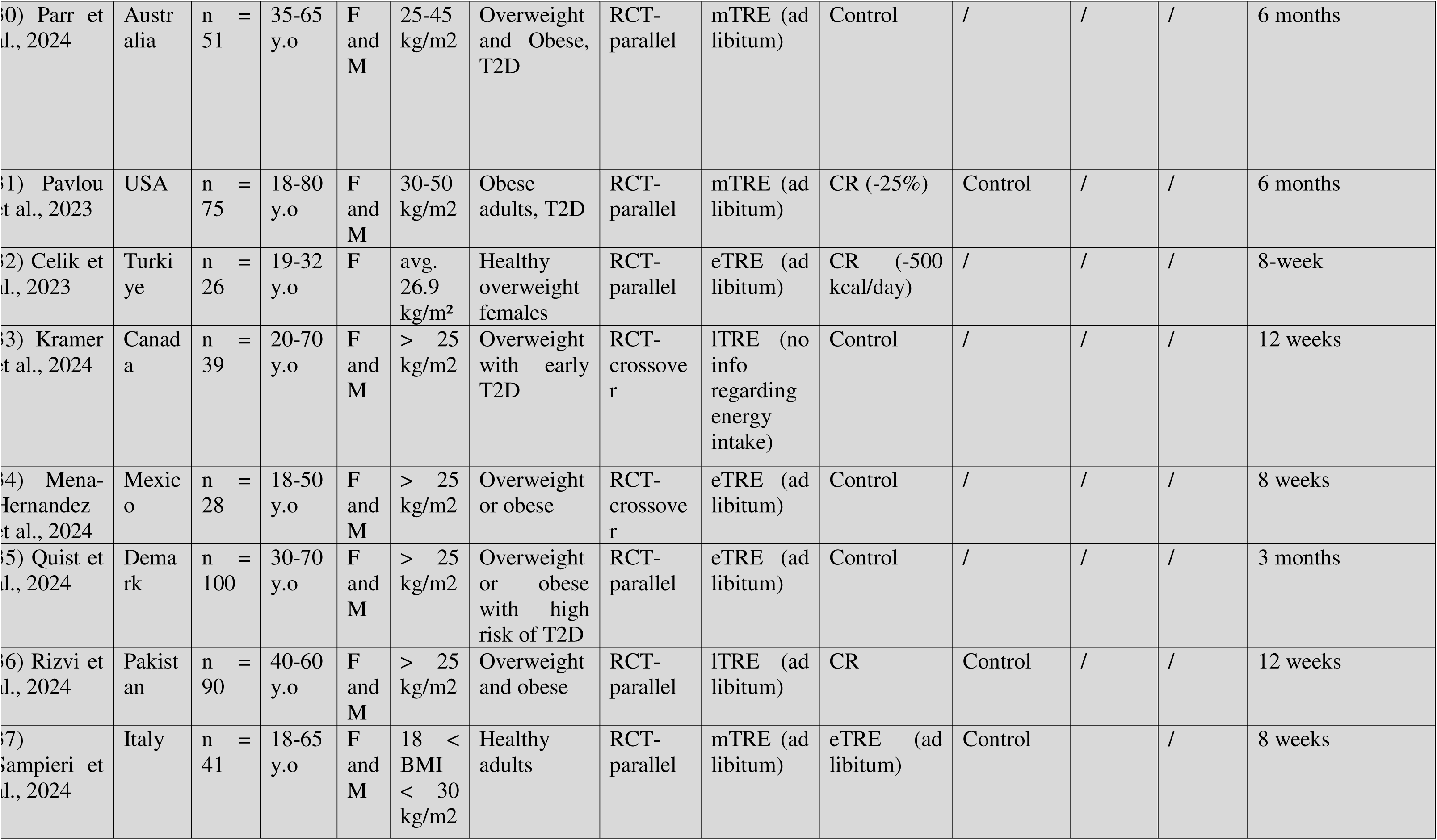
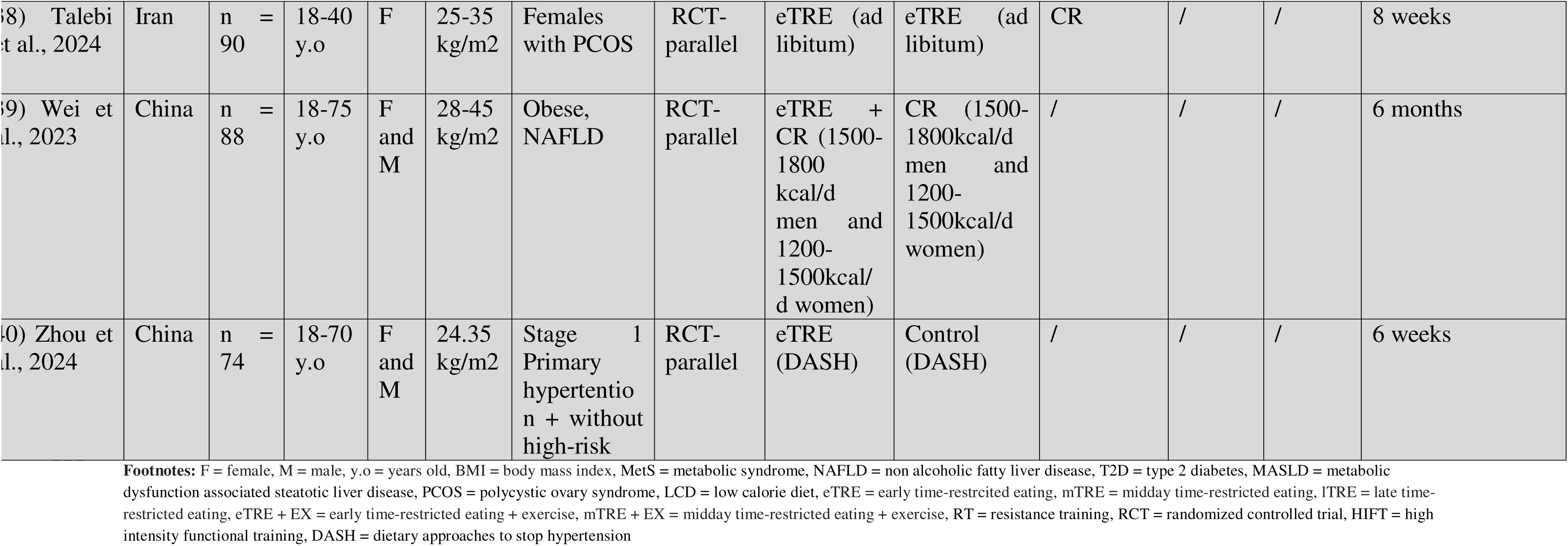
Characteristics of included studies.

For the assessment of intervention effects, prespecified outcome data were extracted for each intervention and comparator arm, including mean changes from baseline and corresponding standard deviation (SD), or these values were calculated using appropriate statistical methods when not directly reported (explained in section 2.7).

### 2.6. Risk of bias assessment

The quality assessment was performed by two independent authors (M.H and W.S), any disagreement between the authors was solved by third author (Y.R). The quality of the studies included was evaluated using the Cochrane ROB-2 tool. This tool includes the following key parts: random sequence generation, allocation concealment, blinding, incomplete outcome data, and selective reporting. Each item was categorized as having low, unclear, or high risk of bias. Accordingly, studies with more than two items of low risk were categorized as studies with good quality, studies with two items of low risk were considered studies with fair quality, and those with fewer than two items were considered studies with low risk of bias (27).

### 2.7. Certainty of evidence

The confidence in Network Meta-Analysis (CINeMA) tool, based on GRADE approach with a Bayesian framework, was applied to evaluate the quality of evidence across a network. In addition, given in the CINeMA framework, qualitative judgments are made across 6 domains: within-study risk of bias, reporting bias, indirectness, imprecision, heterogeneity, and incoherence (28). Briefly, within study bias evaluates the potential flaws in the study design or execution that can lead to a systematic difference between the estimated relative treatment effect and the true effect, as evaluated using the risk of bias tool. Reporting bias refers to when the results in a systematic review do not accurately represent all the findings from the studies conducted. This bias can occur when non-significant findings or unfavorable results are not reported (publication bias or outcome reporting bias). Indirectness assesses the issue of transitivity, referring to the assumption that trials comparing different interventions are similar in terms of important characteristics that might influence the effect estimate, including age, sex, sample size, and study duration (29,30). Imprecision, heterogeneity, and incoherence evaluate the reliability and validity of the results. Confidence rating can be high, moderate, low, or very low (31). Minimal important differences (MIDs) of clinically important changes for all outcomes were adopted from previously published NMAs **(Supplementary Table 2)** (32,33).

### 2.8. Data synthesis and analysis

The Bayesian framework method was used to conduct NMA of RCTs using Metainsight (34), comparing different TRE approaches with and without exercise, CR, and control. Random effect models in our study were used to calculate summary weighted mean differences and 95% confidence intervals (95% CIs), and to assess the heterogeneity between studies (35). Change from baseline of mean and corresponding SD were calculated from the included studies to run the statistical analysis. In any case, reporting the standard error of the mean (SEM), standard deviation (SD) was calculated using the following formula: SD = SEM × sqrt (n), where n refers to the number of participants. When studies didn’t report the SD of the change from baseline, it was calculated using the following formula:

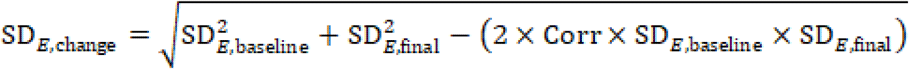

Where R represents the correlation coefficient and was assumed to be 0.9. This assumption was empirically derived from studies that provided sufficient data to calculate this correlation. Subsequently, the same value was imputed to the remaining studies where insufficient data was provided. This approach is consistent with guidance from the Cochrane Handbook, which recommends estimating the correlation coefficient from available studies when it is not directly reported (36). If the upper and lower limits were given with the mean, the SD was calculated using this formula: *SD=n×(upperlimit−lowerlimit)/3.92*. If the median with upper and lower limits was given, the estimation was based on the method described here (37).

For crossover trials in which individual participant data were not available, and where carry-over or period effects were assumed negligible, all measurements from each intervention period were extracted and analyzed as if the trial had a parallel-group design. This method is considered acceptable under these conditions as discussed earlier (38).

To allow pooling using mean differences, outcomes reported in different measurement units were standardized by converting them to a common unit using appropriate equations, when applicable. In case more than one group of the same category were included in a study, their data were combined as Cochrane handbook suggests. The random-effects model was applied for pooling analysis to compensate for the heterogeneity of the studies (27,39). Interstudy heterogeneity was explored quantitatively using Cochran’s Q and I^2^ statistics. I^2^ ≤ 50 % and ≥75 % indicated substantial and considerable heterogeneity, respectively (27) P-values were considered statistically significant at < 0.05.

League tables present network meta-analysis estimates for all pairwise comparisons. Values represent mean differences with corresponding 95% confidence intervals. Each cell compares the intervention in the row against the intervention in the column. Negative values indicate a greater reduction in the outcome favoring the row intervention, while positive values favor the column intervention. Results are considered statistically significant when the interval does not include zero.

The relative ranking of the included arms for BW, BMI, fat %, fat mass, LBM, WC, HC, WHR, FBG, FBI, HOMA-IR, HbA1c, TG, TC, LDL-C, HDL-C, SBP, and DBP levels was assessed using surface under the cumulative ranking curve (SUCRA), values derived from the NMA, where higher values reflect a higher probability of superior effectiveness. We assumed a common heterogeneity parameter in the NMA aimed to compare with the empirical distributions of heterogeneity (40). Therefore, we evaluated local inconsistency based on the node-splitting method (41). Prespecified subgroup analyses were conducted on the following baseline variables: energy intake (*ad libitum* and energy prescription), and duration of intervention (longer term ≥ 11 weeks and shorter term ≤ 10 weeks). Sensitivity analyses were performed by excluding studies that included subjects with normal BMI, and studies on healthy subjects. Forest plots for the NMA of all the outcomes are provided in supplementary materials **(From Supplementary Figure 3 to Supplementary Figure 20)**.

## 3. Results

### 3.1. Screening results of RCTs

The initial search resulted in 1926 articles, and 1393 duplicates were removed. Following initial screening, at the title and abstract level, 380 articles were excluded. It should be noted that the lTRE + EX node was not included in the network, as no published trials evaluating this intervention were available. After a full-text review of the 154 articles, 113 were excluded for various reasons as described in **(Figure 1)**. Thus, 40 articles were included in this NMA.

**Figure 1:**
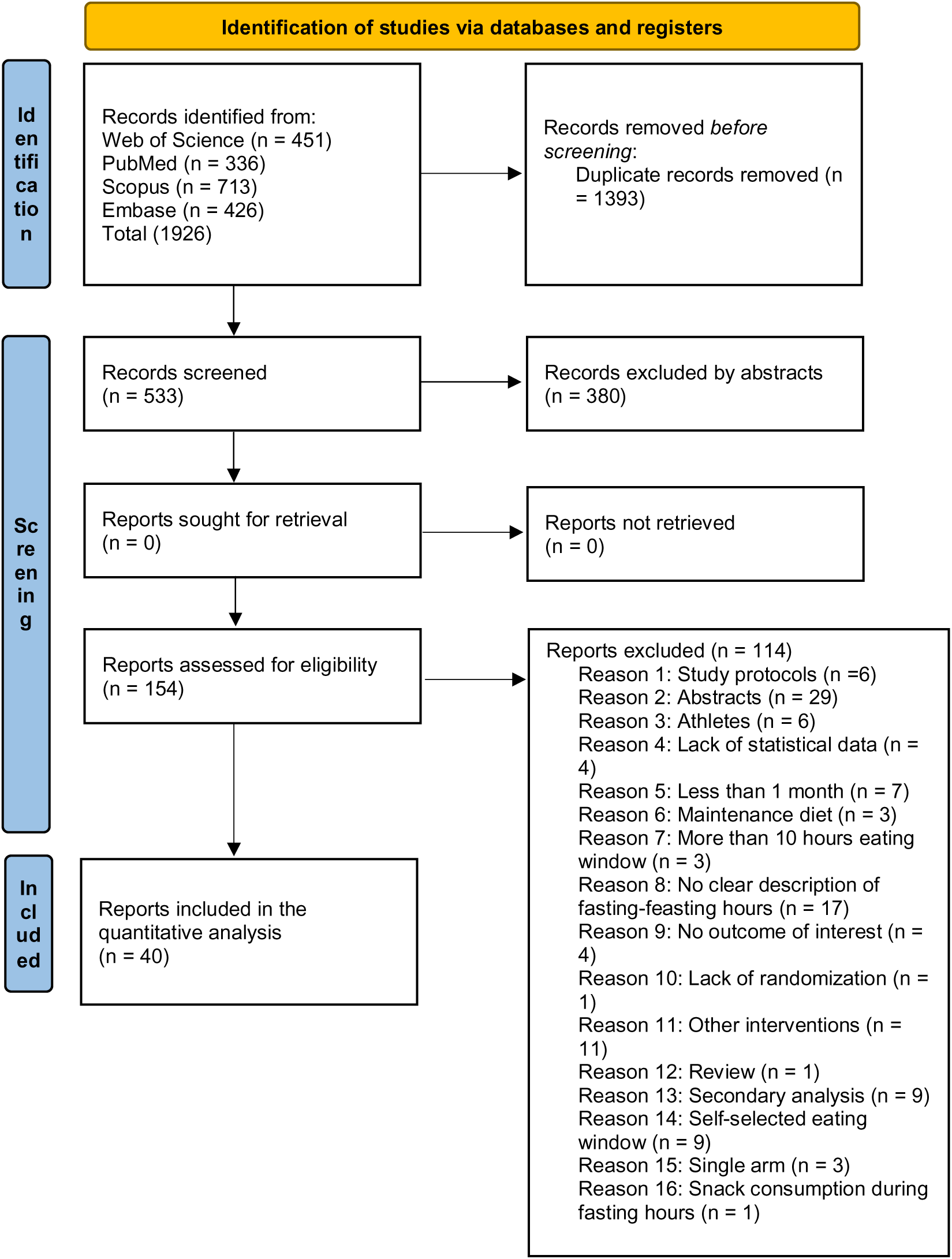
Preferred Reporting Items for Systematic review and Meta analysis (PRISMA) diagram.

### 3.2. Studies’ characteristics

**Table 2** represents the detailed characteristics of the 40 included study. Overall, the age of participants ranged from 18 to 75 years. Most studies included both genders, but 10 (25%) were exclusive to females and 1 (2.5%) to males. The health condition of adults varied among studies, metabolic syndrome (N = 2), T2D (N = 5), non-alcoholic fatty liver disease (N = 4), polycystic ovary syndrome (N = 1), food addiction (1 report), hypertension (N = 1), cardiometabolic diseases risk (N = 2), hidden obesity (N = 1), with mostly included overweight or obese adults (40 trials). Out of the included studies, 5 of them were crossover RCTs and 35 parallel RCTs. Moreover, eTRE was applied in 24 (*ad libitum* 16, energy prescription 9), mTRE in 20 (*ad libitum* 14, energy prescription 6), lTRE in 8 (*ad libitum* 8, energy prescription 1), eTRE + EX in 3, mTRE + EX in 4, CR in 18, and control in 25 reports. Geographical locations of the included studies were diverse by which 10 of them were conducted in China (11,12,14,21,24,42–46), 7 in USA (18,47–52), 3 in Poland (53–55), 2 in Tunisia (22,56), 2 in Iran (57,58), 2 in Turkey (59,60), 2 in Australia (61,62), and 1 study in each of the following countries: Germany (13), Japan (63), Spain (15), Czech Republic (Kahleova et al., 2014), Greece (16), Slovenia (20), Taiwan(65), Canada (66), Mexico (67), Denmark (68), Pakistan (69), and Italy (70). Finally, trials duration ranged from 4 weeks to 12 months.

### 3.3. Risk of bias assessment

In terms of risk of bias, 12 trials were categorized with an unclear risk of bias, 24 low risk of bias, and 4 high risk of bias as shown in risk of bias graph **(Supplementary Figure 1)** and risk of bias summary **(Supplementary Figure 2)**. Blinding of participants and personnel domain was assessed as high risk of bias due to the nature of interventions. Studies that didn’t provide clear randomization methods were assessed as unclear. Studies expressed their data as intention to treat analysis were assessed as low risk.

### 3.4. Network diagrams

The 40 included studies covered seven interventions: Control, CR, eTRE, eTRE + EX, mTRE, mTRE + EX, and lTRE. The network for direct comparison of interventions and control is shown in **(Figure 2)**. In the figure, the thickness of the lines is proportional to the number of studies included in the pairwise comparison, and the diameter of the circles is proportional to the number of participants who received the intervention.

**Figure 2:**
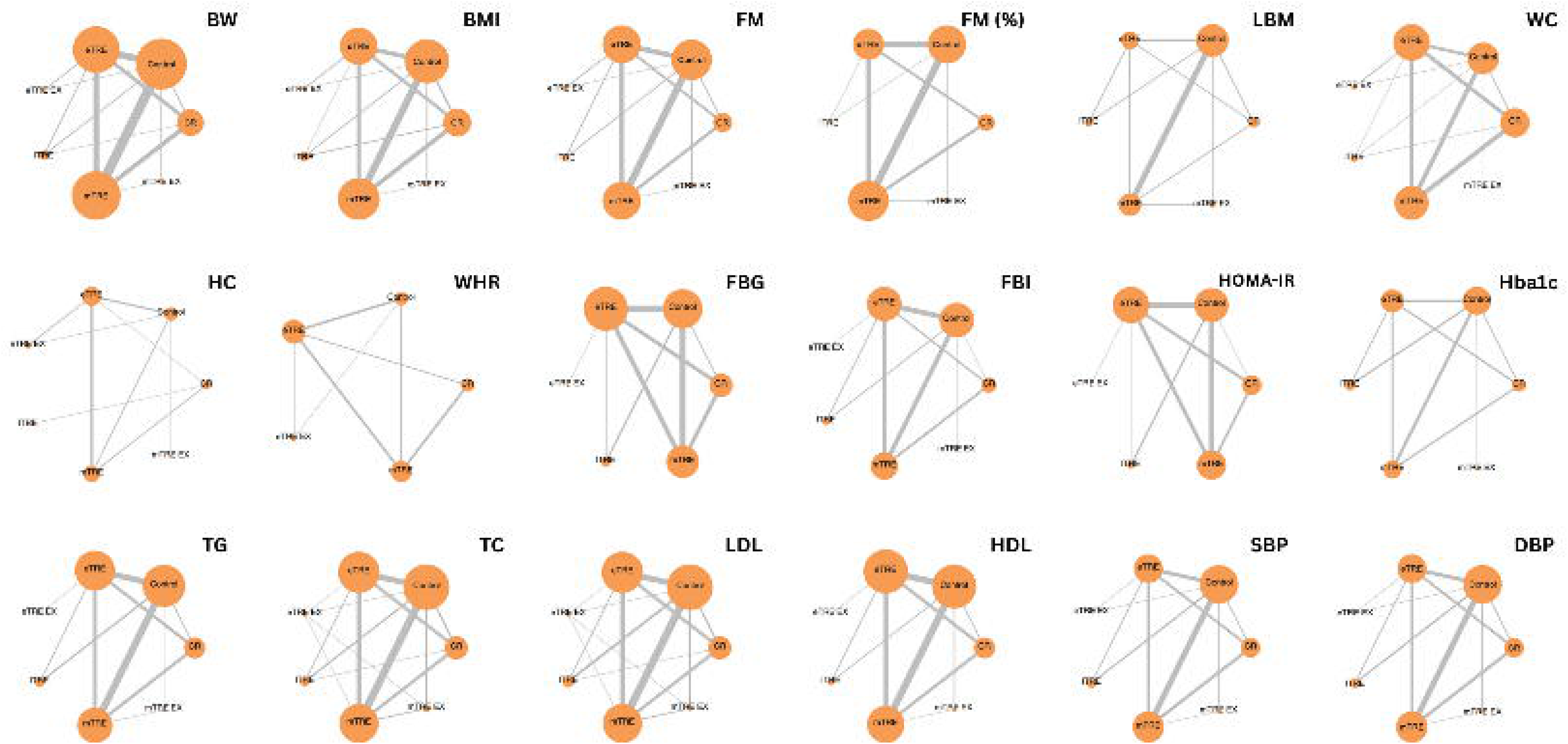
Network of eligible comparisons for all outcomes, Network plots of eligible direct comparisons. The width of the lines is proportional to the number of trials comparing each pair of treatments. The size of the nodes is proportional to the number of participants involving the specific intervention.

### 3.5. Anthropometric measurements

A total of 37 studies reported weight changes during the study. All the interventions were significantly (p < 0.05) more effective than control group as shown in **(Supplementary Figure 3)**. eTRE + EX has produced the highest weight loss among the interventions (MD = −3.20 kg, 95% CI: −4.83, −1.63). The SUCRA rankings from the highest to lowest were eTRE + EX (94%), eTRE (69%), lTRE (55%), mTRE + EX (47%), and mTRE and CR were similar (42%) as shown in **(Table 3)**.

**Table 3:** Surface Under The Cumulative Ranking (SUCRA).

| Treatment | Body weight | BMI | Fat mass (kg) | Fat (%) | Lean mass | WC | HC | WHR | FBG | FBI | HOMA-IR | HbA1c (%) | TG | TC | LDL-C | HDL-C | SBP | DBP |
| --- | --- | --- | --- | --- | --- | --- | --- | --- | --- | --- | --- | --- | --- | --- | --- | --- | --- | --- |
| Control | 0.32 | 0.34 | 2 | 21 | 17 | 6 | 45 | 27 | 13 | 17 | 15 | 22 | 35 | 37 | 61 | 33 | 18 | 36 |
| CR | 42 | 34 | 41 | 62 | 40 | 66 | 53 | 61 | 52 | 40 | 35 | 73 | 55 | 53 | 58 | 23 | 48 | 12 |
| eTRE | 69 | 58 | 46 | 66 | 75 | 66 | **68** | **90** | 75 | 62 | 63 | **90** | 40 | 40 | 56 | 36 | 70 | 66 |
| eTRE + EX | **94** | **84** | **99** | N/A | N/A | **85** | 11 | 35 | **93** | **98** | **94** | N/A | **100** | **100** | **100** | **100** | 52 | **69** |
| ITRE | 55 | 72 | 42 | 70 | **76** | 42 | 52 | N/A | 42 | 75 | 69 | 66 | 61 | 59 | 43 | 38 | 41 | 66 |
| mTRE | 42 | 50 | 59 | **74** | 25 | 35 | 63 | 38 | 24 | 40 | 24 | 45 | 11 | 13 | 16 | 72 | 36 | 68 |
| mTRE + EX | 47 | 53 | 61 | 5 | 68 | 52 | 57 | N/A | N/A | 18 | N/A | 4 | 47 | 47 | 17 | 47 | **85** | 34 |

**Footnotes:** eTRE = early time-restricted eating, mTRE = midday time-restricted eating, ITRE = late time-restricted eating, eTRE + EX = early time-restricted eating + exercise, mTRE + EX = midday time-restricted eating + exercise, BMI = body mass index, WC = waist circumference, HC = hip circumference, WHR = waist to hip ratio, FBG = fasting blood glucose, FBI = fasting blood insulin, HOMA-IR = homeostatic model assessment for insulin resistance, HbA1c = hemoglobin A1C, TG = triglyceride, TC = total cholesterol, LDL-C = low density lipoprotein cholesterol, HDL-C = high density lipoprotein cholesterol, SBP = systolic blood pressure, DBP = diastolic blood pressure.

BMI was assessed by 32 studies. Similar to weight loss, all the interventions were significantly (p < 0.05) more effective than control group in reducing BMI **(Supplementary Figure 4)**. The highest reduction was observed in eTRE + EX (MD = −1.22 kg/m^2^, 95% CI: −2.01, −0.455). The SUCRA rankings from the highest to lowest were eTRE + EX (84%), lTRE (72%), eTRE (58%), mTRE + EX (53%), mTRE (50%), and CR (34%) **(Table 3)**.

Fat mass (kg) was assessed by 27 studies. Except for mTRE + EX, all the interventions reached statistical significance (p < 0.05) for fat mass reduction compared to control group with the highest effect for eTRE + EX (MD = −4.64 kg, 95% CI: −6.20, −3.15) **(Supplementary Figure 5)**. In addition, eTRE + EX significantly reduced fat mass compared to the other intervention groups, but not compared to mTRE + ex. The SUCRA ranking from the highest to lowest were eTRE + EX (99%), mTRE + EX (61%), mTRE (59%), eTRE (46%), lTRE (42%), and CR (41%) (Table 3).

For fat (%) assessment, the number of studies was diminished to 24 studies with the absence of eTRE + EX intervention. mTRE and eTRE, but not the other groups, were found to significantly reduce fat (%) compared to control group (MD = −0.80 %, 95% CI: −1.38, −0.298), and (MD = −0.72 %, 95% CI: −1.30, −0.141) respectively **(Supplementary Figure 6)**. The SUCRA ranking from the highest to lowest were mTRE (74%), lTRE (70%), eTRE (66%), CR (62%), control (21%), and mTRE + EX (5%) **(Table 3)**.

Lean body mass (kg) was reported by a total of 16 studies including all the interventions except eTRE + EX, lTRE and eTRE produced significant reductions (MD = −0.79 kg, 95% CI: −1.47, −0.023), and (MD = −0.74 kg. 95% CI: −1.26, −0.049) compared to control group, respectively **(Supplementary Figure 7)**. The SUCRA ranking from the highest to lowest were lTRE (76%), eTRE (75%), mTRE + EX (68%), CR (40%), and mTRE (25%) **(Table 3)**.

Furthermore, 27 studies have reported the impact of the included interventions on WC. Except for lTRE and mTRE + EX, all the interventions reported significant reduction, compared to control group, in WC with the highest reduction for eTRE + EX group (MD = −4.03 cm, 95% CI: −7.39, −0.581) **(Supplementary Figure 8)**. The SUCRA ranking from the highest to lowest were eTRE + EX (85%), eTRE, and CR (66%), mTRE + EX (52%), lTRE (42%), and mTRE (35%) **(Table 3)**.

Only 11 studies assessed HC change during the study. None of the interventional groups produced significant reduction compared to control group **(Supplementary Figure 9)**. However, the highest SUCRA ranking was achieved by eTRE group (68%) **(Table 3)**.

WHR was also measured by a few studies (n=13) with the absence of lTRE and mTRE + EX groups. Additionally, no significant effect was found compared to control group **(Supplementary Figure 10)**. The highest SUCRA ranking was achieved by eTRE group (90%) (Table 3).

### 3.6. Glucose metabolism

FBG was reported by a total of 28 studies. Compared to control group, a significant reduction was reported by the eTRE group (MD = −3.64 mg/dl, 95% CI: −5.93, −1.36) **(Supplementary Figure 11)**. Despite this, the SUCRA ranking reveals the highest ranking for eTRE + EX (93%) followed by eTRE (75%), and CR (52%) **(Table 3)**.

Of the included studies, 22 have assessed the impact of the interventions on the FBI **(Supplementary Figure 12)**. There was significant reduction in eTRE + EX (MD = −5.41, 95% CI: −9.35, −1.74, SUCRA (98%)), lTRE (MD = −2.48 (mIU/L), 95% CI: −5.08, −0.0974, SUCRA (75%)), and eTRE (MD = −1.52 (mIU/L), 95% CI: −2.96, −0.260, SUCRA (62%)) compared to control group **(Table 3).**

HOMA-IR was measured by 23 studies with the absence of mTRE + EX group. There was a significant reduction in eTRE + EX (MD = −1.33, 95% CI: −2.58, −0.0812) and eTRE (MD = −0.425, 95% CI: −0.832, −0.03) groups, compared to control group **(Supplementary Figure 13)**. The SUCRA ranking from the highest to lowest were eTRE + EX (94%), lTRE (69%), eTRE (63%), CR (35%), and mTRE (24%) **(Table 3)**.

Sixteen studies reported HbA1c with the absence of eTRE + EX **(Supplementary Figure 14)**. Compared to control group, significant reduction was only produced by eTRE (MD = −0.380 %, 95% CI: −0.623, −0.160, SUCRA (90%)) and CR (MD = −0.296 %, 95% CI: −0.611, −0.00714, SUCRA (73%)) groups **(Table 3)**.

### 3.7. Lipid metabolism

A total of 23 studies reported the change in TC. A significant reduction was found in eTRE + EX group, compared to control group (MD = −27.8 mg/dl, 95% CI: −38.3, −17.2), while no effect on TC levels was seen in the other groups **(Supplementary Figure 15)**. The SUCRA ranking from the highest to lowest were eTRE + EX (100%), lTRE (59%), CR (53%), mTRE + EX (47%), control (37%), and mTRE (13%) **(Table 3)**.

LDL-C was assessed in a total of 31 studies. Similar to TC, only eTRE + EX was found to significantly reduce LDL-C levels compared to control group (MD = −14.9 mg/dl, 95% CI: −24.2, −5.64) **(Supplementary Figure 16)**. On the other hand, mTRE exerted a significant increase in LDL-C levels (MD = 3.99 mg/dl, 95% CI: 0.438, 7.90) compared to control group. There was no difference among the other groups. The SUCRA ranking from the highest to lowest were eTRE + EX (100%), control (61%), CR (58%), eTRE + EX (56%), lTRE (43%), mTRE + EX (17%), and mTRE (16%) **(Table 3)**.

HDL-C was measured in a total of 30 studies. Compared to control group, no significant difference was found among the groups **(Supplementary Figure 17)**. However, eTRE + EX group had significantly higher HDL-C levels compared to control group (MD = 19.4 mg/dl, 95% CI: 12.1, 26.5). The SUCRA ranking from the highest to lowest were eTRE + EX (100%), mTRE (72%), mTRE + EX (47%), lTRE (38%), eTRE (36%), control (33%), and CR (23%) (Table 3).

TG was reported in 28 studies. A significant reduction was found in eTRE + EX group (MD = −0.35 mmol/l, 95% CI: −0.65, −0.05) compared to control group **(Supplementary Figure 18)**. No other difference was found among the groups. The SUCRA ranking from the highest to lowest were eTRE + EX (100%), lTRE (61%), CR (55%), mTRE + EX (47%), eTRE (40%), control (35%), and mTRE (11%) **(Table 3)**.

### 3.8. Blood pressure

A total of 24 studies examined the impact of the interventions on SBP. Only eTRE was found to significantly reduce SBP compared to control group (MD = −2.57 mmHg, 95% CI: −5.18, −0.0207) **(Supplementary Figure 19)**. The SUCRA ranking from the highest to lowest were mTRE + EX (85%), eTRE (70%), eTRE + EX (52%), CR (48%), lTRE (41%), and mTRE (36%) (**Table 3**).

Similar studies assessed and reported DBP. However, none of the interventions produced significant changes in DBP compared to the control group **(Supplementary Figure 20)**. The SUCRA ranking from the highest to lowest were eTRE + EX (69%), mTRE (68%), eTRE and lTRE (66%), control (36%), mTRE + EX (34%), and CR (12%) (**Table 3**).

### 3.9. Inconsistency

Node splitting results are presented in the supplementary materials **(From Supplementary Figure 21 to Supplementary Figure 36)**. No indication was observed for inconsistency with the node-splitting approach regarding FM (kg), LBM (kg), WC, WHR, FBG, FBI, HOMA-IR, HbA1c, HDL-C, TG, SBP, and DBP **(Supplementary Figures 23, 25, 26, and 28-36)**. However, some significant inconsistencies were observed by the lTRE vs control for BW and BMI **(Supplementary Figures 21 and 22)**, eTRE vs control, mTRE vs control, CR vs eTRE for fat (%) **(Supplementary Figure 24)**, and eTRE + EX vs control for HC **(Supplementary Figure 27)**. In addition, node-splitting method wasn’t performed on TC and LDL-C because there are no closed loops.

### 3.10. Subgroup and sensitivity analysis

In supplementary materials, we outlined the results of the subgroup **(From Supplementary Table 3 to Supplementary Table 27)** and sensitivity analysis where applicable. The findings from subgroup analysis we planned a prior for energy prescription reveal more pronounced improvements in eTRE and mTRE compared to control regarding BW, BMI, fat (%), FM (kg), FBI, and HOMA-IR in eTRE group when energy intake was *ad libitum*. In contrast, WC reduction and improvements of lipid profile were slightly better when energy intake was prescribed. Interestingly, CR group from studies in which TRE was prescribed without intentional energy restriction (ad libitum) exhibited superior outcomes compared with CR group from studies where TRE participants were instructed to restrict energy intake. These findings suggest that differences in study design or intervention context may have contributed to the observed outcomes, rather than the dietary strategy alone.

Subgroup analysis based on intervention duration overall reveals that shorter durations are associated with better outcomes for all groups, but not eTRE + EX. This could be explained by better dietary adherence during short interventions, which declines in longer studies. Detailed results can be found **(From Supplementary Table 3 to Supplementary Table 27).**

Sensitivity analysis based on both removal of studies with normal BMI and healthy subjects didn’t significantly differ from the first analysis in most outcomes. This might be due to the low number of these studies (N=4), causing minimal effect on the overall results. Detailed results can be found **(From Supplementary Table 28 to Supplementary Table 44).**

### 3.11. Credibility of the evidence

The GRADE certainty of evidence for all diet group comparisons are presented **(From Supplementary Table 45 to Supplementary Table 60)**. For anthropometric measurements and lipid profile, confidence rating ranged from very low to high. For glycemic indices, confidence rating ranged from very low to moderate. For BP, confidence rating ranged from low to moderate. Downgrades from high for comparisons rated as moderate, low or very low were due to concerns in either one or more domain: within study bias, indirectness, imprecision, or heterogeneity.

## 4. Discussion

To the best of our knowledge, this is the first NMA assessing the existing evidence on various TRE interventions regarding human health. A total of 40 RCTs and 3259 participants were included in our analyses. Furthermore, in this article, we aimed to investigate the potential health outcomes related to meal timing. Therefore, we divided the interventions into three main categories: early, midday, and late TRE. With the growing number of RCTs combining exercise with TRE, further division of the studies was employed by including separate nodes for each of eTRE, mTRE, and lTRE + exercise, aiming to isolate the effects provided by exercise and to explore the potential extra benefits of combined interventions.

Overall, our analysis reveals a significant superiority of eTRE + EX over other interventions by achieving reductions in BW, BMI, FM, WC, FBG, FBI, HOMA-IR, TG, and LDL-C, along with an increase in HDL-C levels. Consequently, eTRE + EX gained the highest ranking on SUCRA, confirming its overall superiority. Nonetheless, the remaining TRE patterns, both alone and combined with exercise, as well as CR, showed comparable benefits regarding anthropometric measurements, and lipid profile, all of which were superior to control. This suggests that much of the observed effects are primarily driven by CR rather than meal timing.

However, in this NMA, eTRE significantly improved FBG, HbA1c, HOMA-IR, and SBP compared with control, whereas CR and other TRE variants did not. Although between-group comparisons were not statistically significant, the direction of effect consistently favored eTRE, highlighting the potential importance of meal timing for insulin sensitivity. These findings are consistent with a recent systematic review comparing TRE+CR versus CR alone, which reported improvements in insulin sensitivity in all studies testing eTRE + CR (N = 5) versus baseline, while CR alone did not show such improvements (71). The observed glucoregulatory effects with eTRE are mainly mediated by aligning food intake with the body’s circadian rhythms. This alignment seems to have an influence on such outcomes via hormonal and metabolic responses (e.g. insulin, cortisol, and melatonin), as peripheral tissues (i.e. muscle, liver) tend to be more sensitive during early times of the day due to their intrinsic circadian clocks (72). Thus, eating the larger proportion of calories during early time of the day may result in lower postprandial glucose and insulin levels. In addition, eTRE appears to reduce oxidative stress and preserve endothelial function, all of which can improve insulin resistance, improve beta-cell function, and lower BP (14,17,73). Concurrently, shifting the food intake into earlier times aligns with the upregulated genes affecting sodium excretion in the urine, resulting in lower SBP, while DBP seems to be minimally affected by food timing (74).

Since most the included RCTs in our analyses aimed to produce weight loss either by controlling the amount of caloric intake and/or eating window, there should be a detectable difference among TRE patterns and/or CR group in order to demonstrate notable role of meal timing. However, the role of meal timing in anthropometric measurements and lipid profile doesn’t seem to be as important as in glucose-related outcomes. Aligning with current findings, a recent meta-analysis that compared between different isocaloric IF approaches (e.g. ADF and 5:2) and CR didn’t find meaningful differences between the groups (75).

Interestingly, our subgroup analyses revealed greater weight loss when TRE groups are instructed to *ad libitum* food intake instead of prescribed energy intake, which are confirmed by a previous study (76) Aligning with these findings, a recent meta-analysis has compared the change of caloric intake between pre and post intervention in subjects who applied TRE without energy prescription (*ad libitum*) (77) The analysis revealed an unintended decrease in a total of approximately 400 calories/day in those who lost 3 kg or more, and a smaller reduction among participants lost less than 3 kg.

Although TRE with energy prescription was not as efficient as TRE with *ad libitum* food intake in the current analysis, such results may highlight potential psychological dimensions of the TRE protocol. For instance, it may indicate that the structured eating window of TRE leads to a natural reduction in calorie intake, without the mental effort or stress associated with calorie counting, which could potentially improve adherence and long-term sustainability (78,79) Earlier studies have similarly shown that TRE patterns often result in spontaneous calorie reduction, even when participants eat *ad libitum* (80). In contrast, when we applied subgroup analysis based on energy prescription on both interventional groups (ad libitum TRE vs energy deficit TRE), we also applied the same analysis on CR group in the same included studies. Surprisingly, even the CR group in studies where the TRE group was instructed to follow an ad libitum regimen appeared to perform better compared to their counterparts in the CR group in studies where the TRE group was directed to limit their energy intake (−2.73 vs -0.25 kg), suggesting the possibility of potential confounding factors. It is also possible that these findings may have arisen by chance rather than reflecting a true underlying effect. Therefore, the question whether *ad libitum* intake during TRE practice contributes to better outcomes is yet to be investigated.

Moreover, the evidence regarding the efficacy of meal timing across the meta-analyses is inconsistent. For instance, similar to our findings, differences between eTRE and lTRE regarding BW, FBG, and lipid profile didn’t reach statistical significance in overweight and obese subjects. However, there was a significant reduction in HOMA-IR and DBP in eTRE group compared to lTRE (81). On the other hand,(76) found a superiority of eTRE over lTRE on most of the outcomes which included BW, WC, FM, LBM, FBI, and HbA1c (76). Moreover, eTRE produced a significant reduction in FBG and HOMA-IR under weight maintenance conditions in subjects with excess BW (82). Notably, Egger’s test suggested the existence of publication bias. Further, these findings were disproven in recent studies by which the combination of eTRE and CR showed better results in BW, FM, and WC, but not blood biomarkers, compared to CR alone suggesting CR is the primary driver of metabolic benefits (83,84)

While eTRE is theoretically favoured due to its alignment with circadian biology, our analysis did not reveal significant differences between eTRE, mTRE, lTRE, and CR in all outcomes, which mostly can be explained to the overriding effects of energy deficit. Importantly, our findings suggest that the addition of exercise may act as a metabolic amplifier, enhancing the benefits of eTRE, possibly due to better alignment with circadian rhythms. Although the mechanisms of action are unclear, growth hormone (GH) and testosterone were shown to follow the biological clock, leading to metabolic variations. While GH is known to reach its maximum peak at the night, the highest levels of testosterone are detected at the morning. Subsequently, fat oxidation and insulin sensitivity are higher at the morning, which may affect muscle nutrients utilization (63). Despite this, these results should be interpreted with caution due to the limited number of studies with eTRE + EX intervention (n = 3), which one of them only measured BW. In addition, none of the studies directly compared between eTRE + EX and mTRE + EX, limiting the evidence to indirect comparisons.

### 4.1. Strengths, limitations, and future directions

Strengths of our current NMA include the largest comprehensive evidence synthesis about the comparative effect of meal timing from RCTs so far, capturing all recent publications. Further, unlike traditional pairwise meta-analysis, NMA allows for more precise estimates. The application of NMA methodology, which combined direct and indirect evidence, the extremely stringent inclusion and exclusion criteria, comprehensive literature retrieval, and a prior-registered system review protocol. In addition, we presented our results using the GRADE approach for grading recommendations. Further, this is the first NMA to distinguish between eTRE, mTRE, lTRE, such categorization provides a more nuanced understanding of how food timing may influence outcomes, thereby offering clearer insights.

Several limitations of this NMA should be acknowledged. There was very limited direct evidence for certain interventions such as lTRE, eTRE combined with exercise, and mTRE combined with exercise, which limits certainty in comparative rankings and highlights the need for high-quality RCTs to examine these strategies. No published studies examined lTRE combined with exercise, further restricting evidence in this area. We were unable to isolate the effect of fasting duration, given the variability in eating windows and dietary intake across studies, which likely influences energy consumption, circadian alignment, and metabolic outcomes independently. Additionally, the small number of exercise-based trials precluded separating the effects of exercise type, despite evidence that resistance and aerobic training impact body composition and metabolic indices differently. Further, the superior results based on shorter duration have been suggested to be mediated by better adherence compared to longer duration. However, adherence was not measured in our study. Finally, only local inconsistency was examined; incorporating both local and global inconsistency in future analyses would strengthen the validity and robustness of these conclusions.

## 5. Conclusion

This NMA provides a comprehensive comparison of different TRE patterns, including eTRE, mTRE, and lTRE, with or without the addition of exercise, CR, and control. While our analysis did not detect statistically significant differences between TRE patterns and CR, the consistent SUCRA rankings in favor of eTRE (particularly with exercise) suggest that meal timing may play an important role. However, these findings were limited by very low to high for anthropometric indices and lipid profile, and very low to moderate certainty of evidence for glycemic indices Moreover, owing to the limited number of RCTs testing TRE + EX, these results should be interpreted with cautious. To strengthen the validity of findings, upcoming RCTs should rigorously assess baseline and post-intervention eating windows in both TRE and CR/control groups and avoid unintentional fasting or habitual circadian alignment (e.g., eating within an 8 a.m.–8 p.m. window) in CR arms to ensure a clear distinction between interventions. Accordingly, future studies are needed to draw robust conclusions on the combined health benefits of meal timing, with or without exercise.

## Authors’ contributions

M.H. and W.S. carried out the concept and design. M.H. wrote the first draft of this study. Y.R. and A.F. searched databases, and screened articles. M.H., M.T., and W.S. performed the acquisition, data extraction, analysis, and interpretation of data. Y.R., K.A.V., and A.F. critically revised the manuscript. All authors approved the final version of the manuscript.

## Data availability

All data produced in the present work are contained in the manuscript

## Data Availability

All data produced in the present work are contained in the manuscript

## References

1. Di Francesco A, Di Germanio C, Bernier M, de Cabo R. A time to fast [Internet]. Available from: http://science.sciencemag.org/

2. Charlot A, Hutt F, Sabatier E, Zoll J. Beneficial effects of early time-restricted feeding on metabolic diseases: Importance of aligning food habits with the circadian clock. Vol. 13, Nutrients. MDPI AG; 2021.

3. Wehrens SMT, Christou S, Isherwood C, Middleton B, Gibbs MA, Archer SN, et al. Meal Timing Regulates the Human Circadian System. Current Biology. 2017 Jun 19;27(12):1768–1775.e3.

4. Carrasco-Benso MP, Rivero-Gutierrez B, Lopez-Minguez J, Anzola A, Diez-Noguera A, Madrid JA, et al. Human adipose tissue expresses intrinsic circadian rhythm in insulin sensitivity. FASEB Journal. 2016 Sep 1;30(9):3117–23.

5. Fatima N, Rana S. Metabolic implications of circadian disruption. Vol. 472, Pflugers Archiv European Journal of Physiology. Springer; 2020. p. 513–26.

6. Poggiogalle E, Jamshed H, Peterson CM. Circadian regulation of glucose, lipid, and energy metabolism in humans. Metabolism. 2018 Jul 1;84:11–27.

7. Flanagan A, Bechtold DA, Pot GK, Johnston JD. Chrono-nutrition: From molecular and neuronal mechanisms to human epidemiology and timed feeding patterns. Vol. 157, Journal of Neurochemistry. Blackwell Publishing Ltd; 2021. p. 53–72.

8. Kant AK, Graubard BI. Within-person comparison of eating behaviors, time of eating, and dietary intake on days with and without breakfast: NHANES 2005-20101-3. Vol. 102, American Journal of Clinical Nutrition. American Society for Nutrition; 2015. p. 661–70.

9. Zeballos E, Todd JE. The effects of skipping a meal on daily energy intake and diet quality. Public Health Nutr. 2020 Dec 1;23(18):3346–55.

10. Liu HY, Eso AA, Cook N, O’Neill HM, Albarqouni L. Meal Timing and Anthropometric and Metabolic Outcomes: A Systematic Review and Meta-Analysis. JAMA Netw Open. 2024 Nov 1;7(11):e2442163.

11. He M, Wang J, Liang Q, Li M, Guo H, Wang Y, et al. Time-restricted eating with or without low-carbohydrate diet reduces visceral fat and improves metabolic syndrome: A randomized trial. Cell Rep Med. 2022 Oct 18;3(10).

12. Zhang L min, Liu Z, Wang J qi, Li R qiang, Ren J yi, Gao X, et al. Randomized controlled trial for time-restricted eating in overweight and obese young adults. iScience. 2022 Sep 16;25(9).

13. Peters B, Schwarz J, Schuppelius B, Ottawa A, Koppold DA, Weber D, et al. Intended isocaloric time-restricted eating shifts circadian clocks but does not improve cardiometabolic health in women with overweight. Sci Transl Med. 2025;17(822):eadv6787. doi:10.1126/scitranslmed.adv6787

14. Xie Z, Sun Y, Ye Y, Hu D, Zhang H, He Z, et al. Randomized controlled trial for time-restricted eating in healthy volunteers without obesity. Nat Commun. 2022 Dec 1;13(1).

15. Dote-Montero M, Clavero-Jimeno A, Merchán-Ramírez E, Oses M, Echarte J, Camacho-Cardenosa A, et al. Effects of early, late and self-selected time-restricted eating on visceral adipose tissue and cardiometabolic health in participants with overweight or obesity: a randomized controlled trial. Nat Med. 2025 Feb 1;31(2):524–33.

16. Tsitsou S, Bali T, Adamantou M, Saridaki A, Poulia KA, Karagiannakis DS, et al. Effects of a 12-Week Mediterranean-Type Time-Restricted Feeding Protocol in Patients With Metabolic Dysfunction-Associated Steatotic Liver Disease: A Randomised Controlled Trial—The ‘CHRONO-NAFLD Project.’ Aliment Pharmacol Ther. 2025 Apr 1;61(8):1290–309.

17. Sutton EF, Beyl R, Early KS, Cefalu WT, Ravussin E, Peterson CM. Early Time-Restricted Feeding Improves Insulin Sensitivity, Blood Pressure, and Oxidative Stress Even without Weight Loss in Men with Prediabetes. Cell Metab. 2018 Jun 5;27(6):1212–1221.e3.

18. Jamshed H, Steger FL, Bryan DR, Richman JS, Warriner AH, Hanick CJ, et al. Effectiveness of Early Time-Restricted Eating for Weight Loss, Fat Loss, and Cardiometabolic Health in Adults with Obesity: A Randomized Clinical Trial. JAMA Intern Med. 2022 Sep 1;182(9):953–62.

19. Queiroz JDN, MacEdo RCO, Dos Santos GC, Munhoz SV, MacHado CLF, De Menezes RL, et al. Cardiometabolic effects of early v. delayed time-restricted eating plus energetic restriction in adults with overweight and obesity: an exploratory randomised clinical trial. British Journal of Nutrition. 2023 Feb 28;129(4):637–49.

20. Črešnovar T, Habe B, Mohorko N, Kenig S, Jenko Pražnikar Z, Petelin A. Early time-restricted eating with energy restriction has a better effect on body fat mass, diastolic blood pressure, metabolic age and fasting glucose compared to late time-restricted eating with energy restriction and/or energy restriction alone: A 3-month randomized clinical trial. Clinical Nutrition. 2025 Jun 1;49:57–68.

21. Liu H, Chen S, Ji H, Dai Z. Effects of time-restricted feeding and walking exercise on the physical health of female college students with hidden obesity: a randomized trial. Front Public Health. 2023;11.

22. Ameur R, Maaloul R, Tagougui S, Neffati F, Kacem FH, Najjar MF, et al. Unlocking the power of synergy: High-intensity functional training and early time-restricted eating for transformative changes in body composition and cardiometabolic health in inactive women with obesity. PLoS One. 2024 May 1;19(5 May).

23. Dai Z, Wan K, Miyashita M, Ho RS tak, Zheng C, Poon ET chun, et al. The Effect of Time-Restricted Eating Combined with Exercise on Body Composition and Metabolic Health: A Systematic Review and Meta-Analysis. Vol. 15, Advances in Nutrition. Elsevier B.V.; 2024.

24. Cui T, Sun Y, Ye W, Liu Y, Korivi M. Efficacy of time restricted eating and resistance training on body composition and mood profiles among young adults with overweight/obesity: a randomized controlled trial. J Int Soc Sports Nutr. 2025;22(1).

25. Hays HM, Sefidmooye Azar P, Kang M, Tinsley GM, Wijayatunga NN. Effects of time-restricted eating with exercise on body composition in adults: a systematic review and meta-analysis: Clinical Research. Vol. 49, International Journal of Obesity. Springer Nature; 2025. p. 755–65.

26. Moher D, Liberati A, Tetzlaff J, Altman DG. Preferred reporting items for systematic reviews and meta-analyses: The PRISMA statement. Vol. 339, BMJ (Online). 2009. p. 332–6.

27. Higgins JPT, Altman DG, Gøtzsche PC, Jüni P, Moher D, Oxman AD, et al. The Cochrane Collaboration’s tool for assessing risk of bias in randomised trials. BMJ (Online). 2011 Oct 29;343(7829).

28. Nikolakopoulou A, Higgins JPT, Papakonstantinou T, Chaimani A, Giovane C Del, Egger M, et al. Cinema: An approach for assessing confidence in the results of a network meta-analysis. PLoS Med. 2020 Apr 2;17(4).

29. Cipriani A, Higgins JPT, Geddes JR, Salanti G. Conceptual and Technical Challenges in Network Meta-analysis [Internet]. 2013. Available from: www.annals.org

30. Jansen JP, Naci H. Is network meta-analysis as valid as standard pairwise meta-analysis? It all depends on the distribution of effect modifiers. BMC Med. 2013 Jul 4;11(1).

31. Papakonstantinou T, Nikolakopoulou A, Higgins JPT, Egger M, Salanti G. CINeMA: Software for semiautomated assessment of the confidence in the results of network meta-analysis. Campbell Systematic Reviews. 2020 Mar 1;16(1).

32. Semnani-Azad Z, Khan TA, Chiavaroli L, Chen V, Bhatt HA, Chen A, et al. Intermittent fasting strategies and their effects on body weight and other cardiometabolic risk factors: Systematic review and network meta-analysis of randomised clinical trials. BMJ. 2025;

33. Wu X, Ding Y, Cao Q, Huang J, Xu X, Jiang Y, et al. Comparison of Different Intermittent Fasting Patterns or Different Extents of Calorie Restriction for Weight Loss and Metabolic Improvement in Adults: A Systematic Review and Network Meta-Analysis of Randomized Controlled Trials. Nutr Rev. 2025 May 14;

34. Owen RK, Bradbury N, Xin Y, Cooper N, Sutton A. MetaInsight: An interactive web-based tool for analyzing, interrogating, and visualizing network meta-analyses using R-shiny and netmeta. Res Synth Methods. 2019 Dec 1;10(4):569–81.

35. Brignardello-Petersen R, Murad MH, Walter SD, McLeod S, Carrasco-Labra A, Rochwerg B, et al. GRADE approach to rate the certainty from a network meta-analysis: avoiding spurious judgments of imprecision in sparse networks. J Clin Epidemiol. 2019 Jan 1;105:60–7.

36. Higgins JP, Li T, Deeks JJ. Chapter 6: Choosing effect measures and computing estimates of effect 6.1 Types of data and effect measures 6.1.1 Types of data [Internet]. 2024. Available from: www.documentation.cochrane.org/revman-kb/statistical-methods-210600101.html

37. Hozo SP, Djulbegovic B, Hozo I. Estimating the mean and variance from the median, range, and the size of a sample. BMC Med Res Methodol. 2005 Apr 20;5.

38. Elbourne DR, Altman DG, Higgins JP, Curtin F, Worthington H V, Vail A. Meta-analyses involving cross-over trials: methodological issues. Vol. 31, International Journal of Epidemiology. 2002.

39. Dersimonian R, Laird N. Meta-Analysis in Clinical Trials*.

40. Turner RM, Davey J, Clarke MJ, Thompson SG, Higgins JP. Predicting the extent of heterogeneity in meta-analysis, using empirical data from the Cochrane Database of Systematic Reviews. Int J Epidemiol. 2012 Jun;41(3):818–27.

41. van Valkenhoef G, Dias S, Ades AE, Welton NJ. Automated generation of node-splitting models for assessment of inconsistency in network meta-analysis. Res Synth Methods. 2016 Mar 1;7(1):80–93.

42. Liu D, Huang Y, Huang C, Yang S, Wei X, Zhang P, et al. Calorie Restriction with or without Time-Restricted Eating in Weight Loss. New England Journal of Medicine. 2022 Apr 21;386(16):1495–504.

43. Deng Y, Liu X, Sun Y, Zhou L, Li Q, Lei Z, et al. Effects of time-restricted eating on intrahepatic fat and metabolic health among patients with nonalcoholic fatty liver disease. Obesity. 2024 Mar 1;32(3):494–505.

44. Che T, Yan C, Tian D, Zhang X, Liu X, Wu Z. Time-restricted feeding improves blood glucose and insulin sensitivity in overweight patients with type 2 diabetes: a randomised controlled trial. Nutr Metab (Lond). 2021 Dec 1;18(1).

45. Wei X, Lin B, Huang Y, Yang S, Huang C, Shi L, et al. Effects of Time-Restricted Eating on Nonalcoholic Fatty Liver Disease: The TREATY-FLD Randomized Clinical Trial. JAMA Netw Open. 2023 Mar 17;6(3):E233513.

46. Zhou X, Lin X, Yu J, Yang Y, Muzammel H, Amissi S, et al. Effects of DASH diet with or without time-restricted eating in the management of stage 1 primary hypertension: a randomized controlled trial. Nutr J. 2024 Dec 1;23(1).

47. Kotarsky CJ, Johnson NR, Mahoney SJ, Mitchell SL, Schimek RL, Stastny SN, et al. Time-restricted eating and concurrent exercise training reduces fat mass and increases lean mass in overweight and obese adults. Physiol Rep. 2021 May 1;9(10).

48. Gabel K, Hoddy KK, Haggerty N, Song J, Kroeger CM, Trepanowski JF, et al. Effects of 8-hour time restricted feeding on body weight and metabolic disease risk factors in obese adults: A pilot study. Nutr Healthy Aging. 2018;4(4):345–53.

49. Cienfuegos S, Gabel K, Kalam F, Ezpeleta M, Wiseman E, Pavlou V, et al. Effects of 4- and 6-h Time-Restricted Feeding on Weight and Cardiometabolic Health: A Randomized Controlled Trial in Adults with Obesity. Cell Metab. 2020 Sep 1;32(3):366–378.e3.

50. Lin S, Cienfuegos S, Ezpeleta M, Gabel K, Pavlou V, Mulas A, et al. Time-Restricted Eating Without Calorie Counting for Weight Loss in a Racially Diverse Population. Ann Intern Med. 2023 Jul 1;176(7):885–95.

51. Pavlou V, Cienfuegos S, Lin S, Ezpeleta M, Ready K, Corapi S, et al. Effect of Time-Restricted Eating on Weight Loss in Adults with Type 2 Diabetes: A Randomized Clinical Trial. JAMA Netw Open. 2023 Oct 27;6(10):E2339337.

52. Lowe DA, Wu N, Rohdin-Bibby L, Moore AH, Kelly N, Liu YE, et al. Effects of Time-Restricted Eating on Weight Loss and Other Metabolic Parameters in Women and Men with Overweight and Obesity: The TREAT Randomized Clinical Trial. JAMA Intern Med. 2020 Nov 1;180(11):1491–9.

53. Domaszewski P, Konieczny M, Dybek T, Łukaniszyn-Domaszewska K, Anton S, Sadowska-Krępa E, et al. Comparison of the effects of six-week time-restricted eating on weight loss, body composition, and visceral fat in overweight older men and women. Exp Gerontol. 2023 Apr 1;174.

54. Domaszewski P, Konieczny M, Pakosz P, Łukaniszyn-Domaszewska K, Mikuláková W, Sadowska-Krępa E, et al. Effect of a six-week times restricted eating intervention on the body composition in early elderly men with overweight. Sci Rep. 2022 Dec 1;12(1).

55. Domaszewski P, Konieczny M, Pakosz P, Baczkowicz D, Sadowska-Krępa E. Effect of a six-week intermittent fasting intervention program on the composition of the human body in women over 60 years of age. Int J Environ Res Public Health. 2020 Jun 1;17(11):1–9.

56. Miladi S, Driss T, Ameur R, Miladi SC, Miladi SJ, Najjar MF, et al. Effectiveness of Early Versus Late Time-Restricted Eating Combined with Physical Activity in Overweight or Obese Women. Nutrients . 2025 Jan 1;17(1).

57. Irani H, Abiri B, Khodami B, Yari Z, Lafzi Ghazi M, Hosseinzadeh N, et al. Effect of time restricted feeding on anthropometric measures, eating behavior, stress, serum levels of BDNF and LBP in overweight/obese women with food addiction: a randomized clinical trial. Nutr Neurosci. 2024;27(6):577–89.

58. Talebi S, Shab-Bidar S, Moini A, Mohammadi H, Djafarian K. The effects of time-restricted eating alone or in combination with probiotic supplementation in comparison with a calorie-restricted diet on endocrine and metabolic profiles in women with polycystic ovary syndrome: A randomized clinical trial. Diabetes Obes Metab. 2024 Oct 1;26(10):4468–79.

59. Mengi Çelik Ö, Köksal E, Aktürk M. Time-restricted eating (16/8) and energy-restricted diet: effects on diet quality, body composition and biochemical parameters in healthy overweight females. BMC Nutr. 2023 Dec 1;9(1).

60. Erdem NZ, Bayraktaroğlu E, Samancı RA, Geçgil-Demir E, Tarakçı NG, Mert-Biberoğlu F. The effect of intermittent fasting diets on body weight and composition. Clin Nutr ESPEN. 2022 Oct 1;51:207–14.

61. Feehan J, Mack A, Tuck C, Tchongue J, Holt DQ, Sievert W, et al. Time-Restricted Fasting Improves Liver Steatosis in Non-Alcoholic Fatty Liver Disease—A Single Blinded Crossover Trial. Nutrients. 2023 Dec 1;15(23).

62. Parr EB, Radford BE, Hall RC, Steventon-Lorenzen N, Flint SA, Siviour Z, et al. Comparing the effects of time-restricted eating on glycaemic control in people with type 2 diabetes with standard dietetic practice: A randomised controlled trial. Diabetes Res Clin Pract. 2024 Nov 1;217.

63. Yu Z, Ueda T. Early Time-Restricted Eating Improves Weight Loss While Preserving Muscle: An 8-Week Trial in Young Women. Nutrients . 2025 Mar 1;17(6).

64. Kahleova H, Belinova L, Malinska H, Oliyarnyk O, Trnovska J, Skop V, et al. Eating two larger meals a day (breakfast and lunch) is more effective than six smaller meals in a reduced-energy regimen for patients with type 2 diabetes: A randomised crossover study. Diabetologia. 2014;57(8):1552–60.

65. Lin YJ, Wang YT, Chan LC, Chu NF. Effect of time-restricted feeding on body composition and cardio-metabolic risk in middle-aged women in Taiwan. Nutrition. 2022 Jan 1;93.

66. Kramer CK, Zinman B, Feig DS, Retnakaran R. Effect of Time-Restricted Eating on β-Cell Function in Adults With Type 2 Diabetes. Journal of Clinical Endocrinology and Metabolism. 2025 Jun 1;110(6):e2045–53.

67. Mena-Hernández DR, Jiménez-Domínguez G, Méndez JD, Olvera-Hernández V, Martínez-López MC, Guzmán-Priego CG, et al. Effect of Early Time-Restricted Eating on Metabolic Markers and Body Composition in Individuals with Overweight or Obesity. Nutrients. 2024 Jul 1;16(14).

68. Quist JS, Pedersen HE, Jensen MM, Clemmensen KKB, Bjerre N, Ekblond TS, et al. Effects of 3 months of 10-h per-day time-restricted eating and 3 months of follow-up on bodyweight and cardiometabolic health in Danish individuals at high risk of type 2 diabetes: the RESET single-centre, parallel, superiority, open-label, randomised controlled trial. Lancet Healthy Longev. 2024 May 1;5(5):e314–25.

69. Rizvi ZA, Saleem J, Zeb I, Shahzad R, Kayani JA, Faryal J, et al. Effects of intermittent fasting on body composition, clinical health markers and memory status in the adult population: a single-blind randomised controlled trial. Nutr J. 2024 Dec 1;23(1).

70. Sampieri A, Paoli A, Spinello G, Santinello E, Moro T. Impact of daily fasting duration on body composition and cardiometabolic risk factors during a time-restricted eating protocol: a randomized controlled trial. J Transl Med. 2024 Dec 1;22(1).

71. Ezzati A, McLaren C, Bohlman C, Tamargo JA, Lin Y, Anton SD. Does time-restricted eating add benefits to calorie restriction? A systematic review. Vol. 32, Obesity. John Wiley and Sons Inc; 2024. p. 640–54.

72. Shkorfu W, Fadel A, Hamsho M, Ranneh Y, Shahbaz HM. Intermittent Fasting and Hormonal Regulation: Pathways to Improved Metabolic Health. Vol. 13, Food Science and Nutrition. John Wiley and Sons Inc; 2025.

73. Ezzati A, Tamargo JA, Golberg L, Haub MD, Anton SD. The Effects of Time-Restricted Eating on Inflammation and Oxidative Stress in Overweight Older Adults: A Pilot Study. Nutrients . 2025 Jan 1;17(2).

74. Varady KA, Cienfuegos S, Ezpeleta M, Gabel K. Clinical application of intermittent fasting for weight loss: progress and future directions. Vol. 18, Nature Reviews Endocrinology. Nature Research; 2022. p. 309–21.

75. Hamsho M, Shkorfu W, Ranneh Y, Fadel A. Is isocaloric intermittent fasting superior to calorie restriction? A systematic review and meta-analysis of RCTs. Nutrition, Metabolism and Cardiovascular Diseases. 2025 Mar 1;35(3).

76. Chang Y, Du T, Zhuang X, Ma G. Time-restricted eating improves health because of energy deficit and circadian rhythm: A systematic review and meta-analysis. iScience. 2024 Feb 16;27(2).

77. Quang DT, Di Khanh N, Cu L Le, Thi Hoa HN, Quynh CVT, Ngoc QP, et al. Partially unraveling mechanistic underpinning and weight loss effects of time-restricted eating across diverse adult populations: A systematic review and meta-analyses of prospective studies. PLoS One. 2025 Jan 1;20(1).

78. Ezpeleta M, Cienfuegos S, Lin S, Pavlou V, Gabel K, Tussing-Humphreys L, et al. Time-restricted eating: Watching the clock to treat obesity. Vol. 36, Cell Metabolism. Cell Press; 2024. p. 301–14.

79. Hamsho M, Shkorfu W, Ranneh Y, Fadel A. Nourishing the evidence: exposing bias and filling gaps in isocaloric intermittent fasting research—An opinion [Internet]. Vol. 14, Nutrients. MDPI; 2025 [cited 2025 Sep 17]. Available from: https://www.frontiersin.org/journals/nutrition/articles/10.3389/fnut.2025.1563017/full

80. Petridi F, Geurts JMW, Nyakayiru J, Schaafsma A, Schaafsma D, Meex RCR, et al. Effects of Early and Late Time-Restricted Feeding on Parameters of Metabolic Health: An Explorative Literature Assessment. Vol. 16, Nutrients. 2024.

81. Liu J, Yi P, Liu F. The Effect of Early Time-Restricted Eating vs Later Time-Restricted Eating on Weight Loss and Metabolic Health. Journal of Clinical Endocrinology and Metabolism. 2023 Jul 1;108(7):1824–34.

82. Pureza IR de OM, Macena M de L, da Silva Junior AE, Praxedes DRS, Vasconcelos LGL, Bueno NB. Effect of early time-restricted feeding on the metabolic profile of adults with excess weight: A systematic review with meta-analysis. Clinical Nutrition. 2021 Apr 1;40(4):1788–99.

83. Črešnovar T, Habe B, Jenko Pražnikar Z, Petelin A. Effectiveness of Time-Restricted Eating with Caloric Restriction vs. Caloric Restriction for Weight Loss and Health: Meta-Analysis. Vol. 15, Nutrients. Multidisciplinary Digital Publishing Institute (MDPI); 2023.

84. Sun JC, Tan ZT, He CJ, Hu HL, Zhai CL, Qian G. Time-restricted eating with calorie restriction on weight loss and cardiometabolic risk: a systematic review and meta-analysis. Vol. 77, European Journal of Clinical Nutrition. Springer Nature; 2023. p. 1014–25.

