## Supplementary material for "How Does Early, Midday, and Late Time-Restricted Eating Impact Anthropometry and Cardiometabolic Health? A Systematic Review and Network Meta-Analysis of RCTs": Supplemenary materials

**What Does Evidence Reveal About Meal Timing, Metabolic Health, and Anthropometry? A Systematic Review and Network Meta-Analysis of RCTs.**

**Supplementary materials**

**Contents**

Tables	2-41

Figures...................................................................................................................................46-70

**Supplementary Table 1:** Included interventions and their definition

| **Intervention** | **Definition** |
| --- | --- |
| Early time-restricted eating (eTRE) | A time-restricted eating pattern characterized by a daily fasting duration of ≥14 hours, with the first caloric intake occurring before 10:00 AM. |
| Middle day time-restricted eating (mTRE) | A time-restricted eating pattern characterized by a daily fasting duration of ≥14 hours, with the first caloric intake occurring between 10:01 AM to 12:00 PM. |
| Late time-restricted eating (lTRE) | A time-restricted eating pattern characterized by a daily fasting duration of ≥14 hours, with the first caloric intake occurring after 12:00 PM. |
| eTRE + exercise (eTRE + EX) | Similar definition to eTRE combined with any type of exercise |
| mTRE + exercise (mTRE + EX) | Similar definition to mTRE combined with any type of exercise |
| Caloric restriction (CR) | Calorie restriction without time restriction |
| Control | Control conditions without prescribed eating or fasting windows (e.g., ad libitum intake, usual care, or no intervention), |

**Supplementary Table 2:** Minimally important difference (MID) threshold for each outcome.

| **Variable** | **MID Threshold** | **Rational and reference** |
| --- | --- | --- |
| Body weight | 1.0 kg | 1 kg is an MID reported by Semnani-Azad et al. (2025) (1). |
| BMI | 0.2 kg/m^2^ | Approximately equivalent to the MID for body weight of 1 kg; adjusted to 0.2 kg/m² for consistency (1). |
| Fat (%) | 2% | Absolute reduction based on 5-7% weight loss being clinically meaningful (1, 2). |
| Fat (kg) | 1 kg | Absolute reduction based on approximately 2% as clinically meaningful, consistent with a 1 kg body weight reduction and approximately 2% body fat decrease (2, 3). |
| Lean body mass (kg) | 1 kg | Based on evidence from clinical trials in overweight and obese adults showing typical lean body mass changes of approximately 1.0 kg during weight loss interventions (4). |
| Waist circumference (cm) | 2 cm | Approximately equivalent to the MID for body weight of 1 kg; adjusted to 2 cm for consistency (1, 2). |
| Hip circumference (cm) | 1 cm | No established minimal important difference exists for hip circumference. Given that hip circumference generally shows smaller absolute changes than waist circumference in weight-loss interventions, a conservative threshold of approximately 1 cm was used to aid interpretation, corresponding to half of the threshold applied for waist circumference. |
| Fasting blood glucose (mg/dl) | 9 mg/dl | Approximately equivalent to the MID for A1C of 0.3% (1, 5). |
| Fasting blood insulin (mIU/L) | 0.72 mIU/L | Approximately equivalent to the MID for fasting glucose of 0.5 mmol/L (1). |
| HOMA-IR | 1 | Proportional reduction to fasting glucose (1). |
| HbA1c (%) | 0.3% | Threshold identified as clinically relevant by EMA and FDA (1, 6, 7, 8). |
| Triglyceride (mmol/L) | 0.1 mmol/L | 0.1 mmol/L represents the minimum reduction used to support health claims (9, 10) and CCS guidelines (11). |
| Total cholesterol (mg/dl) | 3.86 mg/dl | 0.1 mmol/L represents the minimum reduction used to support health claims (9, 10) and CCS guidelines (11). |
| Low density lipoprotein (mg/dl) | 3.86 mg/dl | 0.1 mmol/L represents the minimum reduction used to support health claims (9, 10) and CCS guidelines (11). |
| High density lipoprotein (mg/dl) | 3.86 mg/dl | 0.1 mmol/L represents the minimum reduction used to support health claims (9, 10) and CCS guidelines (11). |
| Systolic blood pressure (mmHg) | 4 mmHg | A MID of 4 mmHg was used for systolic and diastolic blood pressure, based on FDA clinical evaluation discussions indicating that reductions of 2–4 mmHg are clinically meaningful; the upper bound was chosen as a threshold (12) |
| Diastolic blood pressure (mmHg) | 4 mmHg |  |

**Footnotes:** MID = minimal important difference, BMI = body mass index, HOMA-IR = homeostatic model assessment of insulin resistance, EMA = European medicines agency, FDA = food and drug administration, CSS = Canadian cardiovascular society.

1 Semnani-Azad, Z., Khan, T. A., Chiavaroli, L., Chen, V., Bhatt, H. A., Chen, A., Chiang, N., Oguntala, J., Kabisch, S., Lau, D. C., Wharton, S., Sharma, A. M., Harris, L., Leiter, L. A., Hill, J. O., Hu, F. B., Lean, M. E., Kahleová, H., Rahelic, D., . . . Sievenpiper, J. L. (2025). Intermittent fasting strategies and their effects on body weight and other cardiometabolic risk factors: systematic review and network meta-analysis of randomised clinical trials. *BMJ*, *389*, e082007. https://doi.org/10.1136/bmj-2024-082007
2 Jayedi, A., Soltani, S., Emadi, A., Zargar, M., & Najafi, A. (2024). Aerobic exercise and weight loss in adults. *JAMA Network Open*, *7*(12), e2452185. https://doi.org/10.1001/jamanetworkopen.2024.52185
3 Wu, X., Ding, Y., Cao, Q., Huang, J., Xu, X., Jiang, Y., Xu, Y., Lu, J., Xu, M., Wang, T., Zhao, Z., Wang, W., Ning, G., Bi, Y., & Li, M. (2025). Comparison of different intermittent fasting patterns or different extents of calorie restriction for weight loss and metabolic improvement in adults: a Systematic Review and Network Meta-Analysis of Randomized Controlled Trials. *Nutrition Reviews*. https://doi.org/10.1093/nutrit/nuaf056
4 Binmahfoz, A., Johnston, L., Dunning, E., Gray, C. M., & Gray, S. R. (2025). The effects of a home-based resistance training programme on body composition and muscle function during weight loss in people living with overweight or obesity: a randomised controlled pilot trial. *Nutrition & Metabolism*, *22*(1), 90. https://doi.org/10.1186/s12986-025-00986-1
5 Nathan DM, Kuenen J, Borg R, Zheng H, Schoenfeld D, Heine RJ. Translating the A1C assay into estimated average glucose values. Diabetes Care [Internet]. 2008 Jun 8;31(8):1473–8. Available from: <https://doi.org/10.2337/dc08-0545>
6 European Medicines Agency. Guideline on clinical investigation of medicinal products in the treatment or prevention of diabetes mellitus. CPMP/EWP/1080/00 Rev. 1. 2018. https://www.ema.europa.eu/en/documents/scientific-guideline/draft-guideline-clinical-investigation-medicinal-products-treatment-prevention-diabetesmellitus_en.pdf
7 Food and Drug Administration. Guidance for Industry on Diabetes Mellitus-Evaluating Cardiovascular Risk in New Antidiabetic Therapies to Treat Type 2 Diabetes; Availability. 2008. <https://www.federalregister.gov/documents/2008/12/19/E8-30086/guidance-for-industry-on-diabetes-mellitus-evaluatingcardiovascular-risk-in-new-antidiabetic>
8 U.S. Food & Drug Administration. Diabetes Mellitus: Efficacy Endpoints for Clinical Trials Investigating Antidiabetic Drugs and Biological Products Guidance for Industry. Draft Guidance. 2023. <https://www.fda.gov/media/168475/download>
9 Food Directorate, Health Products and Food Branch, Health Canada. Summary of Health Canada's assessment of a health claim about soy protein and cholesterol lowering. Ottawa: Bureau of Nutritional Sciences. March 2015. https://www.canada.ca/en/health-canada/services/food-nutrition/food-labelling/healthclaims/assessments/summary-assessment-health-claim-about-protein-cholesterol-lowering.html
10 Food Directorate, Health Products and Food Branch, Health Canada. Oat products and blood cholesterol lowering. Ottawa: Bureau of Nutritional Sciences. 2010.
11 Anderson TJ, Grégoire J, Pearson GJ, et al. 2016 Canadian Cardiovascular Society Guidelines for the Management of Dyslipidemia for the Prevention of Cardiovascular Disease in the Adult. Can J Cardiol. 2016;32(11):1263-82.
12 Fda. (2023). FDA Executive Summary Circulatory System Devices Panel Meeting Clinical Evaluation of Anti-Hypertensive Devices General Issues Panel-Clinical Evaluation of Anti-Hypertensive Devices.

**Subgroups analyses:**

**Supplementary Table 3:** Subgroup analysis presenting mean differences with 95% CI in BW (kg) between different interventions, stratified by energy prescription: energy prescribed (bottom) and *ad libitum* (top).

| Control | -2.73 (-3.8, -1.66) | -2.23 (-2.81, -1.65) | -3.26 (-4.88, -1.7) | -2.19 (-3.2, -1.19) | -1.8 (-2.38, -1.25) | -1.78 (-3.49, -0.12) |
| --- | --- | --- | --- | --- | --- | --- |
| -0.25 (-1.5, 1.21) | CR | 0.49 (-0.56, 1.58) | -0.53 (-2.42, 1.31) | 0.54 (-0.7, 1.77) | 0.92 (-0.15, 1.98) | 0.95 (-1.04, 2.88) |
| -1.3 (-2.31, -0.06) | -1.06 (-1.91, -0.18) | eTRE | -1.03 (-2.64, 0.52) | 0.05 (-0.96, 1.04) | 0.43 (-0.26, 1.07) | 0.45 (-1.33, 2.18) |
| / | / | / | eTRE + EX | 1.07 (-0.72, 2.93) | 1.46 (-0.18, 3.12) | 1.48 (-0.81, 3.78) |
| / | / | / | / | lTRE | 0.39 (-0.71, 1.45) | 0.4 (-1.56, 2.33) |
| -0.89 (-2.17, 0.54) | -0.65 (-1.53, 0.21) | 0.41 (-0.55, 1.34) | / | / | mTRE | 0.02 (-1.68, 1.71) |
| / | / | / | / | / | / | mTRE + EX |

**Footnotes:** CR = calorie restriction, eTRE = early time-restrcited eating, mTRE = midday time-restricted eating, lTRE = late time-restricted eating, eTRE + EX = early time-restricted eating + exercise, mTRE + EX = midday time-restricted eating + exercise.

**Supplementary Table 4**: Subgroup analysis presenting mean differences with 95% CI in BW (kg) between different interventions, stratified by intervention duration: 11 weeks ≤ (bottom) and 10 weeks ≥ (top).

| Control | -1.81 (-3.01, -0.58) | -2.02 (-2.74, -1.21) | -2.78 (-4.58, -0.92) | -1.3 (-3.21, 0.72) | -1.67 (-2.38, -0.97) | -1.72 (-3.48, -0.04) |
| --- | --- | --- | --- | --- | --- | --- |
| -1.84 (-3.12, -0.64) | CR | -0.2 (-1.35, 0.97) | -0.98 (-3.07, 1.17) | 0.51 (-1.62, 2.69) | 0.14 (-0.96, 1.18) | 0.08 (-1.99, 2.06) |
| -2.27 (-3.3, -1.33) | -0.43 (-1.55, 0.7) | eTRE | -0.77 (-2.61, 1.06) | 0.72 (-1.09, 2.54) | 0.34 (-0.45, 1.04) | 0.28 (-1.61, 2.05) |
| -4.87 (-8.76, -1.06) | -3.03 (-6.94, 0.87) | -2.6 (-6.35, 1.13) | eTRE + EX | 1.49 (-1.08, 4.09) | 1.11 (-0.82, 2.97) | 1.05 (-1.5, 3.48) |
| -2.2 (-3.67, -0.74) | -0.36 (-1.92, 1.27) | 0.07 (-1.44, 1.65) | 2.68 (-1.31, 6.74) | lTRE | -0.37 (-2.39, 1.52) | -0.43 (-3.12, 2.05) |
| -1.97 (-3.06, -0.95) | -0.14 (-1.42, 1.17) | 0.29 (-0.88, 1.5) | 2.89 (-0.99, 6.83) | 0.23 (-1.48, 1.87) | mTRE | -0.05 (-1.81, 1.65) |
| / | / | / | / | / | / | mTRE + EX |

**Footnotes:** CR = calorie restriction, eTRE = early time-restrcited eating, mTRE = midday time-restricted eating, lTRE = late time-restricted eating, eTRE + EX = early time-restricted eating + exercise, mTRE + EX = midday time-restricted eating + exercise.

**Supplementary Table 5:** Subgroup analysis presenting mean differences with 95% CI in BMI between different interventions, stratified by energy prescription: energy prescribed (bottom) and *ad libitum* (top).

| Control | -0.87 (-1.53, -0.21) | -1 (-1.46, -0.53) | -1.31 (-2.24, -0.43) | -1.17 (-1.93, -0.5) | -0.86 (-1.25, -0.48) | -0.84 (-1.72, 0.03) |
| --- | --- | --- | --- | --- | --- | --- |
| -0.08 (-0.56, 0.44) | CR | -0.13 (-0.86, 0.61) | -0.44 (-1.56, 0.64) | -0.3 (-1.09, 0.38) | 0.01 (-0.65, 0.65) | 0.03 (-1.05, 1.08) |
| -0.39 (-0.73, 0.05) | -0.31 (-0.65, 0.05) | eTRE | -0.32 (-1.21, 0.53) | -0.18 (-0.97, 0.52) | 0.14 (-0.38, 0.63) | 0.15 (-0.83, 1.12) |
| / | / | / | eTRE + EX | 0.14 (-0.99, 1.21) | 0.46 (-0.48, 1.41) | 0.47 (-0.76, 1.74) |
| / | / | / | / | lTRE | 0.31 (-0.39, 1.09) | 0.33 (-0.74, 1.48) |
| -0.32 (-0.79, 0.16) | -0.25 (-0.58, 0.06) | 0.06 (-0.31, 0.39) | / | / | mTRE | 0.02 (-0.88, 0.91) |
| / | / | / | / | / | / | mTRE + EX |

**Footnotes:** CR = calorie restriction, eTRE = early time-restrcited eating, mTRE = midday time-restricted eating, lTRE = late time-restricted eating, eTRE + EX = early time-restricted eating + exercise, mTRE + EX = midday time-restricted eating + exercise.

**Supplementary Table 6:** Subgroup analysis presenting mean differences with 95% CI in BMI between different interventions, stratified by intervention duration: 11 weeks ≤ (bottom) and 10 weeks ≥ (top).

| Control | -0.82 (-1.41, -0.19) | -0.91 (-1.28, -0.46) | -1.01 (-1.73, -0.23) | -0.45 (-1.25, 0.45) | -0.87 (-1.21, -0.53) | -0.84 (-1.47, -0.21) |
| --- | --- | --- | --- | --- | --- | --- |
| -0.71 (-1.4, -0.07) | CR | -0.09 (-0.72, 0.58) | -0.19 (-1.13, 0.75) | 0.37 (-0.59, 1.38) | -0.06 (-0.59, 0.43) | -0.03 (-0.87, 0.77) |
| -0.82 (-1.43, -0.23) | -0.11 (-0.74, 0.55) | eTRE | -0.1 (-0.86, 0.64) | 0.47 (-0.28, 1.21) | 0.04 (-0.42, 0.41) | 0.07 (-0.69, 0.75) |
| -1.71 (-3.46, 0.04) | -1 (-2.77, 0.78) | -0.89 (-2.54, 0.77) | eTRE + EX | 0.57 (-0.47, 1.64) | 0.13 (-0.67, 0.9) | 0.17 (-0.83, 1.11) |
| -1.41 (-2.43, -0.53) | -0.7 (-1.63, 0.13) | -0.59 (-1.66, 0.35) | 0.3 (-1.71, 2.17) | lTRE | -0.43 (-1.33, 0.38) | -0.39 (-1.47, 0.59) |
| -0.76 (-1.35, -0.18) | -0.05 (-0.71, 0.63) | 0.06 (-0.58, 0.71) | 0.95 (-0.83, 2.72) | 0.65 (-0.3, 1.73) | mTRE | 0.03 (-0.62, 0.68) |
| / | / | / | / | / | / | mTRE + EX |

**Footnotes:** CR = calorie restriction, eTRE = early time-restrcited eating, mTRE = midday time-restricted eating, lTRE = late time-restricted eating, eTRE + EX = early time-restricted eating + exercise, mTRE + EX = midday time-restricted eating + exercise.

**Supplementary Table 7:** Subgroup analysis presenting mean differences with 95% CI in FM (kg) between different interventions, stratified by energy prescription: energy prescribed (bottom) and *ad libitum* (top).

| Control | 0.3 (-1.22, 1.83) | -0.75 (-1.43, 0.23) | / | -0.79 (-1.66, 0.24) | -0.01 (-0.61, 0.66) | -0.91 (-3.42, 1.59) |
| --- | --- | --- | --- | --- | --- | --- |
| 0.69 (-1.24, 2.55) | CR | -1.05 (-2.63, 0.78) | / | -1.09 (-2.83, 0.76) | -0.31 (-1.82, 1.25) | -1.2 (-4.14, 1.67) |
| -0.02 (-1.78, 1.74) | -0.71 (-1.7, 0.34) | eTRE | / | -0.03 (-1.08, 0.85) | 0.74 (-0.29, 1.55) | -0.18 (-2.89, 2.41) |
| / | / | / | eTRE + EX | / | / | / |
| / | / | / | / | lTRE | 0.78 (-0.36, 1.83) | -0.12 (-2.83, 2.5) |
| -0.28 (-2.03, 1.5) | -0.98 (-2.14, 0.29) | -0.26 (-1.51, 0.98) | / | / | mTRE | -0.9 (-3.51, 1.64) |
| / | / | / | / | / | / | mTRE + EX |

**Footnotes:** CR = calorie restriction, eTRE = early time-restrcited eating, mTRE = midday time-restricted eating, lTRE = late time-restricted eating, eTRE + EX = early time-restricted eating + exercise.

**Supplementary Table 8:** Subgroup analysis presenting mean differences with 95% CI in FM (kg) between different interventions, stratified by intervention duration: 11 weeks ≤ (bottom) and 10 weeks ≥ (top).

| Control | -1.92 (-3.83, -0.11) | -1 (-1.92, -0.02) | -4.08 (-6.36, -1.83) | -0.02 (-2.44, 2.44) | -1.45 (-2.41, -0.56) | -1.88 (-4.74, 1.07) |
| --- | --- | --- | --- | --- | --- | --- |
| -0.74 (-1.85, 0.24) | CR | 0.92 (-0.87, 2.89) | -2.15 (-4.95, 0.71) | 1.9 (-0.95, 4.92) | 0.47 (-1.13, 2.13) | 0.06 (-3.31, 3.57) |
| -1.24 (-2.13, -0.57) | -0.52 (-1.4, 0.4) | eTRE | -3.08 (-5.31, -0.96) | 0.97 (-1.27, 3.23) | -0.45 (-1.46, 0.43) | -0.88 (-3.9, 2.19) |
| -5.61 (-8.2, -2.97) | -4.85 (-7.45, -2.13) | -4.33 (-6.8, -1.76) | eTRE + EX | 4.06 (0.96, 7.24) | 2.63 (0.32, 4.92) | 2.21 (-1.43, 5.94) |
| -1.52 (-2.52, -0.58) | -0.78 (-2.07, 0.65) | -0.27 (-1.33, 0.94) | 4.06 (1.34, 6.8) | lTRE | -1.43 (-3.93, 0.96) | -1.86 (-5.63, 1.97) |
| -1.37 (-2.3, -0.6) | -0.63 (-1.72, 0.46) | -0.12 (-1.05, 0.8) | 4.22 (1.46, 6.82) | 0.15 (-1.17, 1.32) | mTRE | -0.42 (-3.39, 2.66) |
| / | / | / | / | / | / | mTRE + EX |

**Footnotes:** CR = calorie restriction, eTRE = early time-restrcited eating, mTRE = midday time-restricted eating, lTRE = late time-restricted eating, eTRE + EX = early time-restricted eating + exercise, mTRE + EX = midday time-restricted eating + exercise.

**Supplementary Table 9:** Subgroup analysis presenting mean differences with 95% CI in fat (%) between different interventions, stratified by energy prescription: energy prescribed (bottom) and *ad libitum* (top).

| Control | -2.85 (-5.06, -0.77) | -0.58 (-1.24, 0.11) | / | -0.8 (-2.42, 0.83) | -1.06 (-1.73, -0.5) | 0.71 (-0.86, 2.02) |
| --- | --- | --- | --- | --- | --- | --- |
| 0.51 (-0.94, 1.93) | CR | 2.27 (0.15, 4.55) | / | 2.05 (-0.55, 4.81) | 1.8 (-0.25, 3.86) | 3.56 (0.98, 6.01) |
| -0.15 (-1.46, 1.22) | -0.66 (-1.42, 0.17) | eTRE | / | -0.22 (-1.84, 1.38) | -0.47 (-1.35, 0.23) | 1.29 (-0.44, 2.72) |
| / | / | / | eTRE + EX | / | / | / |
| / | / | / | / | lTRE | -0.25 (-2.04, 1.36) | 1.52 (-0.81, 3.52) |
| 0.29 (-1.04, 1.64) | -0.21 (-0.94, 0.55) | 0.44 (-0.4, 1.23) | / | / | mTRE | 1.77 (0.25, 3.13) |
| / | / | / | / | / | / | mTRE + EX |

**Footnotes:** CR = calorie restriction, eTRE = early time-restrcited eating, mTRE = midday time-restricted eating, lTRE = late time-restricted eating, eTRE + EX = early time-restricted eating + exercise, mTRE + EX = midday time-restricted eating + exercise.

**Supplementary Table 10:** Subgroup analysis presenting mean differences with 95% CI in fat (%) between different interventions, stratified by intervention duration: 11 weeks ≤ (bottom) and 10 weeks ≥ (top).

| Control | -1.65 (-3.25, -0.29) | -0.58 (-1.44, 0.28) | / | / | -1.3 (-2.18, -0.6) | 0.54 (-1.21, 2.02) |
| --- | --- | --- | --- | --- | --- | --- |
| -0.06 (-0.89, 0.75) | CR | 1.07 (-0.36, 2.76) | / | / | 0.35 (-0.87, 1.63) | 2.18 (0.13, 4.2) |
| -0.85 (-1.42, -0.25) | -0.79 (-1.46, -0.06) | eTRE | / | / | -0.72 (-1.74, 0.09) | 1.29 (-0.44, 2.72) |
| / | / | / | eTRE + EX | / | / | 1.12 (-0.79, 2.74) |
| -0.93 (-1.98, 0.1) | -0.87 (-2.08, 0.35) | -0.09 (-1.13, 0.92) | / | lTRE | / | / |
| -0.03 (-0.61, 0.54) | 0.03 (-0.75, 0.83) | 0.83 (0.15, 1.46) | / | 0.91 (-0.22, 2.03) | mTRE | 1.84 (0.17, 3.41) |
| / | / | / | / | / | / | mTRE + EX |

**Footnotes:** CR = calorie restriction, eTRE = early time-restrcited eating, mTRE = midday time-restricted eating, lTRE = late time-restricted eating, eTRE + EX = early time-restricted eating + exercise, mTRE + EX = midday time-restricted eating + exercise.

**Supplementary Table 11:** Subgroup analysis presenting mean differences with 95% CI in LBM (kg) between different interventions, stratified by intervention duration: 11 weeks ≤ (bottom) and 10 weeks ≥ (top).

| Control | / | 0.04 (-2.16, 2.69) | / | 0.31 (-4.03, 5.25) | 0.54 (-1.96, 3.07) | -0.79 (-4.53, 3.04) |
| --- | --- | --- | --- | --- | --- | --- |
| -0.22 (-0.84, 0.43) | CR | / | / | / | / | / |
| -0.54 (-1.22, 0.12) | -0.33 (-0.94, 0.25) | eTRE | / | 0.29 (-3.66, 4.27) | 0.51 (-2.37, 2.95) | -0.85 (-5.34, 3.43) |
| / | / | / | eTRE + EX | / | / | / |
| -0.84 (-1.45, -0.22) | -0.63 (-1.42, 0.17) | -0.3 (-1.05, 0.47) | / | lTRE | 0.22 (-4.79, 4.75) | -1.13 (-7.17, 4.63) |
| -0.22 (-0.71, 0.26) | -0.01 (-0.71, 0.69) | 0.32 (-0.44, 1.11) | / | 0.62 (-0.14, 1.4) | mTRE | -1.33 (-5.63, 3.01) |
| / | / | / | / | / | / | mTRE + EX |

**Footnotes:** CR = calorie restriction, eTRE = early time-restrcited eating, mTRE = midday time-restricted eating, lTRE = late time-restricted eating, eTRE + EX = early time-restricted eating + exercise, mTRE + EX = midday time-restricted eating + exercise.

**Supplementary Table 12:** Subgroup analysis presenting mean differences with 95% CI in WC (cm) between different interventions, stratified by energy prescription: energy prescribed (bottom) and *ad libitum* (top).

| Control | -3.97 (-6.71, -1.28) | -2.41 (-4.61, -0.17) | -3.76 (-7.8, 0.31) | -2.18 (-5.66, 1.28) | -1.15 (-3.05, 0.8) | -2.18 (-8.18, 3.83) |
| --- | --- | --- | --- | --- | --- | --- |
| -3 (-5.2, -0.94) | CR | 1.56 (-1.24, 4.45) | 0.2 (-4.3, 4.86) | 1.8 (-1.76, 5.34) | 2.82 (0.24, 5.53) | 1.78 (-4.77, 8.43) |
| -3.76 (-5.6, -1.94) | -0.75 (-1.78, 0.37) | eTRE | -1.36 (-5.08, 2.39) | 0.25 (-3.34, 3.68) | 1.26 (-1.05, 3.58) | 0.23 (-6.17, 6.61) |
| / | / | / | eTRE + EX | 1.6 (-3.49, 6.55) | 2.61 (-1.6, 6.83) | 1.57 (-5.66, 8.85) |
| / | / | / | / | lTRE | 1.03 (-2.58, 4.71) | -0.01 (-6.95, 6.93) |
| -3.51 (-5.78, -1.31) | -0.5 (-1.54, 0.59) | 0.25 (-1.04, 1.51) | / | / | mTRE | -1.03 (-7.32, 5.27) |
| / | / | / | / | / | / | mTRE + EX |

**Footnotes:** CR = calorie restriction, eTRE = early time-restrcited eating, mTRE = midday time-restricted eating, lTRE = late time-restricted eating, eTRE + EX = early time-restricted eating + exercise, mTRE + EX = midday time-restricted eating + exercise.

**Supplementary Table 13:** Subgroup analysis presenting mean differences with 95% CI in WC (cm) between different interventions, stratified by intervention duration: 11 weeks ≤ (bottom) and 10 weeks ≥ (top).

| Control | -1.98 (-4.1, 0.17) | -2.45 (-4.28, -0.55) | -1.83 (-5.23, 1.63) | -2.27 (-6.26, 1.78) | -1.77 (-3.38, -0.15) | -2.17 (-6.33, 1.96) |
| --- | --- | --- | --- | --- | --- | --- |
| -3.38 (-6.37, -0.49) | CR | -0.47 (-2.43, 1.58) | 0.16 (-3.54, 3.92) | -0.29 (-4.34, 3.85) | 0.21 (-1.39, 1.77) | -0.18 (-4.84, 4.44) |
| -3.24 (-6.13, -0.39) | 0.13 (-2.34, 2.68) | eTRE | 0.63 (-2.68, 3.92) | 0.17 (-3.41, 3.76) | 0.68 (-1.09, 2.35) | 0.28 (-4.28, 4.78) |
| -7.04 (-14.05, -0.12) | -3.66 (-10.5, 3.21) | -3.81 (-10.17, 2.49) | eTRE + EX | -0.42 (-5.28, 4.39) | 0.06 (-3.49, 3.56) | -0.34 (-5.72, 4.98) |
| -1.86 (-6.56, 2.7) | 1.51 (-3.09, 6.14) | 1.39 (-3.58, 6.27) | 5.19 (-2.87, 13.2) | lTRE | 0.5 (-3.5, 4.43) | 0.11 (-5.66, 5.81) |
| -1.28 (-4.07, 1.54) | 2.1 (-0.64, 4.96) | 1.97 (-0.79, 4.79) | 5.77 (-1.11, 12.81) | 0.6 (-4.33, 5.63) | mTRE | -0.4 (-4.83, 4.01) |
| / | / | / | / | / | / | mTRE + EX |

**Footnotes:** CR = calorie restriction, eTRE = early time-restrcited eating, mTRE = midday time-restricted eating, lTRE = late time-restricted eating, eTRE + EX = early time-restricted eating + exercise, mTRE + EX = midday time-restricted eating + exercise.

**Supplementary Table 14:** Subgroup analysis presenting mean differences with 95% CI in FBG (mg/dl) between different interventions, stratified by energy prescription: energy prescribed (bottom) and *ad libitum* (top).

| Control | -3.36 (-9.8, 2.69) | -3.51 (-6.79, -0.33) | -8.35 (-19.38, 2.55) | -1.68 (-7.05, 3.52) | -1 (-5.02, 2.85) | / |
| --- | --- | --- | --- | --- | --- | --- |
| -2.4 (-6.22, 2) | CR | -0.16 (-6.17, 6.14) | -4.99 (-17, 7.27) | 1.66 (-6.09, 9.57) | 2.34 (-3.53, 8.44) | / |
| -4.51 (-7.59, -0.74) | -2.12 (-4.61, 0.41) | eTRE | -4.86 (-15.31, 5.55) | 1.82 (-3.6, 7.19) | 2.5 (-1.68, 6.59) | / |
| / | / | / | eTRE + EX | 6.69 (-5.09, 18.34) | 7.36 (-3.85, 18.56) | / |
| / | / | / | / | lTRE | 0.68 (-5.62, 6.93) | / |
| -0.65 (-4.57, 3.66) | 1.74 (-0.89, 4.14) | 3.86 (1.01, 6.52) | / | / | mTRE | / |
| / | / | / | / | / | / | mTRE + EX |

**Footnotes:** CR = calorie restriction, eTRE = early time-restrcited eating, mTRE = midday time-restricted eating, lTRE = late time-restricted eating, eTRE + EX = early time-restricted eating + exercise, mTRE + EX = midday time-restricted eating + exercise.

**Supplementary Table 15:** Subgroup analysis presenting mean differences with 95% CI in FBG (mg/dl) between different interventions, stratified by intervention duration: 11 weeks ≤ (bottom) and 10 weeks ≥ (top).

| Control | -0.93 (-6.97, 5.13) | -0.95 (-4.51, 2.48) | / | -0.85 (-9.47, 7.58) | 0.92 (-3.23, 5.16) | / |
| --- | --- | --- | --- | --- | --- | --- |
| -4.05 (-8.52, 0.14) | CR | -0.01 (-5.56, 5.37) | / | 0.07 (-9.46, 9.48) | 1.86 (-3.26, 7.01) | / |
| -6.48 (-9.81, -3.24) | -2.42 (-5.96, 1.26) | eTRE | / | 0.09 (-7.65, 7.93) | 1.87 (-1.84, 5.73) | / |
| -11.32 (-20.38, -2.36) | -7.29 (-16.32, 1.92) | -4.84 (-13.24, 3.56) | eTRE + EX | / | / | / |
| -1.33 (-6.56, 3.49) | 2.73 (-3.62, 8.96) | 5.15 (-0.53, 10.53) | 9.99 (-0.24, 19.83) | lTRE | 1.78 (-6.74, 10.58) | / |
| -1.99 (-6.09, 1.89) | 2.08 (-2.11, 6.34) | 4.5 (0.43, 8.47) | 9.34 (-0.05, 18.63) | -0.66 (-6.76, 5.66) | mTRE | / |
| / | / | / | / | / | / | mTRE + EX |

**Footnotes:** CR = calorie restriction, eTRE = early time-restrcited eating, mTRE = midday time-restricted eating, lTRE = late time-restricted eating, eTRE + EX = early time-restricted eating + exercise, mTRE + EX = midday time-restricted eating + exercise.

**Supplementary Table 16:** Subgroup analysis presenting mean differences with 95% CI in FBI (mIU/L) between different interventions, stratified by energy prescription: energy prescribed (bottom) and *ad libitum* (top).

| Control | -1.09 (-3.8, 1.54) | -1.37 (-2.96, 0.03) | -5.27 (-9.14, -1.58) | -2.38 (-5.01, -0.06) | -1.51 (-3.07, 0) | 1.01 (-3.57, 5.53) |
| --- | --- | --- | --- | --- | --- | --- |
| 3.22 (-1.36, 7.82) | CR | -0.3 (-2.96, 2.36) | -4.19 (-8.57, 0.14) | -1.3 (-4.86, 2.08) | -0.42 (-3.03, 2.21) | 2.1 (-3.17, 7.41) |
| -0.03 (-3.76, 3.11) | -3.33 (-6.82, -0.18) | eTRE | -3.9 (-7.36, -0.43) | -0.99 (-3.6, 1.43) | -0.13 (-1.82, 1.67) | 2.39 (-2.37, 7.22) |
| / | / | / | eTRE + EX | 2.9 (-1.43, 7.06) | 3.77 (-0.05, 7.68) | 6.28 (0.5, 12.22) |
| / | / | / | / | lTRE | 0.87 (-1.79, 3.8) | 3.4 (-1.68, 8.7) |
| 4.07 (-0.39, 8.52) | 0.85 (-1.93, 3.59) | 4.17 (1.03, 7.59) | / | / | mTRE | 2.52 (-2.28, 7.32) |
| / | / | / | / | / | / | mTRE + EX |

**Footnotes:** CR = calorie restriction, eTRE = early time-restrcited eating, mTRE = midday time-restricted eating, lTRE = late time-restricted eating, eTRE + EX = early time-restricted eating + exercise, mTRE + EX = midday time-restricted eating + exercise.

**Supplementary Table 17:** Subgroup analysis presenting mean differences with 95% CI in FBI (mIU/L) between different interventions, stratified by intervention duration: 11 weeks ≤ (bottom) and 10 weeks ≥ (top).

| Control | -1.95 (-5.07, 1.51) | -2.09 (-4.42, 0.24) | / | -2.97 (-9.84, 4.1) | -0.84 (-3.14, 1.74) | 1 (-3.71, 5.66) |
| --- | --- | --- | --- | --- | --- | --- |
| 0.19 (-3.07, 3.8) | CR | -0.14 (-3.13, 2.51) | / | -1.02 (-8.14, 6.05) | 1.1 (-1.45, 3.66) | 2.95 (-2.92, 8.51) |
| -1.19 (-3.71, 1.14) | -1.4 (-5.1, 1.89) | eTRE | / | -0.87 (-7.32, 5.71) | 1.25 (-0.67, 3.46) | 3.1 (-2.13, 8.28) |
| -5.1 (-10.76, 0.42) | -5.29 (-11.66, 0.52) | -3.92 (-8.9, 1.09) | eTRE + EX | / | / | / |
| -2.5 (-6.32, 0.86) | -2.7 (-7.91, 1.71) | -1.3 (-5.36, 2.39) | 2.63 (-3.93, 8.78) | lTRE | 2.14 (-4.66, 8.98) | 3.94 (-4.37, 12.28) |
| -1.21 (-3.72, 1.3) | -1.4 (-4.94, 1.78) | 0 (-3, 3.14) | 3.9 (-1.94, 9.85) | 1.29 (-2.8, 5.78) | mTRE | 1.82 (-3.58, 6.96) |
| / | / | / | / | / | / | mTRE + EX |

**Footnotes:** CR = calorie restriction, eTRE = early time-restrcited eating, mTRE = midday time-restricted eating, lTRE = late time-restricted eating, eTRE + EX = early time-restricted eating + exercise, mTRE + EX = midday time-restricted eating + exercise.

**Supplementary Table 18:** Subgroup analysis presenting mean differences with 95% CI in HOMA-IR between different interventions, stratified by energy prescription: energy prescribed (bottom) and *ad libitum* (top).

| Control | -0.25 (-1.15, 0.64) | -0.38 (-0.86, 0.09) | -1.28 (-2.72, 0.12) | -0.6 (-1.58, 0.3) | -0.22 (-0.72, 0.27) | / |
| --- | --- | --- | --- | --- | --- | --- |
| 1.54 (-0.53, 3.55) | CR | -0.13 (-1.02, 0.75) | -1.03 (-2.64, 0.57) | -0.36 (-1.65, 0.89) | 0.03 (-0.84, 0.9) | / |
| 1.01 (-1.07, 2.98) | -0.53 (-1.08, 0.02) | eTRE | -0.9 (-2.24, 0.43) | -0.22 (-1.23, 0.73) | 0.16 (-0.39, 0.72) | / |
| / | / | / | eTRE + EX | 0.68 (-1.01, 2.32) | 1.06 (-0.37, 2.51) | / |
| / | / | / | / | lTRE | 0.39 (-0.63, 1.45) | / |
| 1.94 (-0.11, 3.94) | 0.41 (-0.12, 0.89) | 0.93 (0.3, 1.56) | / | / | mTRE | / |
| / | / | / | / | / | / | mTRE + EX |

**Footnotes:** CR = calorie restriction, eTRE = early time-restrcited eating, mTRE = midday time-restricted eating, lTRE = late time-restricted eating, eTRE + EX = early time-restricted eating + exercise, mTRE + EX = midday time-restricted eating + exercise.

**Supplementary Table 19:** Subgroup analysis presenting mean differences with 95% CI in HOMA-IR between different interventions, stratified by intervention duration: 11 weeks ≤ (bottom) and 10 weeks ≥ (top).

| Control | -0.02 (-1.07, 1.11) | -0.13 (-0.76, 0.53) | / | / | 0.29 (-0.4, 1.05) | / |
| --- | --- | --- | --- | --- | --- | --- |
| -0.31 (-1.04, 0.41) | CR | -0.11 (-1.12, 0.86) | / | / | 0.31 (-0.62, 1.24) | / |
| -0.68 (-1.36, -0.12) | -0.38 (-1.07, 0.22) | eTRE | / | / | 0.42 (-0.2, 1.09) | / |
| -1.58 (-2.93, -0.36) | -1.27 (-2.64, -0.04) | -0.9 (-2.04, 0.22) | eTRE + EX | / | / | / |
| -0.65 (-1.56, 0.1) | -0.34 (-1.45, 0.61) | 0.04 (-0.89, 0.9) | 0.93 (-0.54, 2.35) | lTRE | / | / |
| -0.42 (-1.01, 0.14) | -0.11 (-0.86, 0.6) | 0.26 (-0.4, 1) | 1.16 (-0.12, 2.54) | 0.23 (-0.68, 1.27) | mTRE | / |
| / | / | / | / | / | / | mTRE + EX |

**Footnotes:** CR = calorie restriction, eTRE = early time-restrcited eating, mTRE = midday time-restricted eating, lTRE = late time-restricted eating, eTRE + EX = early time-restricted eating + exercise, mTRE + EX = midday time-restricted eating + exercise.

**Supplementary Table 20:** Subgroup analysis presenting mean differences with 95% CI in TC (mg/dl) between different interventions, stratified by energy prescription: energy prescribed (bottom) and *ad libitum* (top).

| Control | -1.47 (-10.26, 7.39) | -0.82 (-6.88, 5.29) | -35.55 (-56.97, -13.91) | -2.36 (-11.6, 7.49) | 2.07 (-3.6, 7.95) | 2.63 (-9.89, 15.34) |
| --- | --- | --- | --- | --- | --- | --- |
| -2.28 (-8.67, 5.21) | CR | 0.65 (-8.75, 10.04) | -34.05 (-56.79, -11.31) | -0.88 (-12.21, 10.99) | 3.54 (-5.31, 12.53) | 4.09 (-10.9, 19.25) |
| -0.89 (-6.07, 5.64) | 1.31 (-4.09, 6.73) | eTRE | -34.74 (-55.31, -13.89) | -1.52 (-11.32, 8.61) | 2.88 (-3.92, 9.76) | 3.45 (-10.18, 17.27) |
| -26.02 (-34.45, -16.65) | -23.93 (-34.18, -13.1) | -25.16 (-35.11, -15.19) | eTRE + EX | 33.24 (10.16, 56.22) | 37.6 (15.88, 59.47) | 38.14 (13.39, 62.99) |
| / | / | / | / | lTRE | 4.41 (-6.29, 14.78) | 4.98 (-10.91, 20.55) |
| 0.95 (-3.12, 7.88) | 3.49 (-2.51, 9.85) | 2.11 (-3.34, 8.1) | 27.28 (18.53, 36.49) | / | mTRE | 0.56 (-12.48, 13.64) |
| -10.59 (-24.19, 4.39) | -8.48 (-23.46, 7.47) | -9.76 (-24.36, 5.66) | 15.29 (-0.07, 31.28) | / | -11.9 (-25.84, 2.8) | mTRE + EX |

**Footnotes:** CR = calorie restriction, eTRE = early time-restrcited eating, mTRE = midday time-restricted eating, lTRE = late time-restricted eating, eTRE + EX = early time-restricted eating + exercise, mTRE + EX = midday time-restricted eating + exercise.

**Supplementary Table 21:** Subgroup analysis presenting mean differences with 95% CI in TC (mg/dl) between different interventions, stratified by intervention duration: 11 weeks ≤ (bottom) and 10 weeks ≥ (top).

| Control | 3.15 (-5.02, 13.57) | 3.01 (-2.02, 9.6) | / | 6.88 (-10.79, 25.25) | 4.58 (-0.15, 11.61) | 3.03 (-5.37, 12.86) |
| --- | --- | --- | --- | --- | --- | --- |
| -4.14 (-12.32, 3.91) | CR | -0.09 (-8.97, 7.89) | / | 3.68 (-15.81, 22.47) | 1.44 (-6.32, 9.18) | -0.08 (-12.51, 11.36) |
| -2.31 (-9.02, 4.61) | 1.83 (-5.83, 9.8) | eTRE | / | 3.75 (-13.38, 21) | 1.46 (-3.52, 7.62) | -0.04 (-9.97, 9.94) |
| -30.32 (-43.28, -17.61) | -26.16 (-40.42, -11.94) | -28 (-41.34, -15.15) | eTRE + EX | / | / | / |
| -4.4 (-14.36, 6.35) | -0.26 (-11.57, 11.73) | -2.12 (-13, 9.33) | 25.88 (10.36, 42.37) | lTRE | -2.22 (-19.95, 16.07) | -3.79 (-23.67, 16.08) |
| 0.04 (-6.64, 6.85) | 4.17 (-4.49, 13.01) | 2.31 (-5.75, 10.39) | 30.34 (17.36, 43.68) | 4.43 (-7.59, 15.93) | mTRE | -1.55 (-11.34, 7.36) |
| -12.17 (-31.19, 6.79) | -7.98 (-28.27, 12.28) | -9.85 (-29.8, 9.79) | 18.19 (-2.41, 38.67) | -7.72 (-29.55, 13.37) | -12.21 (-31.4, 6.79) | mTRE + EX |

**Footnotes:** CR = calorie restriction, eTRE = early time-restrcited eating, mTRE = midday time-restricted eating, lTRE = late time-restricted eating, eTRE + EX = early time-restricted eating + exercise, mTRE + EX = midday time-restricted eating + exercise.

**Supplementary Table 22:** Subgroup analysis presenting mean differences with 95% CI in LDL-C (mg/dl) between different interventions, stratified by energy prescription: energy prescribed (bottom) and *ad libitum* (top).

| Control | -0.05 (-7.78, 7.66) | 0.03 (-5.33, 5.17) | -15.47 (-34.25, 2.88) | 2.08 (-5.39, 10.13) | 4.91 (-0.07, 10.19) | 1.54 (-12.59, 15.86) |
| --- | --- | --- | --- | --- | --- | --- |
| -0.32 (-7.05, 7.05) | CR | 0.09 (-8.17, 8.15) | -15.37 (-35.2, 3.87) | 2.09 (-7.42, 12.35) | 4.95 (-2.72, 12.9) | 1.57 (-14.2, 17.46) |
| 0.23 (-5.46, 6.6) | 0.58 (-4.71, 5.82) | eTRE | -15.5 (-33.34, 2.18) | 2.06 (-6.14, 11.13) | 4.87 (-0.87, 11.16) | 1.49 (-13.22, 16.57) |
| -15.49 (-24.92, -5) | -15.12 (-25.92, -3.83) | -15.78 (-26.3, -4.83) | eTRE + EX | 17.52 (-1.76, 37.79) | 20.36 (1.9, 39.46) | 16.99 (-5.95, 40.37) |
| / | / | / | / | lTRE | 2.84 (-6.16, 11.53) | -0.53 (-16.87, 15.23) |
| 0.98 (-3.56, 8.39) | 1.62 (-3.94, 8.38) | 1.04 (-4.22, 7.63) | 16.81 (7.13, 26.97) | / | mTRE | -3.4 (-17.8, 10.88) |
| 8.32 (-2.19, 19.87) | 8.54 (-3.21, 21.02) | 8.02 (-3.39, 20.11) | 23.79 (10.46, 36.96) | / | 6.97 (-4.21, 17.96) | mTRE + EX |

**Footnotes:** CR = calorie restriction, eTRE = early time-restrcited eating, mTRE = midday time-restricted eating, lTRE = late time-restricted eating, eTRE + EX = early time-restricted eating + exercise, mTRE + EX = midday time-restricted eating + exercise.

**Supplementary Table 23:** Subgroup analysis presenting mean differences with 95% CI in LDL-C (mg/dl) between different interventions, stratified by intervention duration: 11 weeks ≤ (bottom) and 10 weeks ≥ (top).

| Control | 5.86 (-2.02, 15.18) | 4.53 (-0.35, 10.24) | / | 39.42 (16.45, 62.72) | 5.61 (0.73, 11.86) | 2.05 (-8.76, 13.28) |
| --- | --- | --- | --- | --- | --- | --- |
| -3.07 (-8.86, 2.81) | CR | -1.32 (-9.53, 6.14) | / | 33.47 (9.34, 57.18) | -0.29 (-7.55, 7) | -3.79 (-17.42, 8.76) |
| -2.19 (-6.82, 2.68) | 0.88 (-4.63, 6.56) | eTRE | / | 34.85 (12.31, 57.32) | 1.04 (-3.84, 6.66) | -2.51 (-14.26, 8.95) |
| -15.36 (-24.58, -6.06) | -12.28 (-22.48, -1.97) | -13.18 (-22.57, -3.87) | eTRE + EX | / | / | / |
| -2.19 (-8.62, 4.5) | 0.86 (-6.76, 8.83) | -0.02 (-7.34, 7.41) | 13.18 (2.17, 24.34) | lTRE | -33.75 (-56.78, -10.44) | -37.38 (-62.54, -12.12) |
| 3.95 (-0.59, 9.1) | 7.07 (0.77, 13.67) | 6.17 (0.52, 12.05) | 19.33 (9.95, 29.08) | 6.18 (-1.57, 14.05) | mTRE | -3.57 (-15.13, 7.11) |
| 9.57 (-2.64, 22.01) | 12.69 (-0.62, 25.94) | 11.76 (-1.07, 24.61) | 24.94 (10.81, 38.86) | 11.81 (-2.1, 25.53) | 5.59 (-6.82, 17.87) | mTRE + EX |

**Footnotes:** CR = calorie restriction, eTRE = early time-restrcited eating, mTRE = midday time-restricted eating, lTRE = late time-restricted eating, eTRE + EX = early time-restricted eating + exercise, mTRE + EX = midday time-restricted eating + exercise.

**Supplementary Table 24:** Subgroup analysis presenting mean differences with 95% CI in HDL (mg/dl) between different interventions, stratified by energy prescription: energy prescribed (bottom) and *ad libitum* (top).

| Control | 0.32 (-2.34, 3.22) | -0.07 (-1.41, 1.18) | 19.33 (11.71, 26.79) | 0.01 (-2.48, 2.55) | 1.04 (-0.18, 2.41) | 0.34 (-2.52, 3.16) |
| --- | --- | --- | --- | --- | --- | --- |
| -0.59 (-3.47, 3.72) | CR | -0.4 (-3.38, 2.27) | 18.99 (10.93, 26.8) | -0.32 (-4.08, 3.22) | 0.72 (-2.09, 3.4) | 0.03 (-4.03, 3.7) |
| 0.08 (-2.08, 3.87) | 0.76 (-1.44, 2.95) | eTRE | 19.42 (11.88, 26.75) | 0.08 (-2.35, 2.63) | 1.11 (-0.26, 2.74) | 0.41 (-2.6, 3.48) |
| / | / | / | eTRE + EX | -19.33 (-27.05, -11.39) | -18.28 (-25.73, -10.55) | -18.99 (-26.88, -10.87) |
| / | / | / | / | lTRE | 1.02 (-1.62, 3.81) | 0.34 (-3.44, 4.04) |
| 0.66 (-3.17, 5.34) | 1.18 (-2.45, 4.56) | 0.41 (-3.31, 3.96) | / | / | mTRE | -0.67 (-3.71, 2.08) |
| / | / | / | / | / | / | mTRE + EX |

**Footnotes:** CR = calorie restriction, eTRE = early time-restrcited eating, mTRE = midday time-restricted eating, lTRE = late time-restricted eating, eTRE + EX = early time-restricted eating + exercise, mTRE + EX = midday time-restricted eating + exercise.

**Supplementary Table 25:** Subgroup analysis presenting mean differences with 95% CI in HDL-C (mg/dl) between different interventions, stratified by intervention duration: 11 weeks ≤ (bottom) and 10 weeks ≥ (top).

| Control | -0.29 (-3.88, 3.76) | -0.13 (-2.19, 1.74) | / | -0.15 (-5.74, 5.23) | 0.57 (-1.35, 2.72) | 0.1 (-3.38, 3.51) |
| --- | --- | --- | --- | --- | --- | --- |
| -0.33 (-2.18, 1.9) | CR | 0.14 (-3.79, 3.51) | / | 0.14 (-6.53, 6.13) | 0.85 (-2.74, 4.24) | 0.43 (-4.84, 4.99) |
| -0.04 (-1.16, 1.65) | 0.35 (-1.43, 2.09) | eTRE | / | -0.01 (-5.14, 5.11) | 0.72 (-1.14, 2.97) | 0.23 (-3.45, 4.04) |
| 19.47 (12.03, 26.68) | 19.76 (12.19, 26.94) | 19.4 (12.07, 26.38) | eTRE + EX | / | / | / |
| 0.06 (-2.43, 2.74) | 0.4 (-2.83, 3.42) | 0.06 (-2.76, 2.67) | -19.36 (-26.93, -11.57) | lTRE | 0.74 (-4.62, 6.45) | 0.25 (-6.07, 6.67) |
| 1.59 (-0.01, 3.25) | 1.91 (-0.54, 4.06) | 1.59 (-0.5, 3.33) | -17.87 (-25.18, -10.31) | 1.52 (-1.54, 4.48) | mTRE | -0.42 (-4.23, 2.92) |
| / | / | / | / | / | / | mTRE + EX |

**Footnotes:** CR = calorie restriction, eTRE = early time-restrcited eating, mTRE = midday time-restricted eating, lTRE = late time-restricted eating, eTRE + EX = early time-restricted eating + exercise, mTRE + EX = midday time-restricted eating + exercise.

**Supplementary Table 26:** Subgroup analysis presenting mean differences with 95% CI in SBP (mmHg) between different interventions, stratified by intervention duration: 11 weeks ≤ (bottom) and 10 weeks ≥ (top).

| Control | -1.21 (-10.53, 7.58) | -1.79 (-6.56, 3.06) | -1.58 (-11.75, 8.66) | 2.98 (-10.87, 16.85) | -0.72 (-5.88, 4.18) | -4.4 (-10.55, 1.67) |
| --- | --- | --- | --- | --- | --- | --- |
| -1.79 (-6.07, 2.17) | CR | -0.62 (-9.75, 9.09) | -0.37 (-13.55, 13.1) | 4.23 (-11.7, 20.41) | 0.46 (-6.97, 8.1) | -3.22 (-13.29, 7.3) |
| -2.96 (-6.87, 0.63) | -1.17 (-4.59, 2.25) | eTRE | 0.19 (-10.06, 10.47) | 4.76 (-8.22, 17.81) | 1.08 (-4.86, 6.63) | -2.59 (-10.18, 4.77) |
| / | / | / | eTRE + EX | 4.63 (-12.15, 21.21) | 0.85 (-10.31, 11.8) | -2.8 (-14.74, 8.89) |
| -1.54 (-6.5, 2.91) | 0.25 (-5.65, 6) | 1.43 (-4.03, 6.72) | / | lTRE | -3.74 (-18.02, 10.39) | -7.42 (-22.33, 7.54) |
| -0.89 (-4.23, 2.19) | 0.91 (-3.1, 4.97) | 2.08 (-1.72, 5.93) | / | 0.65 (-4.7, 6.23) | mTRE | -3.67 (-10.5, 3.35) |
| / | / | / | / | / | / | mTRE + EX |

**Footnotes:** CR = calorie restriction, eTRE = early time-restrcited eating, mTRE = midday time-restricted eating, lTRE = late time-restricted eating, eTRE + EX = early time-restricted eating + exercise, mTRE + EX = midday time-restricted eating + exercise.

**Supplementary Table 27:** Subgroup analysis presenting mean differences with 95% CI in DBP (mmHg) between different interventions, stratified by intervention duration: 11 weeks ≤ (bottom) and 10 weeks ≥ (top).

| Control | 4.32 (-4.96, 12.08) | -0.28 (-4.92, 4.23) | -1.53 (-10.49, 7.5) | -0.07 (-12.66, 12.47) | -0.43 (-5.35, 4.25) | 0.54 (-5.57, 6.45) |
| --- | --- | --- | --- | --- | --- | --- |
| 0.28 (-1.81, 2.21) | CR | -4.6 (-12.73, 4.96) | -5.77 (-17.15, 6.92) | -4.27 (-18.58, 11.03) | -4.74 (-11.16, 2.93) | -3.79 (-12.89, 6.66) |
| -1.78 (-3.63, -0.07) | -2.05 (-3.68, -0.44) | eTRE | -1.22 (-10.08, 7.68) | 0.21 (-11.48, 12.05) | -0.15 (-5.61, 5.18) | 0.81 (-6.48, 8.03) |
| / | / | / | eTRE + EX | 1.48 (-13.31, 16.17) | 1.09 (-8.78, 10.76) | 2.05 (-8.69, 12.73) |
| -1.14 (-3.69, 1.19) | -1.42 (-4.33, 1.38) | 0.65 (-2.06, 3.25) | / | lTRE | -0.37 (-13.4, 12.54) | 0.56 (-13.27, 14.34) |
| -1 (-2.68, 0.45) | -1.3 (-3.28, 0.73) | 0.76 (-1.08, 2.64) | / | 0.13 (-2.58, 2.94) | mTRE | 0.97 (-5.81, 7.72) |
| / | / | / | / | / | / | mTRE + EX |

**Footnotes:** CR = calorie restriction, eTRE = early time-restrcited eating, mTRE = midday time-restricted eating, lTRE = late time-restricted eating, eTRE + EX = early time-restricted eating + exercise, mTRE + EX = midday time-restricted eating + exercise.

**Sensitivity analyses:**

**Supplementary Table 28:** Sensitivity analysis presenting mean differences with 95% CI in BW (kg) (top) and BMI (bottom) between different dietary approaches, after exclusion of studies including subjects with normal BMI and keeping studies with overweight and obese subjects.

| Control | -1.91 (-2.76, -1.07) | -2.28 (-2.93, -1.64) | -3.3 (-5.02, -1.66) | -2.05 (-3.1, -0.99) | -1.96 (-2.62, -1.3) | -3.01 (-7.06, 1.06) |
| --- | --- | --- | --- | --- | --- | --- |
| -0.67 (-1.12, -0.22) | CR | -0.37 (-1.14, 0.4) | -1.39 (-3.21, 0.36) | -0.14 (-1.27, 1.02) | -0.05 (-0.83, 0.75) | -1.1 (-5.23, 3.03) |
| -0.86 (-1.22, -0.5) | -0.19 (-0.63, 0.25) | eTRE | -1.02 (-2.71, 0.6) | 0.23 (-0.79, 1.28) | 0.33 (-0.41, 1.06) | -0.73 (-4.82, 3.38) |
| -1.22 (-2.06, -0.4) | -0.56 (-1.45, 0.34) | -0.36 (-1.18, 0.43) | eTRE + EX | 1.25 (-0.6, 3.21) | 1.35 (-0.36, 3.13) | 0.29 (-4.05, 4.71) |
| -1.03 (-1.71, -0.43) | -0.36 (-1.02, 0.22) | -0.17 (-0.84, 0.42) | 0.19 (-0.83, 1.15) | lTRE | 0.09 (-1.06, 1.22) | -0.96 (-5.15, 3.23) |
| -0.79 (-1.14, -0.45) | -0.12 (-0.53, 0.29) | 0.07 (-0.32, 0.46) | 0.43 (-0.43, 1.3) | 0.24 (-0.38, 0.94) | mTRE | -1.05 (-5.15, 3.06) |
| -0.89 (-2.33, 0.53) | -0.22 (-1.72, 1.27) | -0.03 (-1.51, 1.44) | 0.33 (-1.33, 2) | 0.14 (-1.4, 1.73) | -0.11 (-1.57, 1.37) | mTRE + EX |

**Footnotes:** CR = calorie restriction, eTRE = early time-restrcited eating, mTRE = midday time-restricted eating, lTRE = late time-restricted eating, eTRE + EX = early time-restricted eating + exercise, mTRE + EX = midday time-restricted eating + exercise.

**Supplementary Table 29:** Sensitivity analysis presenting mean differences with 95% CI in BW (kg) (top) and BMI (bottom) between different dietary approaches, after exclusion of studies including healthy subjects and keeping studies with chronically ill subjects.

| Control | -1.57 (-2.62, -0.5) | -1.87 (-2.57, -1.08) | -3.52 (-4.98, -2.05) | -1.72 (-2.58, -0.83) |
| --- | --- | --- | --- | --- |
| -0.55 (-1.15, 0.03) | CR | -0.29 (-1.15, 0.58) | -1.94 (-3.76, -0.16) | -0.15 (-1.16, 0.84) |
| -0.77 (-1.22, -0.31) | -0.21 (-0.74, 0.32) | eTRE | -1.65 (-3.32, -0.04) | 0.14 (-0.67, 0.93) |
| -1.3 (-2.18, -0.41) | -0.75 (-1.8, 0.33) | -0.53 (-1.54, 0.45) | lTRE | 1.8 (0.09, 3.5) |
| -0.64 (-1.11, -0.17) | -0.09 (-0.62, 0.46) | 0.13 (-0.31, 0.55) | 0.66 (-0.34, 1.67) | mTRE |

**Footnotes:** CR = calorie restriction, eTRE = early time-restrcited eating, mTRE = midday time-restricted eating, lTRE = late time-restricted eating.

**Supplementary Table 30:** Sensitivity analysis presenting mean differences with 95% CI in FM (kg) (top) and LBM (kg) (bottom) between different dietary approaches, after exclusion of studies including subjects with normal BMI and keeping studies with overweight and obese subjects.

| Control | -1.24 (-2.13, -0.37) | -1.35 (-2.01, -0.7) | -4.73 (-6.33, -3.17) | -1.21 (-2.21, -0.2) | -1.49 (-2.15, -0.85) | -2.01 (-4.83, 0.78) |
| --- | --- | --- | --- | --- | --- | --- |
| -0.41 (-1, 0.25) | CR | -0.11 (-0.96, 0.75) | -3.49 (-5.19, -1.8) | 0.02 (-1.21, 1.29) | -0.25 (-1.08, 0.57) | -0.77 (-3.71, 2.16) |
| -0.91 (-1.39, -0.3) | -0.5 (-1.07, 0.12) | eTRE | -3.37 (-4.87, -1.91) | 0.14 (-0.86, 1.17) | -0.14 (-0.86, 0.58) | -0.65 (-3.54, 2.2) |
| / | / | / | eTRE + EX | 3.52 (1.76, 5.31) | 3.24 (1.64, 4.87) | 2.72 (-0.49, 5.91) |
| -0.86 (-1.43, -0.24) | -0.45 (-1.25, 0.31) | 0.04 (-0.62, 0.65) | / | lTRE | -0.27 (-1.43, 0.84) | -0.8 (-3.78, 2.15) |
| -0.24 (-0.68, 0.2) | 0.17 (-0.52, 0.81) | 0.67 (0.01, 1.21) | / | 0.63 (-0.11, 1.31) | mTRE | -0.52 (-3.4, 2.35) |
| -0.99 (-3.51, 1.51) | -0.57 (-3.27, 2) | -0.09 (-2.72, 2.47) | / | -0.13 (-2.71, 2.44) | -0.75 (-3.33, 1.79) | mTRE + EX |

**Footnotes:** CR = calorie restriction, eTRE = early time-restrcited eating, mTRE = midday time-restricted eating, lTRE = late time-restricted eating, eTRE + EX = early time-restricted eating + exercise, mTRE + EX = midday time-restricted eating + exercise.

**Supplementary Table 31:** Sensitivity analysis presenting mean differences with 95% CI in FM (kg) (top) between different dietary approaches, after exclusion of studies including healthy subjects and keeping studies with chronically ill subjects.

| Control | -1.9 (-4.15, 0.29) | -2.01 (-4.39, 0.45) | -2.09 (-4.36, 0.16) |
| --- | --- | --- | --- |
| / | CR | -0.12 (-1.73, 1.67) | -0.2 (-1.86, 1.55) |
| / | / | eTRE | -0.08 (-1.39, 1.09) |
| / | / | / | mTRE |

**Footnotes:** CR = calorie restriction, eTRE = early time-restrcited eating, mTRE = midday time-restricted eating.

**Supplementary Table 32:** Sensitivity analysis presenting mean differences with 95% CI in fat (%) (top) and WC (cm) (bottom) between different dietary approaches, after exclusion of studies including subjects with normal BMI and keeping studies with overweight and obese subjects.

| Control | -0.54 (-1.44, 0.31) | -0.73 (-1.4, -0.04) | / | -0.87 (-2.39, 0.67) | -0.61 (-1.2, -0.03) | -0.97 (-3.82, 1.83) |
| --- | --- | --- | --- | --- | --- | --- |
| -2.7 (-4.54, -0.89) | CR | -0.19 (-0.98, 0.66) | / | -0.32 (-1.96, 1.4) | -0.07 (-0.79, 0.7) | -0.41 (-3.38, 2.5) |
| -2.67 (-4.32, -1) | 0.03 (-1.54, 1.66) | eTRE | / | -0.14 (-1.65, 1.4) | 0.12 (-0.58, 0.79) | -0.24 (-3.15, 2.62) |
| -3.99 (-7.45, -0.47) | -1.28 (-4.83, 2.32) | -1.32 (-4.59, 2) | eTRE + EX | / | / | / |
| -1.76 (-4.72, 1.16) | 0.94 (-1.97, 3.82) | 0.9 (-2.02, 3.78) | 2.23 (-2.19, 6.47) | lTRE | 0.26 (-1.34, 1.82) | -0.1 (-3.35, 3.06) |
| -1.6 (-3.15, 0.02) | 1.1 (-0.44, 2.72) | 1.07 (-0.58, 2.74) | 2.39 (-1.2, 5.98) | 0.16 (-2.82, 3.2) | mTRE | -0.35 (-3.27, 2.48) |
| -2.23 (-7.53, 3.13) | 0.48 (-5.15, 6.17) | 0.45 (-5.14, 6.09) | 1.78 (-4.63, 8.14) | -0.47 (-6.49, 5.66) | -0.62 (-6.19, 4.96) | mTRE + EX |

**Footnotes:** CR = calorie restriction, eTRE = early time-restrcited eating, mTRE = midday time-restricted eating, lTRE = late time-restricted eating, eTRE + EX = early time-restricted eating + exercise, mTRE + EX = midday time-restricted eating + exercise.

**Supplementary Table 33:** Sensitivity analysis presenting mean differences with 95% CI in FM (%) (top) and WC (cm) (bottom) between different dietary approaches, after exclusion of studies including healthy subjects and keeping studies with chronically ill subjects.

| Control | 0.3 (-1.2, 1.45) | -0.13 (-1.26, 0.69) | / | / | 0.27 (-0.94, 1.22) | / |
| --- | --- | --- | --- | --- | --- | --- |
| -3.65 (-5.17, -2.34) | CR | -0.46 (-1.38, 0.58) | / | / | -0.02 (-1.05, 1.08) | / |
| -3.82 (-5.06, -2.62) | -0.17 (-1.09, 0.92) | eTRE | / | / | 0.44 (-0.4, 1.23) | / |
| / | / | / | eTRE + EX | / | / | / |
| -3.77 (-6.29, -1.16) | -0.09 (-2.94, 2.97) | 0.07 (-2.71, 2.96) | / | lTRE | / | / |
| -3.37 (-4.66, -2.03) | 0.28 (-0.75, 1.59) | 0.45 (-0.51, 1.52) | / | 0.39 (-2.5, 3.23) | mTRE | / |
| -2.23 (-5.68, 1.33) | 1.44 (-2.27, 5.21) | 1.61 (-2.06, 5.3) | / | 1.56 (-2.79, 5.78) | 1.15 (-2.55, 4.85) | mTRE + EX |

**Footnotes:** CR = calorie restriction, eTRE = early time-restrcited eating, mTRE = midday time-restricted eating, lTRE = late time-restricted eating, eTRE + EX = early time-restricted eating + exercise, mTRE + EX = midday time-restricted eating + exercise.

**Supplementary Table 34:** Sensitivity analysis presenting mean differences with 95% CI in FBG (mg/dl) (top) and FBI (mIU/L) (bottom) between different dietary approaches, after exclusion of studies including subjects with normal BMI and keeping studies with overweight and obese subjects.

| Control | -2.27 (-6.05, 1.44) | -3.66 (-6.4, -0.94) | -8.56 (-17.81, 0.65) | -1.66 (-6.23, 2.77) | -0.55 (-3.9, 2.75) | / |
| --- | --- | --- | --- | --- | --- | --- |
| -0.78 (-2.9, 1.39) | CR | -1.4 (-4.53, 1.84) | -6.3 (-15.59, 3.05) | 0.6 (-4.79, 5.98) | 1.72 (-1.62, 5.1) | / |
| -1.75 (-3.39, -0.34) | -0.97 (-3.22, 1.01) | eTRE | -4.92 (-13.68, 3.9) | 2 (-2.6, 6.46) | 3.1 (-0.09, 6.31) | / |
| -5.66 (-9.82, -1.72) | -4.88 (-9.33, -0.71) | -3.92 (-7.68, -0.13) | eTRE + EX | 6.91 (-3.02, 16.75) | 8.02 (-1.34, 17.37) | / |
| -2.6 (-5.37, -0.08) | -1.82 (-5.24, 1.26) | -0.85 (-3.55, 1.8) | 3.07 (-1.58, 7.63) | lTRE | 1.13 (-4.11, 6.36) | / |
| -0.72 (-2.41, 0.98) | 0.06 (-1.86, 1.92) | 1.03 (-0.72, 3.01) | 4.93 (0.86, 9.23) | 1.88 (-1, 5.03) | mTRE | / |
| 0.99 (-3.78, 5.71) | 1.79 (-3.47, 6.92) | 2.75 (-2.19, 7.78) | 6.64 (0.5, 12.97) | 3.59 (-1.71, 9.11) | 1.72 (-3.36, 6.7) | mTRE + EX |

**Footnotes:** CR = calorie restriction, eTRE = early time-restrcited eating, mTRE = midday time-restricted eating, lTRE = late time-restricted eating, eTRE + EX = early time-restricted eating + exercise, mTRE + EX = midday time-restricted eating + exercise.

**Supplementary Table 35:** Sensitivity analysis presenting mean differences with 95% CI in FBG (mg/dl) (top) and FBI (mIU/L) (bottom) between different dietary approaches, after exclusion of studies including healthy subjects and keeping studies with chronically ill subjects.

| Control | -8.54 (-15.11, -2.77) | -9.8 (-14.95, -5.17) | -4.42 (-14.22, 5.47) | -7.02 (-13.64, -1.16) |
| --- | --- | --- | --- | --- |
| -0.06 (-3.04, 3.01) | CR | -1.27 (-4.89, 2.68) | 4.14 (-7.17, 16.14) | 1.51 (-2.35, 5.5) |
| -0.35 (-2.67, 1.55) | -0.35 (-2.92, 1.93) | eTRE | 5.39 (-5.39, 16.63) | 2.79 (-1.34, 6.69) |
| / | / | / | lTRE | -2.6 (-14.58, 8.73) |
| 0.27 (-2.6, 2.85) | 0.33 (-2.05, 2.36) | 0.67 (-1.53, 2.88) | / | mTRE |

**Footnotes:** CR = calorie restriction, eTRE = early time-restrcited eating, mTRE = midday time-restricted eating, lTRE = late time-restricted eating.

**Supplementary Table 36:** Sensitivity analysis presenting mean differences with 95% CI in HOMA-IR between different dietary approaches, after exclusion of studies including subjects with normal BMI and keeping studies with overweight and obese subjects.

| Control | -0.39 (-0.85, 0.1) | -0.63 (-1.01, -0.28) | -1.52 (-2.47, -0.61) | -0.59 (-1.3, 0.01) | -0.26 (-0.64, 0.13) |
| --- | --- | --- | --- | --- | --- |
| / | CR | -0.24 (-0.69, 0.16) | -1.13 (-2.12, -0.22) | -0.2 (-1.04, 0.51) | 0.13 (-0.3, 0.54) |
| / | / | eTRE | -0.9 (-1.75, -0.05) | 0.04 (-0.68, 0.68) | 0.37 (-0.02, 0.8) |
| / | / | / | eTRE + EX | 0.94 (-0.19, 1.99) | 1.26 (0.34, 2.24) |
| / | / | / | / | lTRE | 0.33 (-0.35, 1.12) |
| / | / | / | / | / | mTRE |

**Footnotes:** CR = calorie restriction, eTRE = early time-restrcited eating, mTRE = midday time-restricted eating, lTRE = late time-restricted eating, eTRE + EX = early time-restricted eating + exercise.

**Supplementary Table 37:** Sensitivity analysis presenting mean differences with 95% CI in HOMA-IR (top) and HbA1c (%) (bottom) between different dietary approaches, after exclusion of studies including healthy subjects and keeping studies with chronically ill subjects.

| Control | -0.3 (-1.3, 0.68) | -0.46 (-1.34, 0.27) | / | / | -0.32 (-1.25, 0.5) |
| --- | --- | --- | --- | --- | --- |
| -0.67 (-1.3, -0.04) | CR | -0.18 (-0.87, 0.44) | / | / | -0.02 (-0.87, 0.72) |
| -0.77 (-1.34, -0.19) | -0.1 (-0.62, 0.41) | eTRE | / | / | 0.16 (-0.51, 0.82) |
| / | / | / | eTRE + EX | / | / |
| -0.32 (-1.15, 0.53) | 0.35 (-0.7, 1.4) | 0.45 (-0.56, 1.47) | / | lTRE | / |
| -0.45 (-1.01, 0.12) | 0.21 (-0.33, 0.77) | 0.32 (-0.17, 0.81) | / | -0.13 (-1.14, 0.88) | mTRE |

**Footnotes:** CR = calorie restriction, eTRE = early time-restrcited eating, mTRE = midday time-restricted eating, lTRE = late time-restricted eating, eTRE + EX = early time-restricted eating + exercise.

**Supplementary Table 38:** Sensitivity analysis presenting mean differences with 95% CI in HbA1c (%) between different dietary approaches, after exclusion of studies including prescribed energy intake and keeping studies with *ad libitum* intake.

| Control | -0.3 (-0.82, 0.17) | -0.37 (-0.69, -0.09) | -0.25 (-0.6, 0.09) | -0.18 (-0.49, 0.11) | 0.4 (-0.31, 1.11) |
| --- | --- | --- | --- | --- | --- |
| / | CR | -0.07 (-0.61, 0.48) | 0.05 (-0.52, 0.66) | 0.12 (-0.36, 0.63) | 0.7 (-0.14, 1.59) |
| / | / | eTRE | 0.12 (-0.24, 0.51) | 0.19 (-0.15, 0.55) | 0.77 (0.01, 1.55) |
| / | / | / | lTRE | 0.07 (-0.37, 0.49) | 0.65 (-0.14, 1.45) |
| / | / | / | / | mTRE | 0.58 (-0.19, 1.36) |
| / | / | / | / | / | mTRE + EX |

**Footnotes:** CR = calorie restriction, eTRE = early time-restrcited eating, mTRE = midday time-restricted eating, lTRE = late time-restricted eating, mTRE + EX = midday time-restricted eating + exercise.

**Supplementary Table 39:** Sensitivity analysis presenting mean differences with 95% CI in TC (mg/dl) (top) and HDL-C (mg/dl) (bottom) between different dietary approaches, after exclusion of studies including subjects with normal BMI and keeping studies with overweight and obese subjects.

| Control | -1.71 (-8.24, 4.88) | 0.16 (-4.96, 5.63) | -28.3 (-40.08, -16.54) | -2.39 (-10.84, 6.67) | 1.05 (-4.3, 6.58) | -4.27 (-17.71, 9.26) |
| --- | --- | --- | --- | --- | --- | --- |
| 0.06 (-1.63, 1.92) | CR | 1.91 (-4.13, 8.1) | -26.58 (-39.2, -13.96) | -0.68 (-9.99, 9.23) | 2.78 (-3.53, 9.25) | -2.57 (-17.11, 11.96) |
| 0.44 (-0.63, 1.71) | 0.38 (-1.21, 1.96) | eTRE | -28.48 (-40.43, -16.72) | -2.56 (-11.4, 6.57) | 0.88 (-5.2, 6.82) | -4.44 (-18.74, 9.71) |
| 19.79 (12.5, 27.23) | 19.75 (12.32, 27.21) | 19.34 (12.09, 26.64) | eTRE + EX | 25.88 (11.9, 40.41) | 29.35 (17.46, 41.3) | 24.03 (7.5, 40.54) |
| 0.27 (-1.96, 2.63) | 0.22 (-2.55, 2.87) | -0.16 (-2.47, 2.06) | -19.53 (-27.12, -11.93) | lTRE | 3.45 (-6.48, 12.9) | -1.87 (-18.05, 13.86) |
| 1.32 (-0.09, 2.72) | 1.26 (-0.67, 3.02) | 0.87 (-0.75, 2.32) | -18.5 (-26.01, -11.14) | 1.03 (-1.61, 3.59) | mTRE | -5.34 (-19.34, 8.44) |
| -0.85 (-7.8, 6.11) | -0.92 (-8.15, 6.24) | -1.32 (-8.42, 5.72) | -20.66 (-30.87, -10.45) | -1.15 (-8.45, 6.12) | -2.17 (-9.24, 4.94) | mTRE + EX |

**Footnotes:** CR = calorie restriction, eTRE = early time-restrcited eating, mTRE = midday time-restricted eating, lTRE = late time-restricted eating, eTRE + EX = early time-restricted eating + exercise, mTRE + EX = midday time-restricted eating + exercise.

**Supplementary Table 40:** Sensitivity analysis presenting mean differences with 95% CI in TC (mg/dl) (top) and HDL-C (mg/dl) (bottom) between different dietary approaches, after exclusion of studies including healthy subjects and keeping studies with chronically ill subjects.

| Control | -7.86 (-18.07, 1.79) | -6.21 (-14.38, 0.76) | 8.86 (-7.84, 25.92) | -1.31 (-9.59, 7.41) |
| --- | --- | --- | --- | --- |
| -0.4 (-2.76, 2.18) | CR | 1.6 (-6.71, 9.42) | 16.77 (-2.54, 36.72) | 6.57 (-2.67, 16.49) |
| 0.18 (-1.13, 2.01) | 0.61 (-1.5, 2.79) | eTRE | 15.19 (-2.81, 34.13) | 4.98 (-2.29, 13.54) |
| / | / | / | lTRE | -10.18 (-28.98, 8.74) |
| -2.21 (-4.27, -0.25) | -2.6 (-5.24, 0.13) | -1.99 (-3.85, 0.1) | / | mTRE |

**Footnotes:** CR = calorie restriction, eTRE = early time-restrcited eating, mTRE = midday time-restricted eating, lTRE = late time-restricted eating.

**Supplementary Table 41:** Sensitivity analysis presenting mean differences with 95% CI in LDL-C (mg/dl) between different dietary approaches, after exclusion of studies including healthy subjects and keeping studies with chronically ill subjects.

| Control | -3.17 (-10.77, 5.49) | -2.79 (-8.29, 3.65) | 8.47 (-5.37, 22.35) | 6.77 (-0.11, 14.33) |
| --- | --- | --- | --- | --- |
| / | CR | 0.41 (-6.29, 6.63) | 11.67 (-4.95, 27.03) | 9.91 (1.98, 17.56) |
| / | / | eTRE | 11.25 (-4.18, 25.93) | 9.5 (3.04, 16.04) |
| / | / | / | lTRE | -1.77 (-16.85, 14.27) |
| / | / | / | / | mTRE |

**Footnotes:** CR = calorie restriction, eTRE = early time-restrcited eating, mTRE = midday time-restricted eating, lTRE = late time-restricted eating.

**Supplementary Table 42:** Sensitivity analysis presenting mean differences with 95% CI in SBP (mmHg) (top) and DBP (mmHg) (bottom) between different dietary approaches, after exclusion of studies including prescribed energy intake and keeping studies with *ad libitum* intake.

| Control | -4.79 (-10.19, 0.53) | -2.08 (-5.16, 0.89) | -1.77 (-10.95, 7.45) | -0.9 (-5.52, 3.61) | -0.88 (-3.62, 1.68) | -4.41 (-9.47, 0.54) |
| --- | --- | --- | --- | --- | --- | --- |
| -1.07 (-4.31, 2.09) | CR | 2.7 (-3.19, 8.59) | 3.05 (-7.48, 13.65) | 3.89 (-3.05, 10.85) | 3.9 (-1.34, 9.04) | 0.36 (-6.76, 7.58) |
| -0.6 (-2.46, 1.16) | 0.44 (-3.06, 4.06) | eTRE | 0.32 (-8.85, 9.59) | 1.19 (-3.7, 5.95) | 1.2 (-2.31, 4.62) | -2.33 (-8.08, 3.41) |
| -1.74 (-7.54, 3.98) | -0.72 (-7.19, 5.93) | -1.13 (-6.85, 4.56) | eTRE + EX | 0.88 (-9.37, 10.95) | 0.87 (-8.67, 10.27) | -2.64 (-13.14, 7.73) |
| -0.91 (-3.84, 1.72) | 0.14 (-4.15, 4.32) | -0.31 (-3.39, 2.56) | 0.86 (-5.53, 7.03) | lTRE | 0.01 (-5.11, 5.11) | -3.52 (-10.22, 3.26) |
| -0.91 (-2.57, 0.58) | 0.15 (-3.05, 3.31) | -0.29 (-2.42, 1.71) | 0.83 (-5.1, 6.69) | 0 (-3.01, 3.18) | mTRE | -3.54 (-8.75, 1.84) |
| 0.48 (-2.63, 3.5) | 1.53 (-2.8, 5.9) | 1.07 (-2.44, 4.58) | 2.21 (-4.28, 8.71) | 1.38 (-2.62, 5.61) | 1.36 (-1.84, 4.68) | mTRE + EX |

**Footnotes:** CR = calorie restriction, eTRE = early time-restrcited eating, mTRE = midday time-restricted eating, lTRE = late time-restricted eating, eTRE + EX = early time-restricted eating + exercise, mTRE + EX = midday time-restricted eating + exercise.

**Supplementary Table 43:** Sensitivity analysis presenting mean differences with 95% CI in SBP (mmHg) (top) and DBP (mmHg) (bottom) between different dietary approaches, after exclusion of studies including subjects with normal BMI and keeping studies with overweight and obese subjects.

| Control | -1.92 (-5.73, 1.68) | -2.66 (-5.65, 0.21) | -2.07 (-11.25, 6.94) | -1.07 (-5.57, 3.23) | -0.57 (-3.51, 2.22) | -4.02 (-11.64, 3.65) |
| --- | --- | --- | --- | --- | --- | --- |
| 1.56 (-0.85, 3.85) | CR | -0.74 (-3.96, 2.6) | -0.13 (-9.76, 9.4) | 0.85 (-4.54, 6.25) | 1.35 (-2.22, 5.04) | -2.11 (-10.46, 6.49) |
| -0.85 (-2.83, 1.06) | -2.42 (-4.46, -0.25) | eTRE | 0.61 (-8.58, 9.59) | 1.59 (-3.14, 6.26) | 2.09 (-1.11, 5.29) | -1.37 (-9.47, 6.91) |
| -1.88 (-8.02, 4.29) | -3.44 (-9.77, 3) | -1.03 (-7.13, 5.12) | eTRE + EX | 1 (-8.94, 10.94) | 1.48 (-7.8, 10.93) | -1.96 (-13.81, 10.04) |
| -1.13 (-4.22, 1.89) | -2.69 (-6.29, 0.98) | -0.27 (-3.56, 2.97) | 0.76 (-6.07, 7.47) | lTRE | 0.5 (-4.51, 5.59) | -2.96 (-11.65, 6) |
| -0.7 (-2.66, 1.18) | -2.27 (-4.42, -0.01) | 0.15 (-1.95, 2.25) | 1.17 (-5.17, 7.45) | 0.43 (-3.02, 3.92) | mTRE | -3.45 (-11.53, 4.77) |
| 2.97 (-2.18, 8.19) | 1.41 (-4.16, 7.21) | 3.82 (-1.67, 9.41) | 4.88 (-3.14, 12.92) | 4.1 (-1.86, 10.17) | 3.67 (-1.78, 9.27) | mTRE + EX |

**Footnotes:** CR = calorie restriction, eTRE = early time-restrcited eating, mTRE = midday time-restricted eating, lTRE = late time-restricted eating, eTRE + EX = early time-restricted eating + exercise, mTRE + EX = midday time-restricted eating + exercise.

**Supplementary Table 44:** Sensitivity analysis presenting mean differences with 95% CI in SBP (mmHg) (top) and DBP (mmHg) (bottom) between different dietary approaches, after exclusion of studies including healthy subjects and keeping studies with chronically ill subjects.

| Control | -0.74 (-7.25, 4.54) | -2.56 (-9.24, 2.82) | -1.02 (-9.72, 7.65) | 0.1 (-5.09, 3.86) |
| --- | --- | --- | --- | --- |
| -0.29 (-3.85, 3.09) | CR | -1.91 (-6.06, 2.44) | -0.23 (-10.26, 10.44) | 0.77 (-3.83, 5.3) |
| -2.7 (-6.52, 0.93) | -2.45 (-5.19, 0.35) | eTRE | 1.65 (-8.58, 12.43) | 2.68 (-1.85, 7.11) |
| -0.91 (-6.46, 4.59) | -0.66 (-7.15, 6.01) | 1.78 (-4.86, 8.53) | lTRE | 1.02 (-8.95, 10.32) |
| -0.86 (-3.88, 1.66) | -0.6 (-3.58, 2.27) | 1.83 (-1.17, 4.69) | 0.05 (-6.35, 6.22) | mTRE |

**Footnotes:** CR = calorie restriction, eTRE = early time-restrcited eating, mTRE = midday time-restricted eating, lTRE = late time-restricted eating.

**Supplementary Table 45:** Confidence in effect estimates in network meta-analysis of diet interventions for BW outcome.

| **Comparison** | **Number of studies** | **Within-study bias** | **Reporting bias** | **Indirectness** | **Imprecision** | **Heterogeneity** | **Incoherence** | **Confidence rating** |
| --- | --- | --- | --- | --- | --- | --- | --- | --- |
| **Control:CR** | 2 | No concerns | Low risk | No concerns | No concerns | No concerns | No concerns | High |
| **CR:eTRE** | 6 | No concerns | Low risk | No concerns | Some concerns | Some concerns | No concerns | Moderate |
| **CR:lTRE** | 1 | No concerns | Low risk | No concerns | Some concerns | Some concerns | Major concerns | Very low |
| **CR:mTRE** | 7 | No concerns | Low risk | No concerns | No concerns | Major concerns | No concerns | Moderate |
| **Control:eTRE** | 11 | No concerns | Low risk | No concerns | No concerns | No concerns | No concerns | High |
| **Control:eTRE + EX** | 1 | Some concerns | Low risk | No concerns | No concerns | No concerns | No concerns | Moderate |
| **Control:lTRE** | 2 | No concerns | Low risk | No concerns | No concerns | No concerns | Some concerns | Moderate |
| **Control:mTRE** | 15 | No concerns | Low risk | No concerns | No concerns | No concerns | No concerns | High |
| **Control:mTRE + EX** | 2 | Some concerns | Low risk | No concerns | No concerns | Some concerns | No concerns | Moderate |
| **eTRE:eTRE + EX** | 2 | Some concerns | Low risk | No concerns | Some concerns | No concerns | No concerns | Moderate |
| **eTRE:lTRE** | 2 | No concerns | Low risk | No concerns | No concerns | Major concerns | No concerns | Moderate |
| **eTRE:mTRE** | 9 | No concerns | Low risk | No concerns | No concerns | Some concerns | No concerns | Moderate |
| **mTRE:mTRE + EX** | 1 | Some concerns | Low risk | No concerns | Major concerns | No concerns | No concerns | Low |
| **CR:eTRE + EX** | 0 | Some concerns | Low risk | No concerns | Some concerns | No concerns | No concerns | Moderate |
| **CR:mTRE + EX** | 0 | Some concerns | Low risk | No concerns | Major concerns | No concerns | No concerns | Low |
| **eTRE:mTRE + EX** | 0 | Some concerns | Low risk | No concerns | Major concerns | No concerns | No concerns | Low |
| **eTRE + EX:lTRE** | 0 | Some concerns | Low risk | No concerns | Some concerns | No concerns | No concerns | Moderate |
| **eTRE + EX:mTRE** | 0 | Some concerns | Low risk | No concerns | Some concerns | No concerns | No concerns | Moderate |
| **eTRE + EX:mTRE + EX** | 0 | Some concerns | Low risk | No concerns | Some concerns | Some concerns | No concerns | Low |
| **lTRE:mTRE** | 0 | No concerns | Low risk | No concerns | Some concerns | Some concerns | No concerns | Moderate |
| **lTRE:mTRE + EX** | 0 | Some concerns | Low risk | No concerns | Major concerns | No concerns | No concerns | Low |

**Footnotes:** CR = calorie restriction, eTRE = early time-restrcited eating, mTRE = midday time-restricted eating, lTRE = late time-restricted eating, eTRE + EX = early time-restricted eating + exercise, mTRE + EX = midday time-restricted eating + exercise.

**Supplementary Table 46:** Confidence in effect estimates in network meta-analysis of diet interventions for BMI outcome.

| **Comparison** | **Number of studies** | **Within-study bias** | **Reporting bias** | **Indirectness** | **Imprecision** | **Heterogeneity** | **Incoherence** | **Confidence rating** |
| --- | --- | --- | --- | --- | --- | --- | --- | --- |
| **Control:CR** | 3 | No concerns | Low risk | No concerns | No concerns | Major concerns | No concerns | Moderate |
| **CR:eTRE** | 5 | No concerns | Low risk | No concerns | Some concerns | Some concerns | No concerns | Moderate |
| **CR:lTRE** | 2 | Some concerns | Low risk | No concerns | Major concerns | No concerns | No concerns | Low |
| **CR:mTRE** | 7 | No concerns | Low risk | No concerns | Major concerns | No concerns | No concerns | Moderate |
| **Control:eTRE** | 7 | No concerns | Low risk | No concerns | No concerns | Some concerns | No concerns | High |
| **Control:eTRE + EX** | 1 | Some concerns | Low risk | No concerns | No concerns | No concerns | No concerns | High |
| **Control:lTRE** | 2 | Some concerns | Low risk | No concerns | No concerns | Some concerns | Some concerns | Moderate |
| **Control:mTRE** | 11 | Some concerns | Low risk | No concerns | No concerns | Some concerns | No concerns | Moderate |
| **Control:mTRE + EX** | 2 | Some concerns | Low risk | No concerns | No concerns | Major concerns | No concerns | Low |
| **eTRE:eTRE + EX** | 2 | Some concerns | Low risk | No concerns | Major concerns | No concerns | No concerns | Low |
| **eTRE:lTRE** | 1 | No concerns | Low risk | No concerns | Major concerns | No concerns | Some concerns | Low |
| **eTRE:mTRE** | 6 | No concerns | Low risk | No concerns | Major concerns | No concerns | No concerns | Moderate |
| **mTRE:mTRE + EX** | 1 | Some concerns | Low risk | No concerns | Major concerns | No concerns | No concerns | Low |
| **CR:eTRE + EX** | 0 | Some concerns | Low risk | No concerns | Major concerns | No concerns | No concerns | Low |
| **CR:mTRE + EX** | 0 | Some concerns | Low risk | No concerns | Major concerns | No concerns | No concerns | Low |
| **eTRE:mTRE + EX** | 0 | Some concerns | Low risk | No concerns | Major concerns | No concerns | No concerns | Low |
| **eTRE + EX:lTRE** | 0 | Some concerns | Low risk | No concerns | Major concerns | No concerns | No concerns | Low |
| **eTRE + EX:mTRE** | 0 | Some concerns | Low risk | No concerns | Major concerns | No concerns | No concerns | Low |
| **eTRE + EX:mTRE + EX** | 0 | Some concerns | Low risk | No concerns | Major concerns | No concerns | No concerns | Low |
| **lTRE:mTRE** | 0 | Some concerns | Low risk | No concerns | Major concerns | No concerns | No concerns | Low |
| **lTRE:mTRE + EX** | 0 | Some concerns | Low risk | No concerns | Major concerns | No concerns | No concerns | Low |

**Footnotes:** CR = calorie restriction, eTRE = early time-restrcited eating, mTRE = midday time-restricted eating, lTRE = late time-restricted eating, eTRE + EX = early time-restricted eating + exercise, mTRE + EX = midday time-restricted eating + exercise.

**Supplementary Table 47:** Confidence in effect estimates in network meta-analysis of diet interventions for fat % outcome.

| **Comparison** | **Number of studies** | **Within-study bias** | **Reporting bias** | **Indirectness** | **Imprecision** | **Heterogeneity** | **Incoherence** | **Confidence rating** |
| --- | --- | --- | --- | --- | --- | --- | --- | --- |
| CR:eTRE | 4 | No concerns | Low risk | No concerns | No concerns | No concerns | Some concerns | High |
| CR:mTRE | 5 | Some concerns | Low risk | No concerns | No concerns | No concerns | Some concerns | Moderate |
| Control:eTRE | 8 | No concerns | Low risk | No concerns | No concerns | Some concerns | Some concerns | Moderate |
| Control:lTRE | 1 | No concerns | Low risk | No concerns | Some concerns | No concerns | No concerns | High |
| Control:mTRE | 11 | Some concerns | Low risk | No concerns | No concerns | Some concerns | No concerns | Moderate |
| Control:mTRE + EX | 2 | Some concerns | Low risk | No concerns | Some concerns | No concerns | Some concerns | Low |
| eTRE:lTRE | 1 | No concerns | Low risk | No concerns | No concerns | Some concerns | No concerns | High |
| eTRE:mTRE | 7 | No concerns | Low risk | No concerns | No concerns | No concerns | No concerns | High |
| mTRE:mTRE + EX | 1 | Some concerns | Low risk | No concerns | No concerns | Some concerns | Major concerns | Very low |
| Control:CR | 0 | No concerns | Low risk | No concerns | No concerns | Some concerns | No concerns | High |
| CR:lTRE | 0 | No concerns | Low risk | No concerns | No concerns | Major concerns | No concerns | Low |
| CR:mTRE + EX | 0 | Some concerns | Low risk | No concerns | Some concerns | No concerns | No concerns | Moderate |
| eTRE:mTRE + EX | 0 | Some concerns | Low risk | No concerns | No concerns | Some concerns | No concerns | Moderate |
| lTRE:mTRE | 0 | No concerns | Low risk | No concerns | No concerns | Major concerns | No concerns | Low |
| lTRE:mTRE + EX | 0 | No concerns | Low risk | No concerns | Some concerns | No concerns | No concerns | High |

**Footnotes:** CR = calorie restriction, eTRE = early time-restrcited eating, mTRE = midday time-restricted eating, lTRE = late time-restricted eating, eTRE + EX = early time-restricted eating + exercise, mTRE + EX = midday time-restricted eating + exercise.

**Supplementary Table 48:** Confidence in effect estimates in network meta-analysis of diet interventions for fat mass (kg) outcome.

| **Comparison** | **Number of studies** | **Within-study bias** | **Reporting bias** | **Indirectness** | **Imprecision** | **Heterogeneity** | **Incoherence** | **Confidence rating** |
| --- | --- | --- | --- | --- | --- | --- | --- | --- |
| Control:CR | 2 | No concerns | Low risk | No concerns | No concerns | Some concerns | No concerns | Moderate |
| CR:eTRE | 4 | No concerns | Low risk | No concerns | No concerns | Major concerns | Major concerns | Very low |
| CR:mTRE | 5 | No concerns | Low risk | No concerns | No concerns | Major concerns | No concerns | Low |
| Control:eTRE | 8 | No concerns | Low risk | No concerns | No concerns | No concerns | Some concerns | Moderate |
| Control:eTRE + EX | 1 | Some concerns | Low risk | No concerns | No concerns | No concerns | No concerns | Moderate |
| Control:lTRE | 2 | No concerns | Low risk | No concerns | No concerns | Some concerns | No concerns | Moderate |
| Control:mTRE | 11 | No concerns | Low risk | No concerns | No concerns | No concerns | No concerns | High |
| Control:mTRE + EX | 2 | Some concerns | Low risk | No concerns | Some concerns | No concerns | No concerns | Moderate |
| eTRE:eTRE + EX | 2 | Some concerns | Low risk | No concerns | No concerns | No concerns | No concerns | Moderate |
| eTRE:lTRE | 2 | No concerns | Low risk | No concerns | No concerns | Major concerns | No concerns | Low |
| eTRE:mTRE | 7 | No concerns | Low risk | No concerns | No concerns | Major concerns | No concerns | Low |
| mTRE:mTRE + EX | 1 | Some concerns | Low risk | No concerns | Major concerns | No concerns | No concerns | Low |
| CR:eTRE + EX | 0 | No concerns | Low risk | No concerns | No concerns | No concerns | No concerns | High |
| CR:lTRE | 0 | No concerns | Low risk | No concerns | Major concerns | No concerns | No concerns | Low |
| CR:mTRE + EX | 0 | No concerns | Low risk | No concerns | Major concerns | No concerns | No concerns | Low |
| eTRE:mTRE + EX | 0 | No concerns | Low risk | No concerns | Major concerns | No concerns | No concerns | Low |
| eTRE + EX:lTRE | 0 | No concerns | Low risk | No concerns | No concerns | No concerns | No concerns | High |
| eTRE + EX:mTRE | 0 | Some concerns | Low risk | No concerns | No concerns | No concerns | No concerns | Moderate |
| eTRE + EX:mTRE + EX | 0 | Some concerns | Low risk | No concerns | Some concerns | No concerns | No concerns | Moderate |
| lTRE:mTRE | 0 | No concerns | Low risk | No concerns | Some concerns | Some concerns | No concerns | Moderate |
| lTRE:mTRE + EX | 0 | No concerns | Low risk | No concerns | Major concerns | No concerns | No concerns | Low |

**Footnotes:** CR = calorie restriction, eTRE = early time-restrcited eating, mTRE = midday time-restricted eating, lTRE = late time-restricted eating, eTRE + EX = early time-restricted eating + exercise, mTRE + EX = midday time-restricted eating + exercise.

**Supplementary Table 49:** Confidence in effect estimates in network meta-analysis of diet interventions for LBM (kg) outcome.

| **Comparison** | **Number of studies** | **Within-study bias** | **Reporting bias** | **Indirectness** | **Imprecision** | **Heterogeneity** | **Incoherence** | **Confidence rating** |
| --- | --- | --- | --- | --- | --- | --- | --- | --- |
| Control:CR | 2 | No concerns | Low risk | No concerns | No concerns | Some concerns | No concerns | Moderate |
| CR:eTRE | 2 | No concerns | Low risk | No concerns | Some concerns | No concerns | No concerns | Moderate |
| CR:mTRE | 2 | No concerns | Low risk | No concerns | No concerns | Some concerns | No concerns | Moderate |
| Control:eTRE | 4 | No concerns | Low risk | No concerns | No concerns | Some concerns | No concerns | Moderate |
| Control:lTRE | 2 | No concerns | Low risk | No concerns | No concerns | Some concerns | No concerns | Moderate |
| Control:mTRE | 9 | No concerns | Low risk | No concerns | No concerns | Some concerns | No concerns | Moderate |
| Control:mTRE + EX | 2 | Some concerns | Low risk | No concerns | Major concerns | No concerns | No concerns | Low |
| eTRE:lTRE | 2 | No concerns | Low risk | No concerns | No concerns | Major concerns | No concerns | Low |
| eTRE:mTRE | 2 | No concerns | Low risk | No concerns | No concerns | Some concerns | No concerns | Moderate |
| mTRE:mTRE + EX | 1 | Some concerns | Low risk | No concerns | Major concerns | No concerns | No concerns | Low |
| CR:lTRE | 0 | No concerns | Low risk | No concerns | Some concerns | No concerns | No concerns | Moderate |
| CR:mTRE + EX | 0 | No concerns | Low risk | No concerns | Major concerns | No concerns | No concerns | Low |
| eTRE:mTRE + EX | 0 | No concerns | Low risk | No concerns | Major concerns | No concerns | No concerns | Low |
| lTRE:mTRE | 0 | No concerns | Low risk | No concerns | Some concerns | No concerns | No concerns | Moderate |
| lTRE:mTRE + EX | 0 | No concerns | Low risk | No concerns | Major concerns | No concerns | No concerns | Low |

**Footnotes:** CR = calorie restriction, eTRE = early time-restrcited eating, mTRE = midday time-restricted eating, lTRE = late time-restricted eating, eTRE + EX = early time-restricted eating + exercise, mTRE + EX = midday time-restricted eating + exercise.

**Supplementary Table 50:** Confidence in effect estimates in network meta-analysis of diet interventions for WC (cm) outcome.

| **Comparison** | **Number of studies** | **Within-study bias** | **Reporting bias** | **Indirectness** | **Imprecision** | **Heterogeneity** | **Incoherence** | **Confidence rating** |
| --- | --- | --- | --- | --- | --- | --- | --- | --- |
| Control:CR | 2 | No concerns | Low risk | No concerns | No concerns | Some concerns | No concerns | Moderate |
| CR:eTRE | 6 | No concerns | Low risk | No concerns | No concerns | Major concerns | No concerns | Low |
| CR:lTRE | 1 | Some concerns | Low risk | No concerns | Some concerns | Some concerns | Some concerns | Low |
| CR:mTRE | 7 | No concerns | Low risk | No concerns | Some concerns | Some concerns | No concerns | Moderate |
| Control:eTRE | 5 | No concerns | Low risk | No concerns | No concerns | Some concerns | No concerns | Moderate |
| Control:eTRE + EX | 1 | Some concerns | Low risk | No concerns | No concerns | Some concerns | No concerns | Moderate |
| Control:lTRE | 1 | Some concerns | Low risk | No concerns | Some concerns | Some concerns | No concerns | Low |
| Control:mTRE | 7 | Some concerns | Low risk | No concerns | No concerns | Some concerns | No concerns | Moderate |
| Control:mTRE + EX | 1 | Some concerns | Low risk | No concerns | Major concerns | No concerns | No concerns | Low |
| eTRE:eTRE + EX | 2 | Some concerns | Low risk | No concerns | Some concerns | Some concerns | No concerns | Moderate |
| eTRE:lTRE | 1 | No concerns | Low risk | No concerns | Some concerns | Some concerns | No concerns | Moderate |
| eTRE:mTRE | 5 | No concerns | Low risk | No concerns | Some concerns | Some concerns | No concerns | Moderate |
| CR:eTRE + EX | 0 | Some concerns | Low risk | No concerns | Some concerns | Some concerns | No concerns | Low |
| CR:mTRE + EX | 0 | Some concerns | Low risk | No concerns | Major concerns | No concerns | No concerns | Low |
| eTRE:mTRE + EX | 0 | Some concerns | Low risk | No concerns | Major concerns | No concerns | No concerns | Low |
| eTRE + EX:lTRE | 0 | Some concerns | Low risk | No concerns | Some concerns | Some concerns | No concerns | Low |
| eTRE + EX:mTRE | 0 | Some concerns | Low risk | No concerns | Some concerns | No concerns | No concerns | Low |
| eTRE + EX:mTRE + EX | 0 | Some concerns | Low risk | No concerns | Major concerns | No concerns | No concerns | Low |
| lTRE:mTRE | 0 | Some concerns | Low risk | No concerns | Major concerns | No concerns | No concerns | Low |
| lTRE:mTRE + EX | 0 | Some concerns | Low risk | No concerns | Major concerns | No concerns | No concerns | Low |
| mTRE:mTRE + EX | 0 | Some concerns | Low risk | No concerns | Major concerns | No concerns | No concerns | Low |

**Footnotes:** CR = calorie restriction, eTRE = early time-restrcited eating, mTRE = midday time-restricted eating, lTRE = late time-restricted eating, eTRE + EX = early time-restricted eating + exercise, mTRE + EX = midday time-restricted eating + exercise.

**Supplementary Table 51:** Confidence in effect estimates in network meta-analysis of diet interventions for HC (cm) outcome.

| **Comparison** | **Number of studies** | **Within-study bias** | **Reporting bias** | **Indirectness** | **Imprecision** | **Heterogeneity** | **Incoherence** | **Confidence rating** |
| --- | --- | --- | --- | --- | --- | --- | --- | --- |
| CR:eTRE | 1 | No concerns | Low risk | No concerns | Major concerns | No concerns | No concerns | Low |
| CR:lTRE | 1 | Some concerns | Low risk | No concerns | Major concerns | No concerns | Major concerns | Very low |
| CR:mTRE | 2 | No concerns | Low risk | No concerns | Major concerns | No concerns | No concerns | Low |
| Control:eTRE | 3 | Some concerns | Low risk | No concerns | Major concerns | No concerns | No concerns | Low |
| Control:eTRE + EX | 1 | Some concerns | Low risk | No concerns | Major concerns | No concerns | Major concerns | Very low |
| Control:mTRE | 2 | No concerns | Low risk | No concerns | Major concerns | No concerns | No concerns | Low |
| Control:mTRE + EX | 1 | Some concerns | Low risk | No concerns | Major concerns | No concerns | Major concerns | Very low |
| eTRE:eTRE + EX | 2 | Some concerns | Low risk | No concerns | No concerns | Major concerns | No concerns | Low |
| eTRE:mTRE | 4 | No concerns | Low risk | No concerns | Major concerns | No concerns | No concerns | Low |
| Control:CR | 0 | No concerns | Low risk | No concerns | Major concerns | No concerns | Major concerns | Very low |
| CR:eTRE + EX | 0 | No concerns | Low risk | No concerns | Major concerns | No concerns | Major concerns | Very low |
| CR:mTRE + EX | 0 | No concerns | Low risk | No concerns | Major concerns | No concerns | Major concerns | Very low |
| Control:lTRE | 0 | No concerns | Low risk | No concerns | Major concerns | No concerns | Major concerns | Very low |
| eTRE:lTRE | 0 | No concerns | Low risk | No concerns | Major concerns | No concerns | Major concerns | Very low |
| eTRE:mTRE + EX | 0 | Some concerns | Low risk | No concerns | Major concerns | No concerns | Major concerns | Very low |
| eTRE + EX:lTRE | 0 | Some concerns | Low risk | No concerns | Major concerns | No concerns | Major concerns | Very low |
| eTRE + EX:mTRE | 0 | Some concerns | Low risk | No concerns | Some concerns | Some concerns | Major concerns | Very low |
| eTRE + EX:mTRE + EX | 0 | Some concerns | Low risk | No concerns | Major concerns | No concerns | Major concerns | Very low |
| lTRE:mTRE | 0 | No concerns | Low risk | No concerns | Major concerns | No concerns | Major concerns | Very low |
| lTRE:mTRE + EX | 0 | Some concerns | Low risk | No concerns | Major concerns | No concerns | Major concerns | Very low |
| mTRE:mTRE + EX | 0 | Some concerns | Low risk | No concerns | Major concerns | No concerns | Major concerns | Very low |

**Footnotes:** CR = calorie restriction, eTRE = early time-restrcited eating, mTRE = midday time-restricted eating, lTRE = late time-restricted eating, eTRE + EX = early time-restricted eating + exercise, mTRE + EX = midday time-restricted eating + exercise.

**Supplementary Table 52:** Confidence in effect estimates in network meta-analysis of diet interventions for FBG outcome.

| **Comparison** | **Number of studies** | **Within-study bias** | **Reporting bias** | **Indirectness** | **Imprecision** | **Heterogeneity** | **Incoherence** | **Confidence rating** |
| --- | --- | --- | --- | --- | --- | --- | --- | --- |
| Control:CR | 2 | No concerns | Low risk | No concerns | No concerns | Some concerns | No concerns | Moderate |
| CR:eTRE | 6 | No concerns | Low risk | No concerns | No concerns | Some concerns | No concerns | Moderate |
| CR:mTRE | 6 | No concerns | Low risk | No concerns | No concerns | Some concerns | No concerns | Moderate |
| Control:eTRE | 10 | No concerns | Low risk | No concerns | No concerns | Some concerns | No concerns | Moderate |
| Control:lTRE | 3 | No concerns | Low risk | No concerns | No concerns | Some concerns | No concerns | Moderate |
| Control:mTRE | 9 | No concerns | Low risk | No concerns | No concerns | Major concerns | No concerns | Low |
| eTRE:eTRE + EX | 1 | Some concerns | Low risk | No concerns | Some concerns | No concerns | No concerns | Moderate |
| eTRE:lTRE | 2 | No concerns | Low risk | No concerns | No concerns | Some concerns | No concerns | Moderate |
| eTRE:mTRE | 7 | No concerns | Low risk | No concerns | No concerns | Some concerns | No concerns | Moderate |
| CR:eTRE + EX | 0 | Some concerns | Low risk | No concerns | Some concerns | No concerns | No concerns | Moderate |
| CR:lTRE | 0 | No concerns | Low risk | No concerns | No concerns | Major concerns | No concerns | Low |
| Control:eTRE + EX | 0 | Some concerns | Low risk | No concerns | Some concerns | No concerns | No concerns | Moderate |
| eTRE + EX:lTRE | 0 | No concerns | Low risk | No concerns | Some concerns | No concerns | No concerns | Moderate |
| eTRE + EX:mTRE | 0 | No concerns | Low risk | No concerns | Some concerns | No concerns | No concerns | Moderate |
| lTRE:mTRE | 0 | No concerns | Low risk | No concerns | No concerns | Major concerns | No concerns | Low |

**Footnotes:** CR = calorie restriction, eTRE = early time-restrcited eating, mTRE = midday time-restricted eating, lTRE = late time-restricted eating, eTRE + EX = early time-restricted eating + exercise, mTRE + EX = midday time-restricted eating + exercise.

**Supplementary Table 53:** Confidence in effect estimates in network meta-analysis of diet interventions for FBI outcome.

| **Comparison** | **Number of studies** | **Within-study bias** | **Reporting bias** | **Indirectness** | **Imprecision** | **Heterogeneity** | **Incoherence** | **Confidence rating** |
| --- | --- | --- | --- | --- | --- | --- | --- | --- |
| Control:CR | 1 | No concerns | Low risk | No concerns | Some concerns | No concerns | No concerns | Moderate |
| CR:eTRE | 3 | No concerns | Low risk | No concerns | Some concerns | Some concerns | Major concerns | Low |
| CR:mTRE | 4 | Some concerns | Low risk | No concerns | Some concerns | No concerns | Major concerns | Low |
| Control:eTRE | 8 | No concerns | Low risk | No concerns | No concerns | Some concerns | Major concerns | Low |
| Control:lTRE | 2 | No concerns | Low risk | No concerns | No concerns | Some concerns | No concerns | Moderate |
| Control:mTRE | 7 | No concerns | Low risk | No concerns | Some concerns | No concerns | Major concerns | Low |
| Control:mTRE + EX | 1 | Some concerns | Low risk | No concerns | Major concerns | No concerns | Major concerns | Very low |
| eTRE:eTRE + EX | 1 | Some concerns | Low risk | No concerns | No concerns | No concerns | Major concerns | Low |
| eTRE:lTRE | 2 | No concerns | Low risk | No concerns | Some concerns | No concerns | No concerns | Moderate |
| eTRE:mTRE | 5 | No concerns | Low risk | No concerns | No concerns | Major concerns | Major concerns | Very low |
| CR:eTRE + EX | 0 | Some concerns | Low risk | No concerns | No concerns | No concerns | Major concerns | Low |
| CR:lTRE | 0 | No concerns | Low risk | No concerns | Some concerns | Some concerns | Major concerns | Low |
| CR:mTRE + EX | 0 | Some concerns | Low risk | No concerns | Major concerns | No concerns | Major concerns | Very low |
| Control:eTRE + EX | 0 | No concerns | Low risk | No concerns | No concerns | No concerns | Major concerns | Low |
| eTRE:mTRE + EX | 0 | No concerns | Low risk | No concerns | Major concerns | No concerns | Major concerns | Very low |
| eTRE + EX:lTRE | 0 | No concerns | Low risk | No concerns | No concerns | No concerns | Major concerns | Low |
| eTRE + EX:mTRE | 0 | No concerns | Low risk | No concerns | No concerns | No concerns | Major concerns | Low |
| eTRE + EX:mTRE + EX | 0 | Some concerns | Low risk | No concerns | No concerns | No concerns | Major concerns | Low |
| lTRE:mTRE | 0 | No concerns | Low risk | No concerns | Some concerns | No concerns | Major concerns | Low |
| lTRE:mTRE + EX | 0 | No concerns | Low risk | No concerns | Major concerns | No concerns | Major concerns | Very low |
| mTRE:mTRE + EX | 0 | No concerns | Low risk | No concerns | Major concerns | No concerns | Major concerns | Very low |

**Footnotes:** CR = calorie restriction, eTRE = early time-restrcited eating, mTRE = midday time-restricted eating, lTRE = late time-restricted eating, eTRE + EX = early time-restricted eating + exercise, mTRE + EX = midday time-restricted eating + exercise.

**Supplementary Table 54:** Confidence in effect estimates in network meta-analysis of diet interventions for HOMA-IR outcome.

| **Comparison** | **Number of studies** | **Within-study bias** | **Reporting bias** | **Indirectness** | **Imprecision** | **Heterogeneity** | **Incoherence** | **Confidence rating** |
| --- | --- | --- | --- | --- | --- | --- | --- | --- |
| Control:CR | 1 | No concerns | Low risk | No concerns | No concerns | Some concerns | No concerns | Moderate |
| CR:eTRE | 5 | No concerns | Low risk | No concerns | No concerns | Some concerns | No concerns | Moderate |
| CR:mTRE | 4 | No concerns | Low risk | No concerns | No concerns | Some concerns | No concerns | Moderate |
| Control:eTRE | 8 | No concerns | Low risk | No concerns | No concerns | Some concerns | Some concerns | Moderate |
| Control:lTRE | 2 | No concerns | Low risk | No concerns | Some concerns | No concerns | No concerns | Moderate |
| Control:mTRE | 8 | No concerns | Low risk | No concerns | No concerns | Some concerns | No concerns | Moderate |
| eTRE:eTRE + EX | 1 | Some concerns | Low risk | No concerns | Some concerns | No concerns | Some concerns | Low |
| eTRE:lTRE | 1 | No concerns | Low risk | No concerns | No concerns | Major concerns | No concerns | Low |
| eTRE:mTRE | 6 | No concerns | Low risk | No concerns | No concerns | Some concerns | No concerns | Moderate |
| CR:eTRE + EX | 0 | Some concerns | Low risk | No concerns | No concerns | Some concerns | Some concerns | Low |
| CR:lTRE | 0 | No concerns | Low risk | No concerns | Some concerns | No concerns | Some concerns | Moderate |
| Control:eTRE + EX | 0 | Some concerns | Low risk | No concerns | No concerns | Some concerns | Some concerns | Low |
| eTRE + EX:lTRE | 0 | No concerns | Low risk | No concerns | Some concerns | No concerns | Some concerns | Moderate |
| eTRE + EX:mTRE | 0 | No concerns | Low risk | No concerns | No concerns | Some concerns | Some concerns | Moderate |
| lTRE:mTRE | 0 | No concerns | Low risk | No concerns | Some concerns | No concerns | Some concerns | Moderate |

**Footnotes:** CR = calorie restriction, eTRE = early time-restrcited eating, mTRE = midday time-restricted eating, lTRE = late time-restricted eating, eTRE + EX = early time-restricted eating + exercise, mTRE + EX = midday time-restricted eating + exercise.

**Supplementary Table 55:** Confidence in effect estimates in network meta-analysis of diet interventions for HbA1c (%) outcome.

| **Comparison** | **Number of studies** | **Within-study bias** | **Reporting bias** | **Indirectness** | **Imprecision** | **Heterogeneity** | **Incoherence** | **Confidence rating** |
| --- | --- | --- | --- | --- | --- | --- | --- | --- |
| Control:CR | 2 | Some concerns | Low risk | No concerns | Some concerns | Some concerns | No concerns | Low |
| CR:eTRE | 3 | No concerns | Low risk | No concerns | Some concerns | Some concerns | No concerns | Moderate |
| CR:mTRE | 3 | Some concerns | Low risk | No concerns | Some concerns | Some concerns | No concerns | Low |
| Control:eTRE | 4 | No concerns | Low risk | No concerns | No concerns | Major concerns | No concerns | Low |
| Control:lTRE | 3 | No concerns | Low risk | No concerns | Some concerns | Some concerns | No concerns | Moderate |
| Control:mTRE | 5 | No concerns | Low risk | No concerns | Some concerns | Some concerns | No concerns | Moderate |
| Control:mTRE + EX | 1 | Some concerns | Low risk | No concerns | Some concerns | Some concerns | Major concerns | Very low |
| eTRE:lTRE | 2 | No concerns | Low risk | No concerns | Some concerns | Some concerns | No concerns | Moderate |
| eTRE:mTRE | 3 | No concerns | Low risk | No concerns | Some concerns | Some concerns | No concerns | Moderate |
| CR:lTRE | 0 | No concerns | Low risk | No concerns | Major concerns | No concerns | Major concerns | Very low |
| CR:mTRE + EX | 0 | Some concerns | Low risk | No concerns | Some concerns | Some concerns | Major concerns | Very low |
| eTRE:mTRE + EX | 0 | Some concerns | Low risk | No concerns | No concerns | Some concerns | Major concerns | Low |
| lTRE:mTRE | 0 | No concerns | Low risk | No concerns | Some concerns | Some concerns | Major concerns | Very low |
| lTRE:mTRE + EX | 0 | Some concerns | Low risk | No concerns | Some concerns | Some concerns | Major concerns | Very low |
| mTRE:mTRE + EX | 0 | Some concerns | Low risk | No concerns | Some concerns | Some concerns | Major concerns | Very low |

**Footnotes:** CR = calorie restriction, eTRE = early time-restrcited eating, mTRE = midday time-restricted eating, lTRE = late time-restricted eating, eTRE + EX = early time-restricted eating + exercise, mTRE + EX = midday time-restricted eating + exercise.

**Supplementary Table 56:** Confidence in effect estimates in network meta-analysis of diet interventions for TG outcome.

| **Comparison** | **Number of studies** | **Within-study bias** | **Reporting bias** | **Indirectness** | **Imprecision** | **Heterogeneity** | **Incoherence** | **Confidence rating** |
| --- | --- | --- | --- | --- | --- | --- | --- | --- |
| Control:CR | 2 | No concerns | Low risk | No concerns | Major concerns | No concerns | No concerns | Low |
| CR:eTRE | 5 | No concerns | Low risk | No concerns | Some concerns | Some concerns | No concerns | Moderate |
| CR:mTRE | 5 | Some concerns | Low risk | No concerns | Some concerns | Some concerns | No concerns | Low |
| Control:eTRE | 9 | No concerns | Low risk | No concerns | No concerns | Major concerns | No concerns | Low |
| Control:lTRE | 3 | No concerns | Low risk | No concerns | Some concerns | Some concerns | No concerns | Moderate |
| Control:mTRE | 11 | No concerns | Low risk | No concerns | Some concerns | Some concerns | No concerns | Moderate |
| Control:mTRE + EX | 1 | Some concerns | Low risk | No concerns | Major concerns | No concerns | Some concerns | Low |
| eTRE:eTRE + EX | 1 | Some concerns | Low risk | No concerns | No concerns | No concerns | No concerns | Moderate |
| eTRE:lTRE | 2 | No concerns | Low risk | No concerns | Major concerns | No concerns | No concerns | Low |
| eTRE:mTRE | 6 | No concerns | Low risk | No concerns | Some concerns | Some concerns | No concerns | Moderate |
| mTRE:mTRE + EX | 1 | Some concerns | Low risk | No concerns | Major concerns | No concerns | Major concerns | Very low |
| CR:eTRE + EX | 0 | Some concerns | Low risk | No concerns | No concerns | Some concerns | No concerns | Moderate |
| CR:lTRE | 0 | No concerns | Low risk | No concerns | Major concerns | No concerns | No concerns | Low |
| CR:mTRE + EX | 0 | Some concerns | Low risk | No concerns | Major concerns | No concerns | No concerns | Low |
| Control:eTRE + EX | 0 | Some concerns | Low risk | No concerns | No concerns | Some concerns | No concerns | Moderate |
| eTRE:mTRE + EX | 0 | Some concerns | Low risk | No concerns | Major concerns | No concerns | No concerns | Low |
| eTRE + EX:lTRE | 0 | No concerns | Low risk | No concerns | No concerns | Some concerns | No concerns | Moderate |
| eTRE + EX:mTRE | 0 | Some concerns | Low risk | No concerns | No concerns | Some concerns | No concerns | Moderate |
| eTRE + EX:mTRE + EX | 0 | Some concerns | Low risk | No concerns | Some concerns | Some concerns | No concerns | Low |
| lTRE:mTRE | 0 | No concerns | Low risk | No concerns | Major concerns | No concerns | No concerns | Low |
| lTRE:mTRE + EX | 0 | Some concerns | Low risk | No concerns | Major concerns | No concerns | No concerns | Low |

**Footnotes:** CR = calorie restriction, eTRE = early time-restrcited eating, mTRE = midday time-restricted eating, lTRE = late time-restricted eating, eTRE + EX = early time-restricted eating + exercise, mTRE + EX = midday time-restricted eating + exercise.

**Supplementary Table 57:** Confidence in effect estimates in network meta-analysis of diet interventions for TC outcome.

| **Comparison** | **Number of studies** | **Within-study bias** | **Reporting bias** | **Indirectness** | **Imprecision** | **Heterogeneity** | **Incoherence** | **Confidence rating** |
| --- | --- | --- | --- | --- | --- | --- | --- | --- |
| Control:CR | 3 | No concerns | Low risk | No concerns | Some concerns | Some concerns | Major concerns | Low |
| CR:eTRE | 5 | No concerns | Low risk | No concerns | Some concerns | Some concerns | No concerns | Moderate |
| CR:lTRE | 1 | No concerns | Low risk | No concerns | Some concerns | Some concerns | Some concerns | Low |
| CR:mTRE | 5 | Some concerns | Low risk | No concerns | Some concerns | No concerns | No concerns | Moderate |
| Control:eTRE | 9 | No concerns | Low risk | No concerns | No concerns | Major concerns | No concerns | Low |
| Control:eTRE+EX | 1 | Some concerns | Low risk | No concerns | No concerns | No concerns | No concerns | Moderate |
| Control:lTRE | 3 | No concerns | Low risk | No concerns | Some concerns | No concerns | No concerns | Moderate |
| Control:mTRE | 12 | Some concerns | Low risk | No concerns | No concerns | Major concerns | No concerns | Low |
| Control:mTRE+EX | 3 | Some concerns | Low risk | No concerns | Major concerns | No concerns | No concerns | Low |
| eTRE:eTRE+EX | 1 | Some concerns | Low risk | No concerns | No concerns | No concerns | No concerns | Moderate |
| eTRE:lTRE | 2 | No concerns | Low risk | No concerns | Some concerns | Some concerns | Some concerns | Low |
| eTRE:mTRE | 7 | No concerns | Low risk | No concerns | Some concerns | No concerns | No concerns | Moderate |
| eTRE+EX:mTRE | 1 | Some concerns | Low risk | No concerns | No concerns | No concerns | No concerns | Moderate |
| eTRE+EX:mTRE+EX | 1 | Major concerns | Low risk | No concerns | No concerns | No concerns | Some concerns | Low |
| mTRE:mTRE+EX | 2 | Some concerns | Low risk | No concerns | Major concerns | No concerns | No concerns | Low |
| CR:eTRE+EX | 0 | Some concerns | Low risk | No concerns | No concerns | No concerns | Major concerns | Low |
| CR:mTRE+EX | 0 | Some concerns | Low risk | No concerns | Major concerns | No concerns | Major concerns | Very low |
| eTRE:mTRE+EX | 0 | Some concerns | Low risk | No concerns | Major concerns | No concerns | Major concerns | Very low |
| eTRE+EX:lTRE | 0 | Some concerns | Low risk | No concerns | No concerns | No concerns | Major concerns | Low |
| lTRE:mTRE | 0 | No concerns | Low risk | No concerns | No concerns | Some concerns | Major concerns | Low |
| lTRE:mTRE+EX | 0 | Some concerns | Low risk | No concerns | Major concerns | No concerns | Major concerns | Very low |

**Footnotes:** CR = calorie restriction, eTRE = early time-restrcited eating, mTRE = midday time-restricted eating, lTRE = late time-restricted eating, eTRE + EX = early time-restricted eating + exercise, mTRE + EX = midday time-restricted eating + exercise.

**Supplementary Table 58:** Confidence in effect estimates in network meta-analysis of diet interventions for LDL-C outcome.

| **Comparison** | **Number of studies** | **Within-study bias** | **Reporting bias** | **Indirectness** | **Imprecision** | **Heterogeneity** | **Incoherence** | **Confidence rating** |
| --- | --- | --- | --- | --- | --- | --- | --- | --- |
| Control:CR | 3 | No concerns | Low risk | No concerns | No concerns | Some concerns | Major concerns | Low |
| CR:eTRE | 5 | No concerns | Low risk | No concerns | No concerns | Some concerns | No concerns | Moderate |
| CR:lTRE | 1 | No concerns | Low risk | No concerns | Major concerns | No concerns | Major concerns | Very low |
| CR:mTRE | 5 | Some concerns | Low risk | No concerns | Some concerns | No concerns | No concerns | Moderate |
| Control:eTRE | 9 | No concerns | Low risk | No concerns | No concerns | No concerns | No concerns | High |
| Control:eTRE + EX | 1 | Some concerns | Low risk | No concerns | No concerns | No concerns | No concerns | Moderate |
| Control:lTRE | 4 | No concerns | Low risk | No concerns | Some concerns | Some concerns | No concerns | Moderate |
| Control:mTRE | 12 | Some concerns | Low risk | No concerns | No concerns | Some concerns | No concerns | Moderate |
| Control:mTRE + EX | 2 | Some concerns | Low risk | No concerns | Some concerns | No concerns | Major concerns | Low |
| eTRE:eTRE + EX | 1 | Some concerns | Low risk | No concerns | No concerns | No concerns | No concerns | Moderate |
| eTRE:lTRE | 2 | No concerns | Low risk | No concerns | Major concerns | No concerns | Major concerns | Low |
| eTRE:mTRE | 7 | No concerns | Low risk | No concerns | No concerns | Some concerns | No concerns | Moderate |
| eTRE + EX:mTRE | 1 | Some concerns | Low risk | No concerns | No concerns | No concerns | No concerns | Moderate |
| eTRE + EX:mTRE + EX | 1 | Major concerns | Low risk | No concerns | No concerns | No concerns | No concerns | Low |
| mTRE:mTRE + EX | 2 | Some concerns | Low risk | No concerns | Some concerns | No concerns | No concerns | Moderate |
| CR:eTRE + EX | 0 | Some concerns | Low risk | No concerns | No concerns | No concerns | Major concerns | Low |
| CR:mTRE + EX | 0 | Some concerns | Low risk | No concerns | Some concerns | No concerns | Major concerns | Low |
| eTRE:mTRE + EX | 0 | Some concerns | Low risk | No concerns | Some concerns | No concerns | Major concerns | Low |
| eTRE + EX:lTRE | 0 | Some concerns | Low risk | No concerns | No concerns | No concerns | Major concerns | Low |
| lTRE:mTRE | 0 | No concerns | Low risk | No concerns | Some concerns | No concerns | Major concerns | Low |
| lTRE:mTRE + EX | 0 | Some concerns | Low risk | No concerns | Some concerns | No concerns | Major concerns | Low |

**Footnotes:** CR = calorie restriction, eTRE = early time-restrcited eating, mTRE = midday time-restricted eating, lTRE = late time-restricted eating, eTRE + EX = early time-restricted eating + exercise, mTRE + EX = midday time-restricted eating + exercise.

**Supplementary Table 59:** Confidence in effect estimates in network meta-analysis of diet interventions for HDL-C outcome.

| **Comparison** | **Number of studies** | **Within-study bias** | **Reporting bias** | **Indirectness** | **Imprecision** | **Heterogeneity** | **Incoherence** | **Confidence rating** |
| --- | --- | --- | --- | --- | --- | --- | --- | --- |
| Control:CR | 2 | No concerns | Low risk | No concerns | No concerns | No concerns | No concerns | High |
| CR:eTRE | 5 | No concerns | Low risk | No concerns | No concerns | No concerns | No concerns | High |
| CR:mTRE | 5 | No concerns | Low risk | No concerns | No concerns | No concerns | No concerns | High |
| Control:eTRE | 10 | No concerns | Low risk | No concerns | No concerns | No concerns | No concerns | High |
| Control:lTRE | 2 | No concerns | Low risk | No concerns | No concerns | No concerns | No concerns | High |
| Control:mTRE | 11 | No concerns | Low risk | No concerns | No concerns | No concerns | No concerns | High |
| Control:mTRE+EX | 2 | Some concerns | Low risk | No concerns | No concerns | No concerns | No concerns | Moderate |
| eTRE:eTRE+EX | 1 | Some concerns | Low risk | No concerns | No concerns | No concerns | No concerns | Moderate |
| eTRE:lTRE | 2 | No concerns | Low risk | No concerns | No concerns | No concerns | No concerns | High |
| eTRE:mTRE | 7 | No concerns | Low risk | No concerns | No concerns | No concerns | No concerns | High |
| mTRE:mTRE+EX | 1 | Some concerns | Low risk | No concerns | No concerns | No concerns | No concerns | Moderate |
| CR:eTRE+EX | 0 | Some concerns | Low risk | No concerns | No concerns | No concerns | No concerns | Moderate |
| CR:lTRE | 0 | No concerns | Low risk | No concerns | No concerns | No concerns | No concerns | High |
| CR:mTRE+EX | 0 | Some concerns | Low risk | No concerns | No concerns | No concerns | No concerns | Moderate |
| Control:eTRE+EX | 0 | Some concerns | Low risk | No concerns | No concerns | No concerns | No concerns | Moderate |
| eTRE:mTRE+EX | 0 | Some concerns | Low risk | No concerns | No concerns | No concerns | No concerns | Moderate |
| eTRE+EX:lTRE | 0 | No concerns | Low risk | No concerns | No concerns | No concerns | No concerns | High |
| eTRE+EX:mTRE | 0 | Some concerns | Low risk | No concerns | No concerns | No concerns | No concerns | Moderate |
| eTRE+EX:mTRE+EX | 0 | Some concerns | Low risk | No concerns | No concerns | No concerns | No concerns | Moderate |
| lTRE:mTRE | 0 | No concerns | Low risk | No concerns | No concerns | No concerns | No concerns | High |
| lTRE:mTRE+EX | 0 | No concerns | Low risk | No concerns | No concerns | No concerns | No concerns | High |

**Footnotes:** CR = calorie restriction, eTRE = early time-restrcited eating, mTRE = midday time-restricted eating, lTRE = late time-restricted eating, eTRE + EX = early time-restricted eating + exercise, mTRE + EX = midday time-restricted eating + exercise.

**Supplementary Table 60:** Confidence in effect estimates in network meta-analysis of diet interventions for SBP and DBP outcomes.

| **Comparison** | **Number of studies** | **Within-study bias** | **Reporting bias** | **Indirectness** | **Imprecision** | **Heterogeneity** | **Incoherence** | **Confidence rating** |
| --- | --- | --- | --- | --- | --- | --- | --- | --- |
| Control:CR | 2 | No concerns | Low risk | No concerns | Some concerns | Some concerns | No concerns | Moderate |
| CR:eTRE | 4 | No concerns | Low risk | No concerns | No concerns | Major concerns | No concerns | Low |
| CR:mTRE | 5 | No concerns | Low risk | No concerns | No concerns | Major concerns | No concerns | Low |
| Control:eTRE | 6 | No concerns | Low risk | No concerns | No concerns | Some  concerns | No concerns | Moderate |
| Control:eTRE + EX | 1 | Some concerns | Low risk | No concerns | Major concerns | No concerns | No concerns | Low |
| Control:lTRE | 3 | No concerns | Low risk | No concerns | Some concerns | Some  concerns | No concerns | Moderate |
| Control:mTRE | 9 | No concerns | Low risk | No concerns | No concerns | Major concerns | No concerns | Low |
| Control:mTRE + EX | 2 | Some concerns | Low risk | No concerns | No concerns | Some  concerns | No concerns | Moderate |
| eTRE:eTRE + EX | 1 | Some concerns | Low risk | No concerns | Major concerns | No concerns | No concerns | Low |
| eTRE:lTRE | 2 | No concerns | Low risk | No concerns | Some concerns | Some  concerns | No concerns | Moderate |
| eTRE:mTRE | 4 | No concerns | Low risk | No concerns | Some concerns | Some concerns | No concerns | Moderate |
| mTRE:mTRE + EX | 1 | Some concerns | Low risk | No concerns | Some concerns | No concerns | No concerns | Moderate |
| CR:eTRE + EX | 0 | Some concerns | Low risk | No concerns | Major concerns | No concerns | No concerns | Low |
| CR:lTRE | 0 | No concerns | Low risk | No concerns | Major concerns | No concerns | No concerns | Low |
| CR:mTRE + EX | 0 | Some concerns | Low risk | No concerns | Some concerns | Some concerns | No concerns | Moderate |
| eTRE:mTRE + EX | 0 | Some concerns | Low risk | No concerns | Some  concerns | Some  concerns | No concerns | Moderate |
| eTRE + EX:lTRE | 0 | Some concerns | Low risk | No concerns | Major concerns | No concerns | No concerns | Low |
| eTRE + EX:mTRE | 0 | Some concerns | Low risk | No concerns | Major concerns | No concerns | No concerns | Low |
| eTRE + EX:mTRE + EX | 0 | Some concerns | Low risk | No concerns | Major concerns | No concerns | No concerns | Low |
| lTRE:mTRE | 0 | No concerns | Low risk | No concerns | Major concerns | No concerns | No concerns | Low |
| lTRE:mTRE + EX | 0 | Some concerns | Low risk | No concerns | Some concerns | Some concerns | No concerns | Moderate |

**Footnotes:** CR = calorie restriction, eTRE = early time-restrcited eating, mTRE = midday time-restricted eating, lTRE = late time-restricted eating, eTRE + EX = early time-restricted eating + exercise, mTRE + EX = midday time-restricted eating + exercise.

**Supplementary Figure 1:** Risk of bias graph.

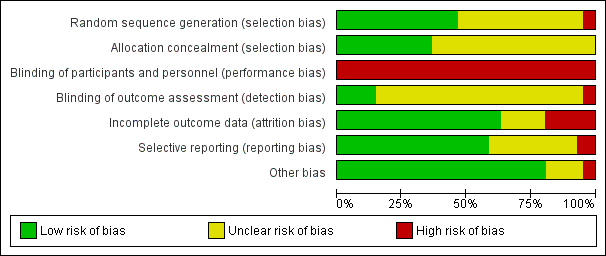

**Supplementary Figure 2:** Risk of bias summary.

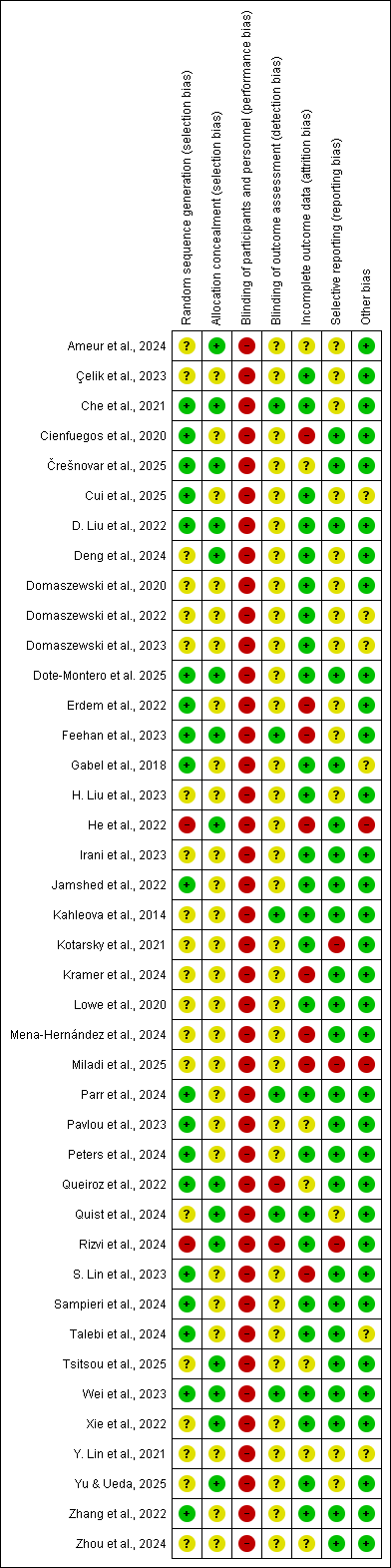

**Supplementary Figure 3:** Forest plots for network meta-analysis of BW.

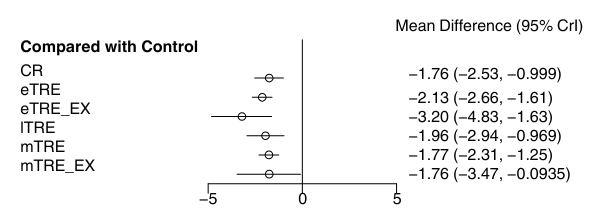
**Footnotes:** CR = calorie restriction, eTRE = early time-restrcited eating, mTRE = midday time-restricted eating, lTRE = late time-restricted eating, eTRE + EX = early time-restricted eating + exercise, mTRE + EX = midday time-restricted eating + exercise.

**Supplementary Figure 4:** Forest plots for network meta-analysis of BMI.

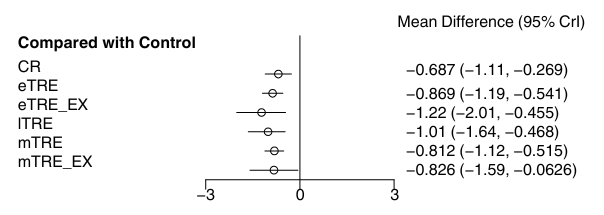

**Footnotes:** CR = calorie restriction, eTRE = early time-restrcited eating, mTRE = midday time-restricted eating, lTRE = late time-restricted eating, eTRE + EX = early time-restricted eating + exercise, mTRE + EX = midday time-restricted eating + exercise.

**Supplementary Figure 5:** Forest plots for network meta-analysis of FM (kg).

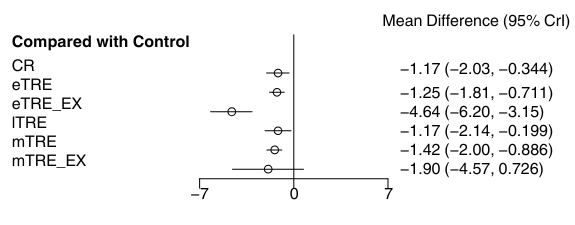

**Footnotes:** CR = calorie restriction, eTRE = early time-restrcited eating, mTRE = midday time-restricted eating, lTRE = late time-restricted eating, eTRE + EX = early time-restricted eating + exercise, mTRE + EX = midday time-restricted eating + exercise.

**Supplementary Figure 6:** Forest plots for network meta-analysis of FM (%).

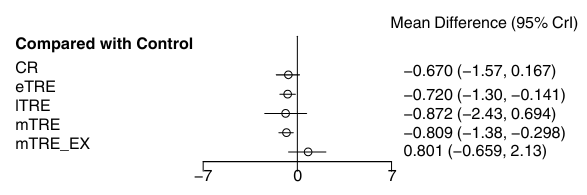

**Footnotes:** CR = calorie restriction, eTRE = early time-restrcited eating, mTRE = midday time-restricted eating, lTRE = late time-restricted eating, eTRE + EX = early time-restricted eating + exercise, mTRE + EX = midday time-restricted eating + exercise.

**Supplementary Figure 7:** Forest plots for network meta-analysis of LBM (kg).

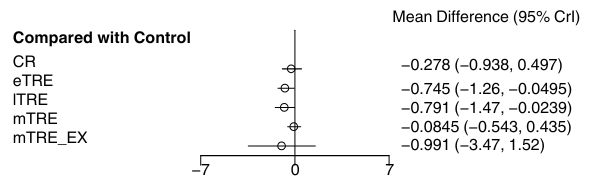

**Footnotes:** CR = calorie restriction, eTRE = early time-restrcited eating, mTRE = midday time-restricted eating, lTRE = late time-restricted eating, eTRE + EX = early time-restricted eating + exercise, mTRE + EX = midday time-restricted eating + exercise.

**Supplementary Figure 8:** Forest plots for network meta-analysis of WC (cm).

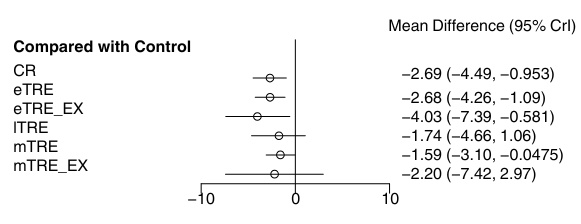

**Footnotes:** CR = calorie restriction, eTRE = early time-restrcited eating, mTRE = midday time-restricted eating, lTRE = late time-restricted eating, eTRE + EX = early time-restricted eating + exercise, mTRE + EX = midday time-restricted eating + exercise.

**Supplementary Figure 9:** Forest plots for network meta-analysis of HC (cm).

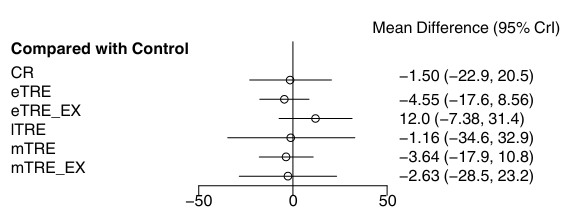

**Footnotes:** CR = calorie restriction, eTRE = early time-restrcited eating, mTRE = midday time-restricted eating, lTRE = late time-restricted eating, eTRE + EX = early time-restricted eating + exercise, mTRE + EX = midday time-restricted eating + exercise.

**Supplementary Figure 10:** Forest plots for network meta-analysis of WHR.

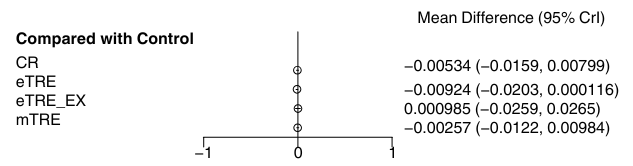

**Footnotes:** CR = calorie restriction, eTRE = early time-restrcited eating, mTRE = midday time-restricted eating, lTRE = late time-restricted eating, eTRE + EX = early time-restricted eating + exercise, mTRE + EX = midday time-restricted eating + exercise.

**Supplementary Figure 11:** Forest plots for network meta-analysis of FBG (mg/dl).

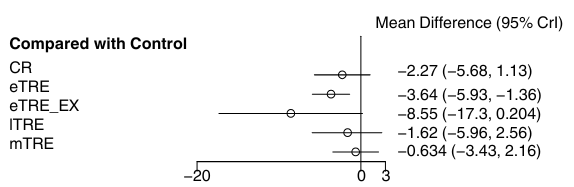

**Footnotes:** CR = calorie restriction, eTRE = early time-restrcited eating, mTRE = midday time-restricted eating, lTRE = late time-restricted eating, eTRE + EX = early time-restricted eating + exercise, mTRE + EX = midday time-restricted eating + exercise.

**Supplementary Figure 12:** Forest plots for network meta-analysis of FBI (mIU/L).

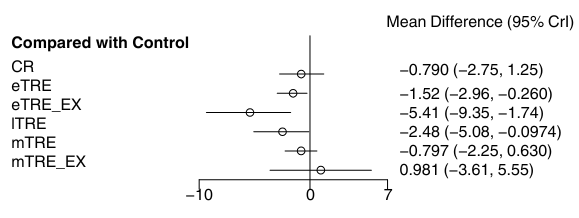

**Footnotes:** CR = calorie restriction, eTRE = early time-restrcited eating, mTRE = midday time-restricted eating, lTRE = late time-restricted eating, eTRE + EX = early time-restricted eating + exercise, mTRE + EX = midday time-restricted eating + exercise.

**Supplementary Figure 13:** Forest plots for network meta-analysis of HOMA-IR.

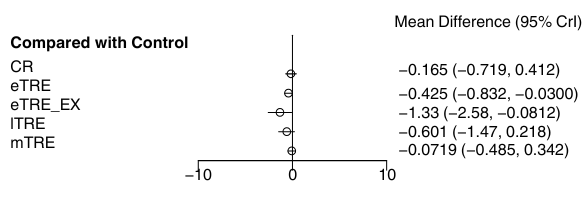

**Footnotes:** CR = calorie restriction, eTRE = early time-restrcited eating, mTRE = midday time-restricted eating, lTRE = late time-restricted eating, eTRE + EX = early time-restricted eating + exercise.

**Supplementary Figure 14:** Forest plots for network meta-analysis of HbA1c.

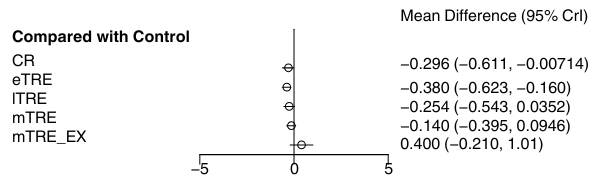

**Footnotes:** CR = calorie restriction, eTRE = early time-restrcited eating, mTRE = midday time-restricted eating, lTRE = late time-restricted eating, eTRE + EX = early time-restricted eating + exercise, mTRE + EX = midday time-restricted eating + exercise.

**Supplementary Figure 15:** Forest plots for network meta-analysis of TC (mg/dl).

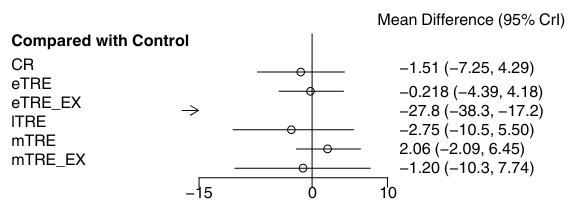
 **Footnotes:** CR = calorie restriction, eTRE = early time-restrcited eating, mTRE = midday time-restricted eating, lTRE = late time-restricted eating, eTRE + EX = early time-restricted eating + exercise, mTRE + EX = midday time-restricted eating + exercise.

**Supplementary Figure 16:** Forest plots for network meta-analysis of LDL-C (mg/dl).

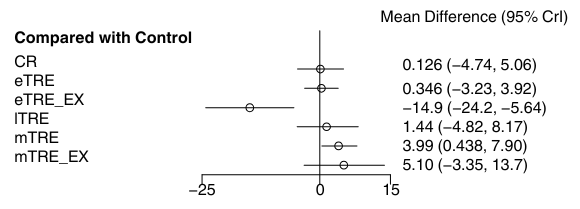

**Footnotes:** CR = calorie restriction, eTRE = early time-restrcited eating, mTRE = midday time-restricted eating, lTRE = late time-restricted eating, eTRE + EX = early time-restricted eating + exercise, mTRE + EX = midday time-restricted eating + exercise.

**Supplementary Figure 17:** Forest plots for network meta-analysis of HDL-C (mg/dl).

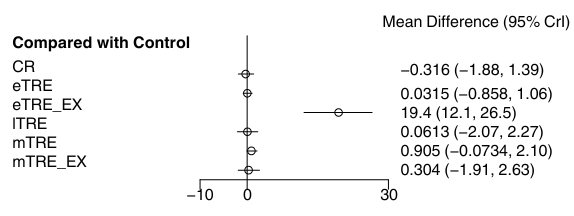

**Footnotes:** CR = calorie restriction, eTRE = early time-restrcited eating, mTRE = midday time-restricted eating, lTRE = late time-restricted eating, eTRE + EX = early time-restricted eating + exercise, mTRE + EX = midday time-restricted eating + exercise.

**Supplementary Figure 18:** Forest plots for network meta-analysis of TG (mmol/L).

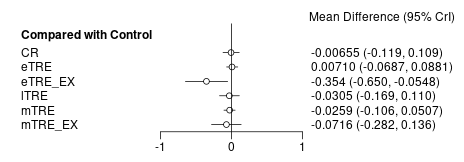

**Footnotes:** CR = calorie restriction, eTRE = early time-restrcited eating, mTRE = midday time-restricted eating, lTRE = late time-restricted eating, eTRE + EX = early time-restricted eating + exercise, mTRE + EX = midday time-restricted eating + exercise.

**Supplementary Figure 19:** Forest plots for network meta-analysis of SBP (mmHg).

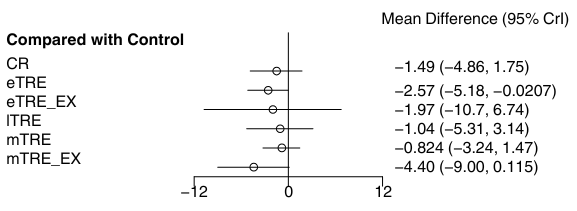

**Footnotes:** CR = calorie restriction, eTRE = early time-restrcited eating, mTRE = midday time-restricted eating, lTRE = late time-restricted eating, eTRE + EX = early time-restricted eating + exercise, mTRE + EX = midday time-restricted eating + exercise.

**Supplementary Figure 20:** Forest plots for network meta-analysis of DBP (mmHg).

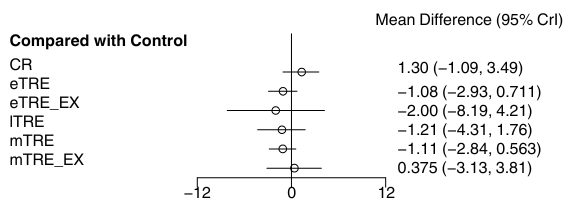

**Footnotes:** CR = calorie restriction, eTRE = early time-restrcited eating, mTRE = midday time-restricted eating, lTRE = late time-restricted eating, eTRE + EX = early time-restricted eating + exercise, mTRE + EX = midday time-restricted eating + exercise.

**Node-splitting approach:**

**Supplementary Figure 21:** Node-splitting approach to assess inconsistency for BW.

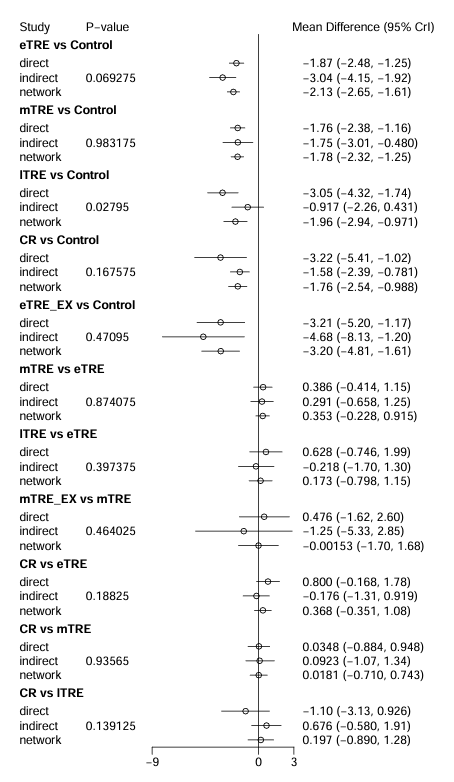

**Footnotes:** CR = calorie restriction, eTRE = early time-restrcited eating, mTRE = midday time-restricted eating, lTRE = late time-restricted eating, eTRE + EX = early time-restricted eating + exercise, mTRE + EX = midday time-restricted eating + exercise.

**Supplementary Figure 22:** Node-splitting approach to assess inconsistency for BMI.

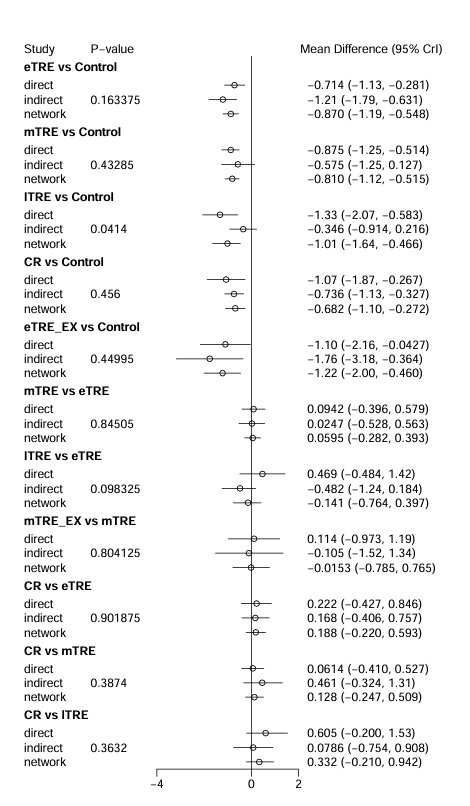

**Footnotes:** CR = calorie restriction, eTRE = early time-restrcited eating, mTRE = midday time-restricted eating, lTRE = late time-restricted eating, eTRE + EX = early time-restricted eating + exercise, mTRE + EX = midday time-restricted eating + exercise.

**Supplementary Figure 23:** Node-splitting approach to assess inconsistency for FM (kg).

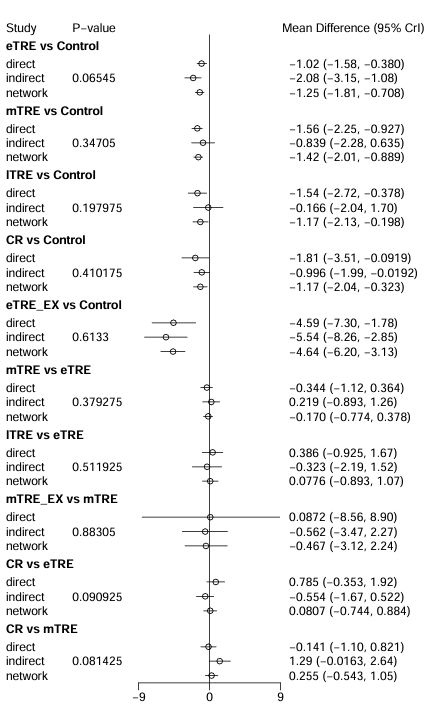

**Footnotes:** CR = calorie restriction, eTRE = early time-restrcited eating, mTRE = midday time-restricted eating, lTRE = late time-restricted eating, eTRE + EX = early time-restricted eating + exercise, mTRE + EX = midday time-restricted eating + exercise.

**Supplementary Figure 24:** Node-splitting approach to assess inconsistency for fat (%).

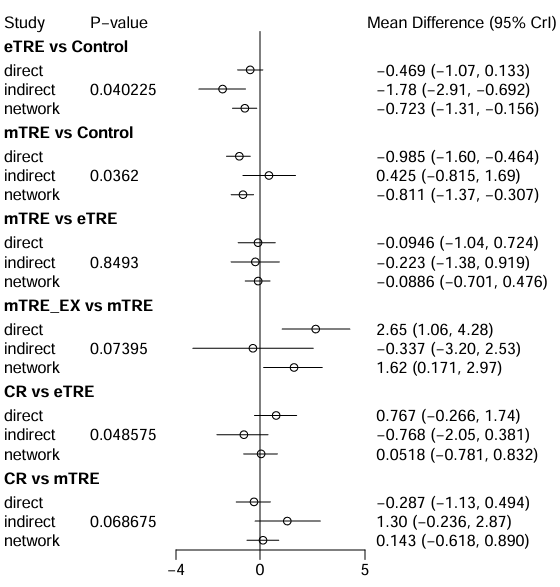
.

**Footnotes:** CR = calorie restriction, eTRE = early time-restrcited eating, mTRE = midday time-restricted eating, mTRE + EX = midday time-restricted eating + exercise.

**Supplementary Figure 25:** Node-splitting approach to assess inconsistency for LBM (kg).

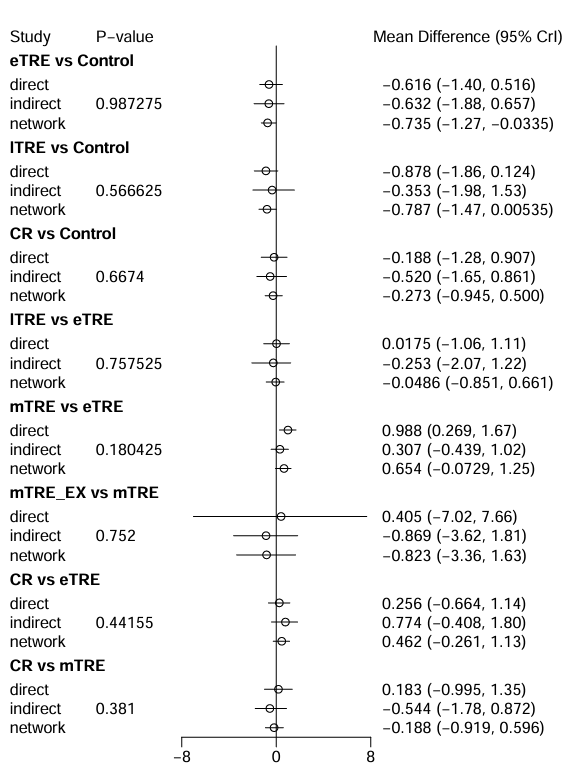

**Footnotes:** CR = calorie restriction, eTRE = early time-restrcited eating, mTRE = midday time-restricted eating, lTRE = late time-restricted eating, eTRE + EX = early time-restricted eating + exercise, mTRE + EX = midday time-restricted eating + exercise.

**Supplementary Figure 26:** Node-splitting approach to assess inconsistency for WC.

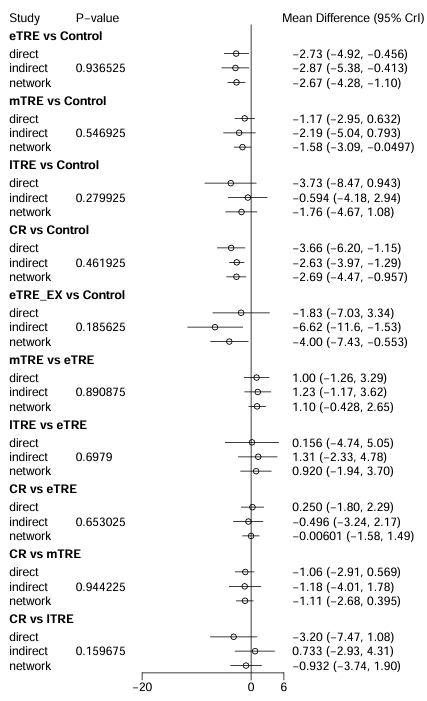

**Footnotes:** CR = calorie restriction, eTRE = early time-restrcited eating, mTRE = midday time-restricted eating, lTRE = late time-restricted eating, eTRE + EX = early time-restricted eating + exercise, mTRE + EX = midday time-restricted eating + exercise.

**Supplementary Figure 27:** Node-splitting approach to assess inconsistency for HC.

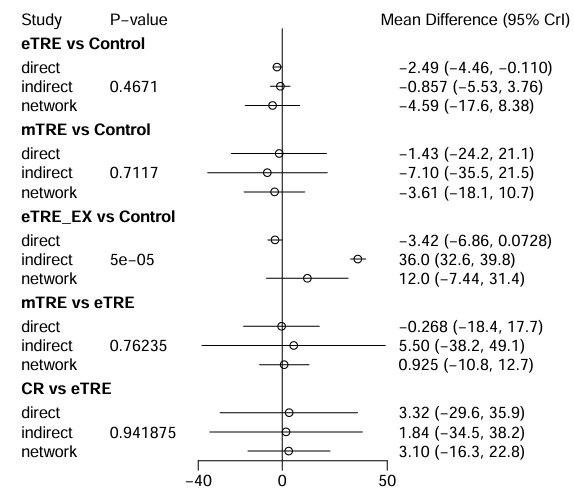

**Footnotes:** CR = calorie restriction, eTRE = early time-restrcited eating, mTRE = midday time-restricted eating, eTRE + EX = early time-restricted eating + exercise.

**Supplementary Figure 28:** Node-splitting approach to assess inconsistency for WHR.

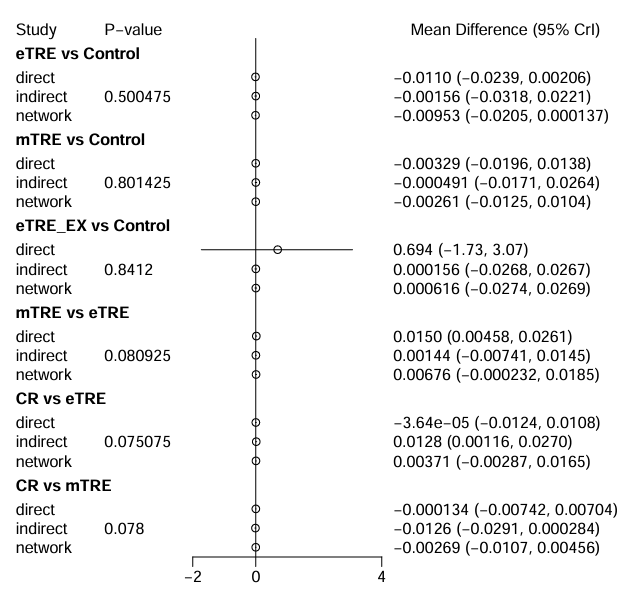
 **Footnotes:** CR = calorie restriction, eTRE = early time-restrcited eating, mTRE = midday time-restricted eating, eTRE + EX = early time-restricted eating + exercise.

.

**Supplementary Figure 29:** Node-splitting approach to assess inconsistency for FBG.

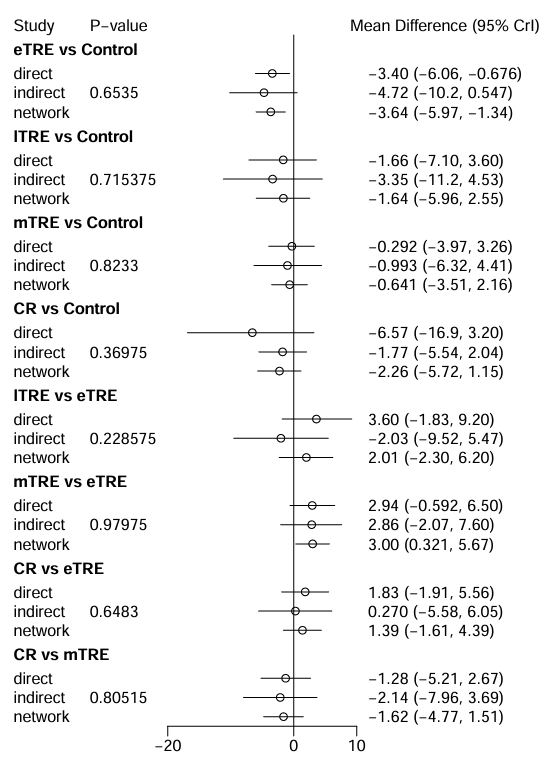

**Footnotes:** CR = calorie restriction, eTRE = early time-restrcited eating, mTRE = midday time-restricted eating, lTRE = late time-restricted eating, eTRE + EX = early time-restricted eating + exercise, mTRE + EX = midday time-restricted eating + exercise.

**Supplementary Figure 30:** Node-splitting approach to assess inconsistency for FBI.

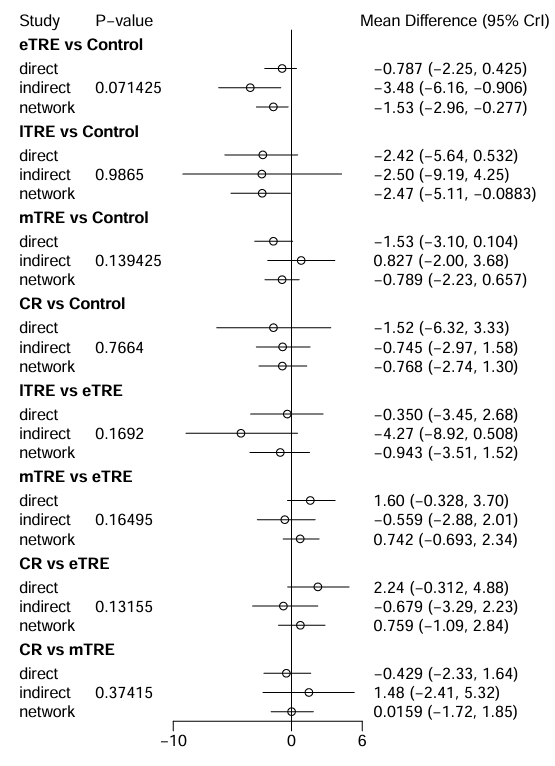

**Footnotes:** CR = calorie restriction, eTRE = early time-restrcited eating, mTRE = midday time-restricted eating, lTRE = late time-restricted eating, eTRE + EX = early time-restricted eating + exercise, mTRE + EX = midday time-restricted eating + exercise.

**Supplementary Figure 31:** Node-splitting approach to assess inconsistency for HOMA-IR

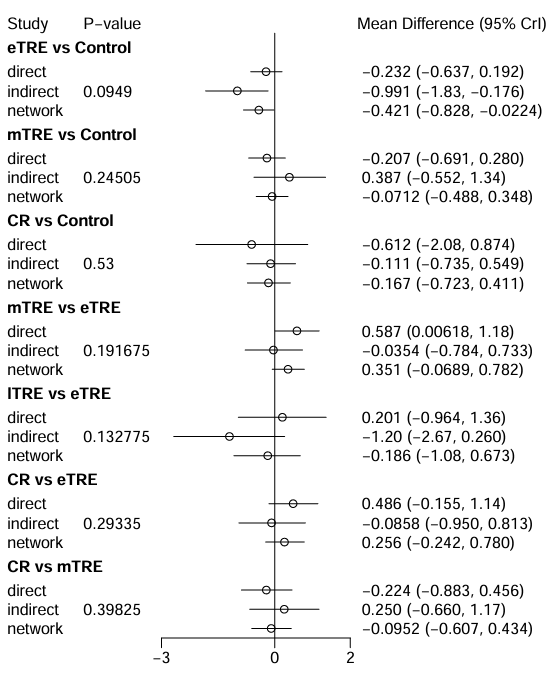

**Footnotes:** CR = calorie restriction, eTRE = early time-restrcited eating, mTRE = midday time-restricted eating, lTRE = late time-restricted eating.

**Supplementary Figure 32:** Node-splitting approach to assess inconsistency for HbA1c:

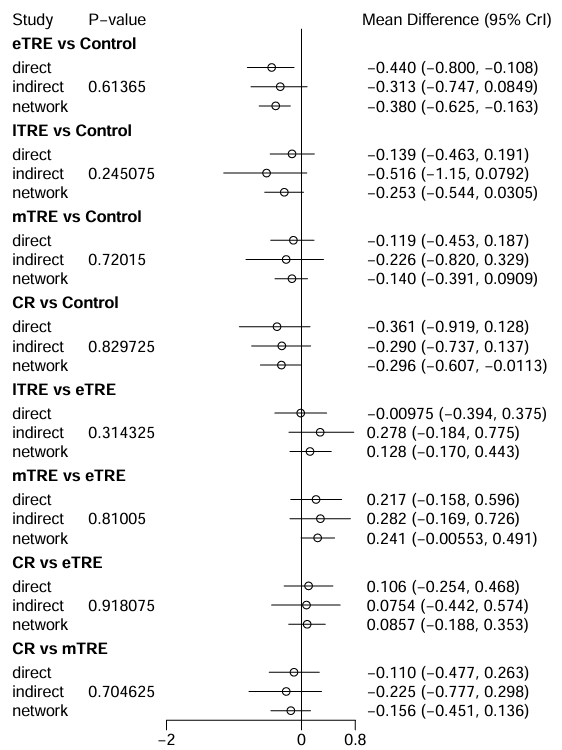

**Footnotes:** CR = calorie restriction, eTRE = early time-restrcited eating, mTRE = midday time-restricted eating, lTRE = late time-restricted eating.

**Supplementary Figure 33:** Node-splitting approach to assess inconsistency for HDL.

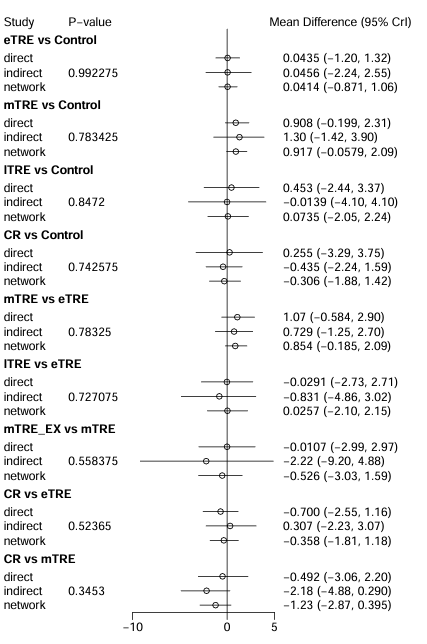

**Footnotes:** CR = calorie restriction, eTRE = early time-restrcited eating, mTRE = midday time-restricted eating, lTRE = late time-restricted eating, mTRE + EX = midday time-restricted eating + exercise.

**Supplementary Figure 34:** Node-splitting approach to assess inconsistency for TG.

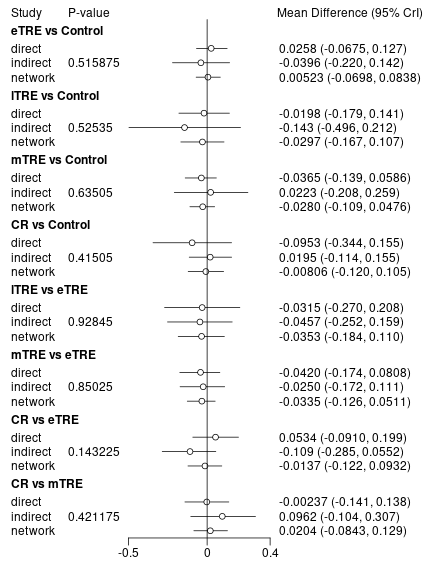

**Footnotes:** CR = calorie restriction, eTRE = early time-restrcited eating, mTRE = midday time-restricted eating, lTRE = late time-restricted eating, mTRE + EX = midday time-restricted eating + exercise

**Supplementary Figure 35:** Node-splitting approach to assess inconsistency for SBP.

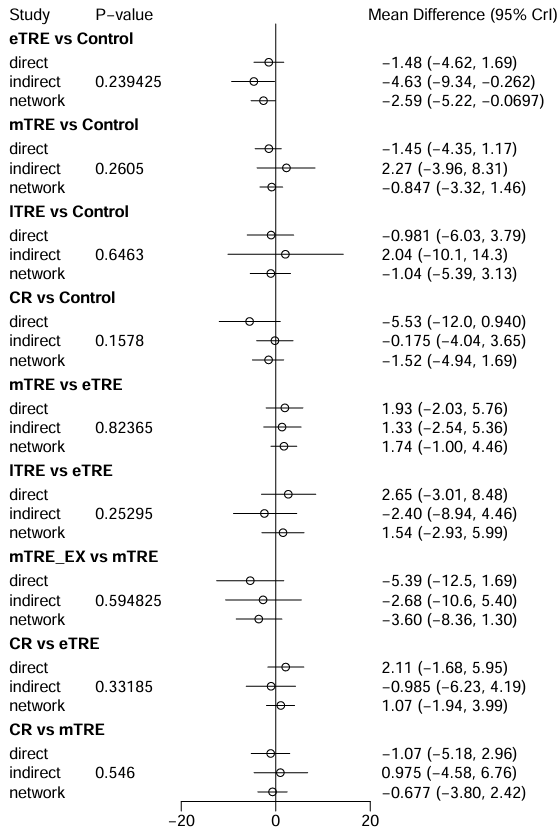

**Footnotes:** CR = calorie restriction, eTRE = early time-restrcited eating, mTRE = midday time-restricted eating, lTRE = late time-restricted eating, mTRE + EX = midday time-restricted eating + exercise.

**Supplementary Figure 36:** Node-splitting approach to assess inconsistency for DBP.

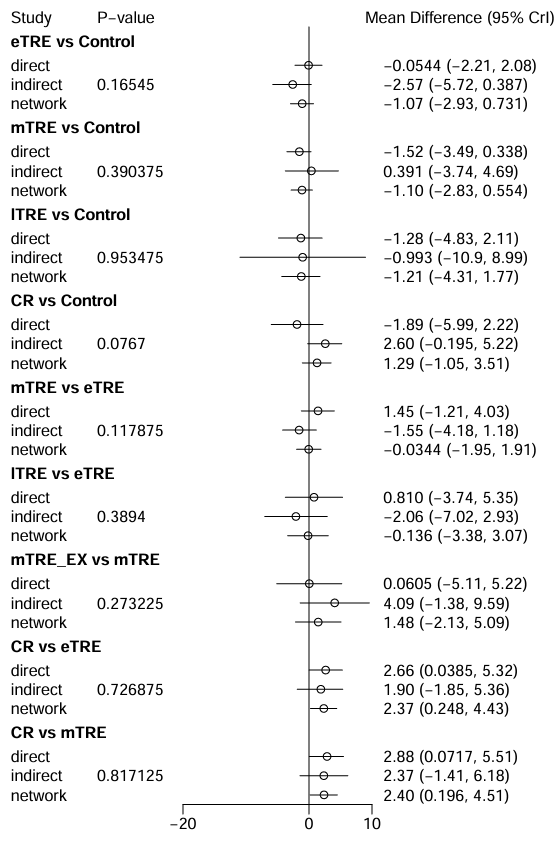

**Footnotes:** CR = calorie restriction, eTRE = early time-restrcited eating, mTRE = midday time-restricted eating, lTRE = late time-restricted eating, mTRE + EX = midday time-restricted eating + exercise
